# Association of COVID-19 with risks of all-cause and cause-specific mortality post-infection: A UK Biobank cohort study

**DOI:** 10.1101/2025.08.01.25332496

**Authors:** Ruoyu Zhang, Yong Xiang, Jinghong Qiu, Hon-Cheong So

## Abstract

**Background:** While SARS-CoV-2 causes multi-organ complications, comprehensive assessments of long-term mortality across organ systems remain limited. This study systematically evaluated COVID-19’s impact on all-cause and cause-specific mortality.

**Methods:** This cohort study followed 467,522 UK Biobank participants (Jan2020-Dec2022). COVID-19 exposure (representing clinically apparent infection) was classified as overall, hospitalized, and non-hospitalized, and compared with reference cohort without documented SARS-CoV-2 records. Post-acute mortality (>30 days post-infection) was assessed using landmark analyses; overall mortality (all mortalities post-infection) served as complementary analyses. Adjusted Cox models estimated risks for 12 organ systems and 47 diseases, with subgroup analyses by key comorbidities and demographics.

**Results:** Post-acute all-cause mortality was elevated in overall (hazard ratio [HR]: 1.50) and hospitalized (HR: 3.06) COVID-19 cohorts, but not non-hospitalized group. COVID-19 infection increased post-acute mortality from circulatory, digestive, genitourinary, neurological, respiratory, and external causes, as well as neoplasms (though elevated cancer mortality without prior diagnoses may reflect detection bias). Hospitalized cases showed elevated risks across 11/11 organ systems and 27/37 diseases; non-hospitalized cases showed increased risks for external-cause and neurological outcomes. Advanced age, atrial fibrillation, chronic kidney disease, and hypertension exacerbated post-acute all-cause mortality; atrial fibrillation also amplified respiratory and neurological risks.

**Conclusion:** Clinically apparent COVID-19 was associated with elevated post-acute and overall mortality across multiple systems, with hospitalized cases exhibiting the broadest risk spectrum. As controls may include unrecorded infections, these estimates are likely conservative. Sustained monitoring is warranted, particularly for older survivors and those with high-risk comorbidities.

## Introduction

The COVID-19 pandemic, caused by coronavirus SARS-CoV-2, has posed an unprecedented challenge to global health. By 5-Jan-2025, the World Health Organization reported over 777 million confirmed-cases and 7 million deaths worldwide attributed to COVID-19 [1]. Emerging evidence indicates that SARS-CoV-2 infection can trigger complications in respiratory, cardiovascular, neurological, renal, gastrointestinal, and other systems [2]. While the increased morbidity/mortality during the acute phase of COVID-19 are well-characterized [3], with recent data suggesting acute mortality still causing a huge burden in early-2025 [4], the post-acute risks across various organs remain incompletely understood. Evaluating the long-term impacts of COVID-19 sequelae is essential for guiding patient care strategies and healthcare resources allocation.

Previous research on post-COVID-19 disorders mainly focused on the prevalence of persistent symptoms and hospitalizations. It was reported that COVID-19, especially hospitalization-required case, confers a greater risk of downstream symptoms/hospitalizations compared to COVID-19-negative individuals [5,6]. In parallel, population-level analyses documented substantial excess pandemic mortality based on historical trends [7,8]. However, by relying on aggregated data, these excess-death estimates cannot fully adjust for individual-level confounders nor accurately attribute deaths to SARS-CoV-2 infection. Although one work [5] quantified mortality risks in COVID-19 patients, they primarily focused on overall mortality rather than specific death causes. While some studies explored post-COVID-19 mortality for neurological [9], respiratory, and cardiovascular outcomes [10], the associations between COVID-19 and mortality spanning a broader spectrum of sequelae remain unclear. Therefore, a systematic and comprehensive analysis of the impact of COVID-19 on mortality across all body systems is urgently needed.

To address this gap, we conducted a longitudinal analysis by following a prospective UK Biobank (UKBB) cohort from 31-Jan-2020 to 19-Dec-2022. We leveraged Clinical-Classifications-Software-Refined (CCSR) to facilitate an inclusive grouping of ICD-10 diagnoses, thereby classifying causes of post-COVID-19 deaths into 12 composite organ-system and 47 single-disease outcomes. We primarily estimated the associations between COVID-19 (overall, hospitalized, and non-hospitalized) and post-acute mortality (≥31 days post-infection) due to these disorders, with overall mortality (all mortalities post-infection, including acute and post-acute phases) also presented for reference and as complementary analyses. For each single outcome, deaths were further stratified by prior history of that condition. Additionally, we performed subgroup analyses stratified by various characteristics to explore potential differences in the mortality risks of different sequelae across subgroups.

This study makes several novel contributions to the understanding of post-COVID-19 mortality. To our knowledge, it is the first to comprehensively examine post-acute and overall mortality across a wide range of diseases spanning all organ systems, identifying novel associations between COVID-19 and elevated risk of death from digestive, genitourinary, and external causes. Furthermore, our work is the first to stratify COVID-19 mortality by patients’ clinical characteristics, revealing how pre-existing conditions amplify specific risks. For example, we found that pre-existing atrial fibrillation significantly increases post-acute/overall mortality from neurological and respiratory diseases. These associations were not previously reported.

## Methods

### Study design and setting

This prospective cohort study used data from UKBB (project #28732), which tracks the electronic health records of ∼500,000 participants aged 50-87 years [11]. To evaluate the hazard ratios (HRs) for post-COVID-19 deaths, UKBB sample was followed from 31-Jan-2020 (date of the first UK-confirmed COVID-19 case) to 19-Dec-2022 (last date for available records during analysis). Totally 467,522 individuals alive at the start of follow-up were included for subsequent analysis.

### Data Sources, Outcomes and Covariates

Participants’ baseline characteristics, comorbidities, SARS-CoV-2 infection status, and mortality records were ascertained from UKBB-linked electronic health records and national death registries. Our primary study outcomes were post-acute (≥31 days after SARS-CoV-2 infection) all-cause and cause-specific mortality. We focused solely on the primary cause of death to avoid confounding from secondary conditions. The ICD-10-coded causes of mortality were categorized using CCSR [12]. Overall mortality across the entire follow-up (including days 0-30) was additionally assessed as complementary analysis using the same outcome definitions. By restricting outcomes with ≥5 deaths in both COVID-19 exposure and reference cohorts, up to 12 composite (organ-system) outcomes and 47 single (CCSR-defined-disease) categories were included (Table_S1). For each single outcome, we also performed analysis stratified by whether patients had a pre-existing diagnosis of the same condition. To adjust for baseline differences between cohorts, we selected a set of covariates [6] including sociodemographic, lifestyle, biochemical and clinical factors, as well as pre-existing comorbidities and vaccination status (see Supplementary_methods for full list). Imputation of missing values was conducted by *missRanger* [13].

### Cohort

In our primary analysis, the COVID-19 exposure cohort comprised individuals with a single episode of SARS-CoV-2 infection, where reinfection was censored at the second infection date. Further sensitivity analysis was also performed for reinfections (see below). Based on initial infection severity, the exposure was divided into hospitalized and non-hospitalized groups. Follow-up for COVID-19-infected individuals commenced on the first positive SARS-CoV-2 test date (*T_0_*) and continued until death, the first reinfection, or 19-Dec-2022, whichever occurred first. For the primary post-acute mortality outcome, a landmark analysis [14] was performed. Individuals who died within 30 days after *T_0_* were excluded, and follow-up began at *T_0_+30* days (*T_landmark_*). Deaths from causes other than the outcome of interest were censored.

The reference cohort comprised subjects without documented history of SARS-CoV-2. As this cohort was not defined by confirmed seronegative status, it may include individuals with unrecorded mild/asymptomatic infections. To mitigate potential survival bias from imbalanced follow-up length and ensure comparability, each control participant was assigned a pseudo-index date (*T’_0_*) drawn randomly from the *T_0_*distribution observed in the exposure cohort, following the prescription time distribution matching method as described in [15]. Reference individuals were followed from their assigned *T’_0_* until death or 19-Dec-2022. For the post-acute analysis, the same 30-day landmark was applied to reference (*T’_landmark_ = T’_0_+30 days*).

### Statistical Analyses

Proportional hazards Cox regression was employed to model time to the target death event. Primary analyses focused on post-acute mortality using follow-up starting at *T_landmark_ or T’_landmark_*; overall mortality (including days 0-30) was additionally evaluated as supplementary analyses. To mitigate potential convergence issues, we performed variable selection for covariates included in the Cox model (see Supplementary_methods) [16]. False discovery rate (FDR) was used to control for multiple testing, with FDR-adjusted p-values < 0.05 considered statistically significant. Overall analytic workflow is illustrated in Fig. S1.

HR for each (post-acute/overall mortality) outcome was estimated by comparing the exposure (overall, hospitalized, and non-hospitalized) to the reference cohort; as the reference may include unrecorded infections, these estimates reflect mortality risk associated with clinically apparent COVID-19 relative to the general population without known infection. For individual (post-acute/overall) outcomes, HRs were also separately calculated for individuals with and without a prior history of the corresponding sequela. For subgroup analyses (post-acute/overall), we assessed risk heterogeneity by computing the ratio of hazard ratios (RHR) across subgroups stratified by multiple demographic/comorbidity characteristics.

#### Additional analyses

Additional analyses evaluated the robustness of primary post-acute organ-system findings for overall COVID-19 exposure, addressing three potential concerns: (1) misclassification of undocumented infections, through quantitative bias analysis (QBA) under scenarios informed by external estimates of cumulative UK COVID-19 prevalence [17], and use of a reference group restricted to those tested negative for COVID-19 infection; (2) effects of vaccination status on mortalities, through stratified analyses and testing for interaction with COVID-19 infection; (3) reinfection-based censoring, by extending follow-up beyond reinfection, and stratifying the exposure cohort by number of infections, with follow-up beginning at the first infection. Further methodological details are provided in Supplementary_methods.

## Results

For the post-acute analysis, after excluding participants (COVID-19: 446; reference: 345) who died within the 30-day landmark, the COVID-19 cohort included 112,611 participants (median follow-up: 247 days), comprising 16,260 hospitalized (median 292 days) and 96,351 non-hospitalized (median 241 days) individuals; 4,942 (4.4%) experienced reinfection during follow-up. The reference cohort included 354,120 participants (median 335 days), of whom 169,400 (47.8%) received SARS-CoV-2 testing but never tested positive. Detailed cohort characteristics (including overall-mortality analysis) are presented in Table_S2.

As summarized in Table_S3a, post-acute deaths totaled 2,888 (2.56%) in the overall COVID-19-infected cohort, 2,382 (14.65%) in hospitalized, 506 (0.53%) in non-hospitalized, and 5,317 (1.50%) in reference participants. Neoplasms, circulatory, neurological, and respiratory disorders were the most common mortality causes across all cohorts. Corresponding overall-mortality distributions are provided in Table_S3b.

### Mortality risks of composite organ-system outcomes

Compared to the reference (general population without documented infection), *post-acute* all-cause mortality was significantly elevated in overall COVID-19-infected (HR: 1.50, 95% confidence interval(CI): 1.42-1.58) and hospitalized (3.06, 2.87-3.26) groups, but not in the non-hospitalized (1.05, 0.98-1.14) group (Fig. 1a, Table_1). In the *overall*-mortality analysis, all three exposure cohorts showed increased all-cause deaths (overall COVID-19-infected: 2.39, 2.29-2.50; hospitalized: 6.29, 5.99-6.61; non-hospitalized: 1.23, 1.15-1.32) (Fig. S2a, Table_1).

**Figure 1.**
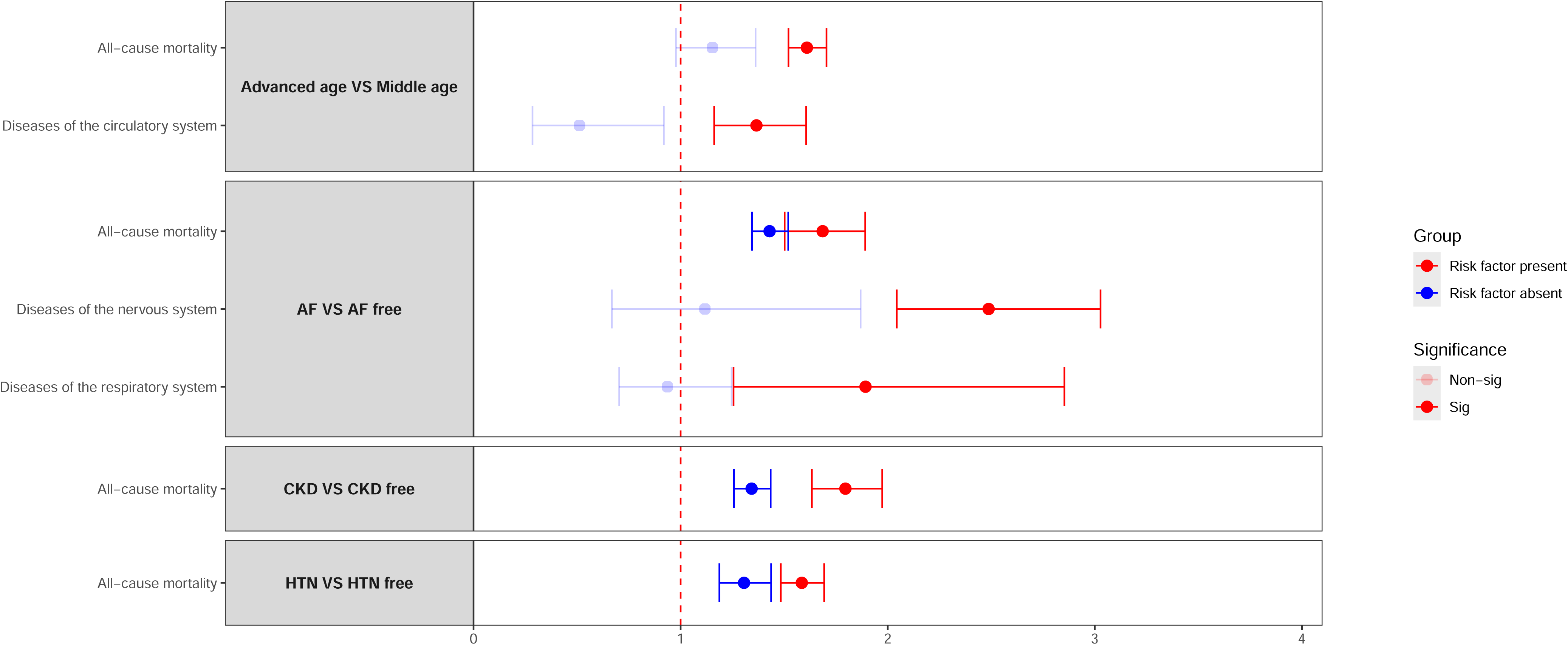
Associations Between All-Cause and Cause-Specific Post-acute Mortality Risks in Different Organ Systems and Overall/Hospitalized/Non-hospitalized COVID-19. **Legend:** The vertical red dashed line represents the line of no effect (hazard ratio (HR) = 1). X-axis indicates the HR of post-acute mortality after COVID-19 infection. Y-axis indicates each COVID-19 disease severity. Confidence intervals are also shown in the figure. Solid bars indicate significance, and transparent bars indicate non-significant associations. We only present the results if the number of events ≥5 for both COVID-19 exposure and reference cohort

**Table 1.** Associations Between All-Cause and Cause-Specific Mortality Risks in Different Organ Systems and Overall/Hospitalized/Non-hospitalized COVID-19.

| Analysis | Outcome | Post-acute Mortality<sup>1</sup> |  | Overall Mortality<sup>2</sup> |  | Organ System Description |
| --- | --- | --- | --- | --- | --- | --- |
|  |  | HR (95% CI) | p.adj<sup>3</sup> | HR (95% CI) | p.adj<sup>3</sup> |  |
| Overall COVID-19<br>vs. reference | All-cause mortality | **1.50 (1.42 - 1.58)** | **2.83E-48** | **2.39 (2.29 - 2.50)** | **< 2.23E-308** |  |
|  | BLD |  | N/A<sup>4</sup> | **7.85 (1.80 - 34.33)** | **8.95E-03** | Diseases of the blood and blood forming organs and certain disorders involving the immune mechanism |
|  | CIR | **1.22 (1.05 - 1.43)** | **2.17E-02** | **1.45 (1.26 - 1.66)** | **7.12E-07** | Diseases of the circulatory system |
|  | DIG | **1.50 (1.04 - 2.16)** | **4.26E-02** | **1.98 (1.45 - 2.70)** | **4.38E-05** | Diseases of the digestive system |
|  | END | 1.61 (0.75 - 3.46) | 2.88E-01 | 1.75 (0.86 - 3.54) | 1.32E-01 | Endocrine nutritional and metabolic diseases |
|  | EXT | **3.75 (1.98 - 7.09)** | **1.45E-04** | **3.42 (1.89 - 6.21)** | **1.13E-04** | External cause codes |
|  | GEN | **2.19 (1.29 - 3.71)** | **8.51E-03** | **2.54 (1.58 - 4.09)** | **2.17E-04** | Diseases of the genitourinary system |
|  | INF | 1.25 (0.79 - 1.98) | 3.77E-01 | 1.46 (0.97 - 2.21) | 9.12E-02 | Certain infectious and parasitic diseases |
|  | INJ | 1.05 (0.52 - 2.10) | 8.94E-01 | 1.31 (0.71 - 2.41) | 3.91E-01 | Injury poisoning and certain other consequences of external causes |
|  | MUS | 1.61 (0.73 - 3.58) | 2.88E-01 | 1.84 (0.88 - 3.83) | 1.24E-01 | Diseases of the musculoskeletal system and connective tissue |
|  | NEO | **1.38 (1.26 - 1.51)** | **6.22E-12** | **1.53 (1.41 - 1.67)** | **1.03E-23** | Neoplasms |
|  | NVS | **2.13 (1.78 - 2.56)** | **2.66E-15** | **2.20 (1.85 - 2.62)** | **1.80E-18** | Diseases of the nervous system |
|  | RSP | **1.21 (1.04 - 1.41)** | **2.62E-02** | **1.39 (1.12 - 1.72)** | **4.00E-03** | Diseases of the respiratory system |
| Hospitalized<br>COVID-19<br>vs. reference | All-cause mortality | **3.06 (2.87 - 3.26)** | **1.25E-258** | **6.29 (5.99 - 6.61)** | **< 2.23E-308** |  |
|  | CIR | **2.37 (1.96 - 2.86)** | **2.20E-18** | **3.16 (2.68 - 3.72)** | **4.92E-43** | Diseases of the circulatory system |
|  | DIG | **3.07 (1.98 - 4.76)** | **7.42E-07** | **4.98 (3.51 - 7.07)** | **7.22E-19** | Diseases of the digestive system |
|  | END | **4.35 (1.70 - 11.10)** | **2.10E-03** | **4.71 (2.02 - 10.99)** | **3.33E-04** | Endocrine nutritional and metabolic diseases |
|  | EXT | **11.08 (5.35 - 22.96)** | **2.29E-10** | **10.36 (5.27 - 20.37)** | **1.63E-11** | External cause codes |
|  | GEN | **5.10 (2.88 - 9.06)** | **4.28E-08** | **6.16 (3.69 - 10.27)** | **5.73E-12** | Diseases of the genitourinary system |
|  | INF | **4.88 (2.83 - 8.43)** | **2.65E-08** | **5.75 (3.57 - 9.26)** | **1.29E-12** | Certain infectious and parasitic diseases |
|  | INJ | **4.99 (2.39 - 10.40)** | **1.94E-05** | **6.20 (3.27 - 11.77)** | **2.84E-08** | Injury poisoning and certain other consequences of external causes |
|  | MUS | **8.57 (3.50 - 20.99)** | **3.52E-06** | **9.06 (3.97 - 20.65)** | **1.73E-07** | Diseases of the musculoskeletal system and connective tissue |
|  | NEO<sup>5</sup> | **3.10 (2.79 - 3.45)** | **3.67E-96** | **3.81 (3.47 - 4.19)** | **4.34E-167** | Neoplasms |
|  | NVS | **5.06 (4.03 - 6.35)** | **7.86E-44** | **5.61 (4.55 - 6.92)** | **7.48E-58** | Diseases of the nervous system |
|  | RSP | **1.87 (1.41 - 2.47)** | **1.44E-05** | **2.47 (1.91 - 3.19)** | **6.11E-12** | Diseases of the respiratory system |
| Non-hospitalized<br>COVID-19<br>vs. reference | All-cause mortality | 1.05 (0.98 - 1.14) | 4.70E-01 | **1.23 (1.15 - 1.32)** | **2.26E-08** |  |
|  | CIR | 0.74 (0.37 - 1.49) | 5.61E-01 | 1.04 (0.86 - 1.26) | 7.62E-01 | Diseases of the circulatory system |
|  | DIG | 0.90 (0.53 - 1.53) | 7.07E-01 | 1.41 (0.94 - 2.13) | 2.51E-01 | Diseases of the digestive system |
|  | EXT | **2.77 (1.29 - 5.96)** | **4.63E-02** | **2.72 (1.35 - 5.49)** | **1.69E-02** | External cause codes |
|  | INF | 0.65 (0.34 - 1.23) | 4.70E-01 | 0.76 (0.43 - 1.34) | 4.81E-01 | Certain infectious and parasitic diseases |
|  | MUS | 0.67 (0.21 - 2.18) | 5.61E-01 | 0.85 (0.30 - 2.43) | 7.67E-01 | Diseases of the musculoskeletal system and connective tissue |
|  | NEO | 0.76 (0.38 - 1.52) | 5.61E-01 | 0.93 (0.82 - 1.05) | 4.10E-01 | Neoplasms |
|  | NVS | **1.37 (1.09 - 1.74)** | **4.63E-02** | **1.47 (1.18 - 1.82)** | **2.81E-03** | Diseases of the nervous system |

1. Post-acute Mortality measures the risk of death by setting the follow-up start date as 30 days after the date of first positive SARS-CoV-2 test. 2. Overall Mortality measures the risk of death by setting the follow-up start date as the date of first positive SARS-CoV-2 test. 3. FDR correction was separately performed for each COVID-19 exposure cohort; Statistically significant associations ( $p_{\text{adj}} < 0.05$ ) were labelled in bold face. 4. N/A indicates that the analysis for target outcome cannot proceed due to insufficient number ( $<5$ ) of events in either the COVID-19 exposure or reference cohort. 5. Elevated neoplasm mortality, especially among hospitalized patients without prior cancer history, should be interpreted with caution, as this finding is more likely attributable to detection bias (unmasking of pre-existing undiagnosed malignancies) rather than representing a direct post-infection sequela of COVID-19. See Discussion for detailed interpretation.
Please refer to the following abbreviations used in all main tables:
HR: hazard ratio; 95% CI represents the lower and upper 95% confidence interval of HR; $p_{\text{adj}}^3$ : FDR-adjusted p-value.
CCSR: Clinical Classifications Software Refined, a method to classify disease categories.
$\text{HR}(\text{CI})_{\text{present}}$ : Hazard ratio and 95% confidence interval for the risk factor present subgroup with the examined characteristics; $p_{\text{adj}}_{\text{present}}$ : FDR-adjusted p-value for the risk factor present subgroup;
$\text{HR}(\text{CI})_{\text{absent}}$ : Hazard ratio and 95% confidence interval for the risk factor absent subgroup without the examined characteristics; $p_{\text{adj}}_{\text{absent}}$ : FDR-adjusted p-value for the risk factor absent subgroup;
$\text{Sig}_{\text{present}}$ and $\text{Sig}_{\text{absent}}$ : \* indicates an FDR-adjusted p-value between 0.01 and 0.05, \*\* indicates between 0.001 and 0.01, and \*\*\* indicates a value smaller than 0.001 for the risk factor present and absent subgroups, respectively.
RHR: Ratio of hazard ratio for comparisons between the risk factor present and absent subgroups.
$p_{\text{adj}}_{\text{RHR}}^*$ : FDR-adjusted p-value for comparisons between the risk factor present and absent subgroups, where a $p_{\text{adj}}_{\text{RHR}} < 0.05$ indicates significant differences.

For organ-system outcomes, a wide range of significant associations were found, demonstrating increased mortality risks post infection (Fig. 1b, Table_1). Hospitalized COVID-19 exhibited significant increased post-acute mortality in all (11/11) examined systems. Overall COVID-19 infection was associated with significantly elevated post-acute mortality in 7 of 11 systems (circulatory, digestive, external-cause, genitourinary, neoplasms, nervous, and respiratory). For the non-hospitalized cohort, only external-cause and nervous-system post-acute mortality (2/7) reached significance. Overall-mortality analyses showed generally larger effect estimates (except for external causes), with a largely similar pattern of significant outcomes (Fig. S2b, Table_1).

### Mortality risks of individual outcomes for overall COVID-19 exposure

Among 42 individual disorders investigated, COVID-19 infection significantly increased post-acute mortality for 8 (19.0%) conditions (Table_2a, Fig. S3), including (ranked from the largest to smallest HR per system):

- Circulatory: Peripheral/visceral vascular disease.
- External-cause injuries: Accidental/unintentional intent of injury, and subsequent encounter of external cause codes.
- Genitourinary: Urinary tract infection.
- Neoplasm: Bile duct cancer, and unspecified malignant neoplasm.
- Neurological: Neurocognitive disorders, and Parkinson’s disease.

In the overall-mortality analysis, more conditions reached significance (17/47, 36.2%; Table_2a, Fig. S4), with generally larger HRs.

**Table 2.** Associations Between Cause-Specific Mortality Risks in Single CCSR Disorders and COVID-19.

| *a. Compared to reference cohort without known infection, associations of overall COVID-19 with mortality due to various disorders (i.e. Overall COVID-19 vs. reference)* |  |  |  |  |  |
| --- | --- | --- | --- | --- | --- |
| Outcome | Post-acute Mortality<sup>1</sup> |  | Overall Mortality<sup>2</sup> |  | CCSR Category Description |
|  | HR (95% CI) | p.adj<sup>3</sup> | HR (95% CI) | p.adj<sup>3</sup> |  |
| CIR001 |  | N/A<sup>4</sup> | 0.94 (0.33 - 2.68) | 9.28E-01 | Chronic rheumatic heart disease |
| CIR003 | 1.13 (0.48 - 2.65) | 8.44E-01 | 1.47 (0.70 - 3.09) | 4.04E-01 | Nonrheumatic and unspecified valve disorders |
| CIR004 | 1.81 (0.64 - 5.16) | 4.12E-01 | **3.12 (1.47 - 6.62)** | **1.26E-02** | Endocarditis and endocardial disease |
| CIR005 | 0.68 (0.29 - 1.62) | 5.10E-01 | 0.96 (0.46 - 2.00) | 9.28E-01 | Myocarditis and cardiomyopathy |
| CIR008 | 1.38 (0.72 - 2.66) | 4.50E-01 | 1.54 (0.84 - 2.80) | 2.51E-01 | Hypertension with complications and secondary hypertension |
| CIR009 | 0.78 (0.55 - 1.11) | 2.85E-01 | 0.95 (0.70 - 1.29) | 8.73E-01 | Acute myocardial infarction |
| CIR011 | 1.34 (0.97 - 1.85) | 2.18E-01 | **1.51 (1.12 - 2.03)** | **2.22E-02** | Coronary atherosclerosis and other heart disease |
| CIR019 | 1.93 (0.97 - 3.85) | 2.00E-01 | **2.23 (1.18 - 4.22)** | **3.82E-02** | Heart failure |
| CIR020 |  | N/A | 0.94 (0.33 - 2.62) | 9.28E-01 | Cerebral infarction |
| CIR021 | 1.19 (0.75 - 1.88) | 5.42E-01 | 1.37 (0.91 - 2.08) | 2.18E-01 | Acute hemorrhagic cerebrovascular disease |
| CIR024 | 1.69 (0.63 - 4.52) | 4.41E-01 | 2.21 (0.90 - 5.44) | 1.45E-01 | Other and ill-defined cerebrovascular disease |
| CIR026 | **4.11 (2.13 - 7.94)** | **2.76E-04** | **4.40 (2.40 - 8.05)** | **2.66E-05** | Peripheral and visceral vascular disease |
| CIR029 |  | N/A | 1.18 (0.43 - 3.23) | 8.73E-01 | Aortic; peripheral; and visceral artery aneurysms |
| DIG006 |  | N/A | 2.26 (0.70 - 7.24) | 2.58E-01 | Gastrointestinal and biliary perforation |
| DIG012 | 1.13 (0.35 - 3.63) | 8.78E-01 | 1.16 (0.41 - 3.26) | 8.82E-01 | Intestinal obstruction and ileus |
| DIG017 | 2.17 (0.77 - 6.11) | 2.70E-01 | **3.26 (1.36 - 7.83)** | **2.49E-02** | Biliary tract disease |
| DIG019 | 1.42 (0.57 - 3.51) | 5.42E-01 | 1.61 (0.71 - 3.64) | 3.59E-01 | Other specified and unspecified liver disease |
| DIG021 | 2.58 (0.81 - 8.20) | 2.59E-01 | 1.74 (0.60 - 5.08) | 4.04E-01 | Gastrointestinal hemorrhage |
| DIG025 | 2.12 (0.92 - 4.87) | 2.18E-01 | **2.79 (1.36 - 5.75)** | **1.79E-02** | Other specified and unspecified gastrointestinal disorders |
| END016 | 2.07 (0.81 - 5.33) | 2.70E-01 | 2.44 (1.03 - 5.79) | 8.78E-02 | Other specified and unspecified nutritional and metabolic disorders |
| EXT020 | **4.58 (2.30 - 9.14)** | **2.11E-04** | **4.04 (2.10 - 7.75)** | **3.36E-04** | External cause codes: intent of injury, accidental/unintentional |
| EXT029 | **4.00 (2.05 - 7.82)** | **4.23E-04** | **3.59 (1.90 - 6.79)** | **5.85E-04** | External cause codes: subsequent encounter |
| GEN003 | 1.95 (0.70 - 5.39) | 3.22E-01 | 2.35 (0.99 - 5.59) | 9.90E-02 | Chronic kidney disease |
| GEN004 | **3.11 (1.52 - 6.39)** | **1.18E-02** | **4.00 (2.04 - 7.83)** | **4.96E-04** | Urinary tract infections |
| INF002 | 2.19 (0.77 - 6.21) | 2.70E-01 | 2.30 (0.92 - 5.75) | 1.33E-01 | Septicemia |
| INF003 | 2.07 (0.89 - 4.85) | 2.42E-01 | 2.22 (1.02 - 4.80) | 8.78E-02 | Bacterial infections |
| NEO012 | 0.86 (0.52 - 1.41) | 6.20E-01 | 0.94 (0.60 - 1.48) | 8.82E-01 | Gastrointestinal cancers - esophagus |
| NEO013 | 1.40 (0.72 - 2.76) | 4.50E-01 | 1.87 (1.05 - 3.32) | 7.24E-02 | Gastrointestinal cancers - stomach |
| NEO015 | 1.37 (1.02 - 1.84) | 1.62E-01 | **1.55 (1.18 - 2.03)** | **7.73E-03** | Gastrointestinal cancers - colorectal |
| NEO017 | 0.95 (0.43 - 2.08) | 8.99E-01 | 0.97 (0.47 - 1.98) | 9.31E-01 | Gastrointestinal cancers - liver |
| NEO018 | **2.30 (1.24 - 4.25)** | **4.10E-02** | **2.45 (1.38 - 4.35)** | **1.03E-02** | Gastrointestinal cancers - bile duct |
| NEO024 | 0.92 (0.28 - 2.99) | 8.99E-01 | 0.92 (0.28 - 2.99) | 9.28E-01 | Sarcoma |
| NEO025 | 1.90 (1.02 - 3.55) | 1.79E-01 | 1.98 (1.10 - 3.56) | 5.76E-02 | Skin cancers - melanoma |
| NEO043 | 1.23 (0.71 - 2.15) | 5.42E-01 | 1.75 (1.08 - 2.84) | 5.82E-02 | Urinary system cancers - bladder |
| NEO048 | 1.27 (0.79 - 2.05) | 4.50E-01 | 1.33 (0.85 - 2.10) | 3.08E-01 | Nervous system cancers - brain |
| NEO051 | 0.91 (0.62 - 1.31) | 6.64E-01 | 1.15 (0.83 - 1.59) | 5.22E-01 | Endocrine system cancers - pancreas |
| NEO067 | 2.20 (0.79 - 6.16) | 2.70E-01 | 2.47 (1.00 - 6.14) | 9.82E-02 | Mesothelioma |
| NEO068 | 1.89 (0.73 - 4.91) | 3.21E-01 | 1.61 (0.66 - 3.92) | 4.04E-01 | Myelodysplastic syndrome (MDS) |
| NEO071 | **2.13 (1.35 - 3.37)** | **8.36E-03** | **2.16 (1.40 - 3.32)** | **2.75E-03** | Malignant neoplasm, unspecified |
| NVS004 | **2.29 (1.64 - 3.19)** | **2.39E-05** | **2.17 (1.59 - 2.98)** | **2.66E-05** | Parkinson`s disease |
| NVS005 | 1.91 (0.78 - 4.65) | 2.85E-01 | 1.90 (0.82 - 4.38) | 2.18E-01 | Multiple sclerosis |
| NVS006 | 1.79 (0.87 - 3.66) | 2.59E-01 | 2.11 (1.06 - 4.19) | 7.24E-02 | Other specified hereditary and degenerative nervous system conditions |
| NVS011 | **2.32 (1.81 - 2.97)** | **1.18E-09** | **2.48 (1.96 - 3.13)** | **1.93E-12** | Neurocognitive disorders |
| RSP002 | 1.62 (0.98 - 2.68) | 2.00E-01 | **1.98 (1.26 - 3.11)** | **1.26E-02** | Pneumonia (except that caused by tuberculosis) |
| RSP008 | 1.36 (1.00 - 1.85) | 1.90E-01 | **1.53 (1.15 - 2.04)** | **1.44E-02** | Chronic obstructive pulmonary disease and bronchiectasis |
| RSP010 |  | N/A | **6.56 (1.41 - 30.51)** | **4.36E-02** | Aspiration pneumonitis |
| RSP016 | 0.76 (0.39 - 1.49) | 5.32E-01 | 0.84 (0.45 - 1.58) | 7.35E-01 | Other specified and unspecified lower respiratory disease |

| Outcome | Post-acute Mortality<sup>1</sup> |  | Overall Mortality<sup>2</sup> |  | CCSR Category Description |
| --- | --- | --- | --- | --- | --- |
|  | HR (95% CI) | p.adj<sup>3</sup> | HR (95% CI) | p.adj<sup>3</sup> |  |
| CIR003 | 1.39 (0.46 - 4.17) | 5.90E-01 | 2.09 (0.81 - 5.41) | 1.43E-01 | Nonrheumatic and unspecified valve disorders |
| CIR004 |  | N/A<sup>4</sup> | **9.79 (4.25 - 22.56)** | **2.47E-07** | Endocarditis and endocardial disease |
| CIR005 |  | N/A | **3.23 (1.36 - 7.65)** | **9.14E-03** | Myocarditis and cardiomyopathy |
| CIR008 | 1.99 (0.81 - 4.88) | 1.53E-01 | **3.06 (1.45 - 6.45)** | **4.27E-03** | Hypertension with complications and secondary hypertension |
| CIR009 | 1.54 (0.99 - 2.39) | 6.95E-02 | **2.21 (1.54 - 3.16)** | **3.26E-05** | Acute myocardial infarction |
| CIR011 | **2.71 (1.81 - 4.04)** | **3.64E-06** | **3.29 (2.29 - 4.72)** | **6.07E-10** | Coronary atherosclerosis and other heart disease |
| CIR019 | **4.53 (2.06 - 9.93)** | **4.14E-04** | **5.49 (2.69 - 11.20)** | **6.89E-06** | Heart failure |
| CIR020 |  | N/A | 2.59 (0.87 - 7.67) | 9.84E-02 | Cerebral infarction |
| CIR021 | 1.57 (0.75 - 3.28) | 2.47E-01 | **2.56 (1.46 - 4.49)** | **1.45E-03** | Acute hemorrhagic cerebrovascular disease |
| CIR024 | 2.93 (1.01 - 8.54) | 6.39E-02 | **4.13 (1.58 - 10.78)** | **4.76E-03** | Other and ill-defined cerebrovascular disease |
| CIR026 | **8.61 (4.21 - 17.58)** | **1.76E-08** | **10.01 (5.22 - 19.18)** | **3.45E-11** | Peripheral and visceral vascular disease |
| DIG012 | 1.47 (0.36 - 5.93) | 6.08E-01 | 1.87 (0.56 - 6.19) | 3.23E-01 | Intestinal obstruction and ileus |
| DIG017 | **5.99 (1.82 - 19.71)** | **5.90E-03** | **9.99 (3.69 - 27.08)** | **1.36E-05** | Biliary tract disease |
| DIG019 | **3.96 (1.40 - 11.21)** | **1.36E-02** | **4.90 (1.96 - 12.23)** | **9.54E-04** | Other specified and unspecified liver disease |
| DIG025 | 2.86 (0.99 - 8.27) | 6.75E-02 | 5.29 (2.28 - 12.27) | **1.83E-04** | Other specified and unspecified gastrointestinal disorders |
| END016 | **4.76 (1.51 - 14.97)** | **1.13E-02** | **6.27 (2.29 - 17.17)** | **5.54E-04** | Other specified and unspecified nutritional and metabolic disorders |
| EXT020 | **12.01 (5.42 - 26.60)** | **5.51E-09** | **10.53 (4.95 - 22.43)** | **3.65E-09** | External cause codes: intent of injury, accidental/unintentional |
| EXT029 | **11.91 (5.52 - 25.66)** | **1.91E-09** | **10.53 (5.06 - 21.94)** | **1.24E-09** | External cause codes: subsequent encounter |
| GEN003 | **4.21 (1.35 - 13.13)** | **1.79E-02** | **5.25 (2.03 - 13.60)** | **9.50E-04** | Chronic kidney disease |
| GEN004 | **7.20 (3.35 - 15.47)** | **1.74E-06** | **9.84 (4.86 - 19.93)** | **1.06E-09** | Urinary tract infections |
| INF002 |  | N/A | **4.57 (1.51 - 13.84)** | **8.72E-03** | Septicemia |
| INF003 | **4.45 (1.49 - 13.30)** | **1.13E-02** | **5.09 (1.99 - 13.01)** | **9.54E-04** | Bacterial infections |
| NEO012<sup>5</sup> | **2.33 (1.31 - 4.16)** | **6.80E-03** | **2.72 (1.62 - 4.55)** | **2.41E-04** | Gastrointestinal cancers - esophagus |
| NEO013<sup>5</sup> | **4.37 (2.03 - 9.42)** | **4.14E-04** | **6.56 (3.49 - 12.35)** | **1.65E-08** | Gastrointestinal cancers - stomach |
| NEO015<sup>5</sup> | **3.40 (2.34 - 4.95)** | **1.50E-09** | **4.27 (3.07 - 5.94)** | **6.88E-17** | Gastrointestinal cancers - colorectal |
| NEO017 | 1.20 (0.38 - 3.84) | 7.58E-01 | 1.53 (0.57 - 4.08) | 4.03E-01 | Gastrointestinal cancers - liver |
| NEO018<sup>5</sup> | **7.52 (3.84 - 14.72)** | **1.76E-08** | **8.10 (4.37 - 15.01)** | **1.83E-10** | Gastrointestinal cancers - bile duct |
| NEO024 | 2.45 (0.62 - 9.72) | 2.28E-01 | 2.45 (0.62 - 9.72) | 2.18E-01 | Sarcoma |
| NEO025<sup>5</sup> | **4.26 (1.93 - 9.43)** | **7.98E-04** | **5.18 (2.51 - 10.71)** | **1.94E-05** | Skin cancers - melanoma |
| NEO043<sup>5</sup> | **2.56 (1.36 - 4.83)** | **6.30E-03** | **4.27 (2.50 - 7.29)** | **2.61E-07** | Urinary system cancers - bladder |
| NEO048<sup>5</sup> | **2.59 (1.46 - 4.59)** | **2.42E-03** | **2.91 (1.71 - 4.97)** | **1.60E-04** | Nervous system cancers - brain |
| NEO051<sup>5</sup> | **1.94 (1.20 - 3.13)** | **1.13E-02** | **2.92 (1.97 - 4.33)** | **2.61E-07** | Endocrine system cancers - pancreas |
| NEO067 |  | N/A | **8.07 (2.84 - 22.95)** | **1.60E-04** | Mesothelioma |
| NEO068<sup>5</sup> | **4.93 (1.82 - 13.36)** | **3.27E-03** | **3.90 (1.51 - 10.10)** | **6.10E-03** | Myelodysplastic syndrome (MDS) |
| NEO071<sup>5</sup> | **11.36 (6.49 - 19.86)** | **2.91E-16** | **12.02 (7.18 - 20.12)** | **6.49E-20** | Malignant neoplasm, unspecified |
| NVS004 | **6.74 (4.53 - 10.04)** | **2.31E-19** | **6.46 (4.46 - 9.37)** | **3.06E-21** | Parkinson`s disease |
| NVS005 | **7.32 (2.72 - 19.66)** | **2.26E-04** | **7.56 (3.00 - 19.10)** | **3.63E-05** | Multiple sclerosis |
| NVS006 | **4.10 (1.79 - 9.38)** | **1.84E-03** | **4.72 (2.11 - 10.53)** | **2.41E-04** | Other specified hereditary and degenerative nervous system conditions |
| NVS011 | **3.47 (2.49 - 4.82)** | **1.79E-12** | **4.17 (3.09 - 5.61)** | **8.08E-20** | Neurocognitive disorders |
| RSP002 | **4.61 (2.52 - 8.44)** | **2.50E-06** | **5.98 (3.54 - 10.11)** | **1.55E-10** | Pneumonia (except that caused by tuberculosis) |
| RSP008 | **2.20 (1.55 - 3.12)** | **2.87E-05** | **2.65 (1.93 - 3.66)** | **7.90E-09** | Chronic obstructive pulmonary disease and bronchiectasis |
| RSP016 | 0.35 (0.10 - 1.31) | 1.42E-01 | 0.66 (0.23 - 1.89) | 4.37E-01 | Other specified and unspecified lower respiratory disease |

| Outcome | Post-acute Mortality<sup>1</sup> |  | Overall Mortality<sup>2</sup> |  | CCSR Category Description |
| --- | --- | --- | --- | --- | --- |
|  | HR (95 % CI) | p.adj<sup>3</sup> | HR (95 % CI) | p.adj<sup>3</sup> |  |
| CIR003 |  | N/A<sup>4</sup> | 1.81 (0.71 - 4.60) | 4.44E-01 | Nonrheumatic and unspecified valve disorders |
| CIR004 |  | N/A | 2.41 (0.98 - 5.89) | 2.11E-01 | Endocarditis and endocardial disease |
| CIR005 |  | N/A | 0.60 (0.23 - 1.61) | 5.36E-01 | Myocarditis and cardiomyopathy |
| CIR008 | 1.00 (0.41 - 2.45) | 9.96E-01 | 1.19 (0.54 - 2.62) | 7.15E-01 | Hypertension with complications and secondary hypertension |
| CIR009 | 0.48 (0.17 - 1.32) | 4.49E-01 | 0.71 (0.48 - 1.06) | 2.74E-01 | Acute myocardial infarction |
| CIR011 | 0.73 (0.45 - 1.18) | 4.49E-01 | 1.03 (0.69 - 1.53) | 9.15E-01 | Coronary atherosclerosis and other heart disease |
| CIR019 |  | N/A | 1.29 (0.52 - 3.21) | 7.15E-01 | Heart failure |
| CIR021 | 1.22 (0.73 - 2.05) | 5.85E-01 | 1.37 (0.85 - 2.21) | 4.44E-01 | Acute hemorrhagic cerebrovascular disease |
| CIR026 | 2.19 (0.85 - 5.65) | 4.16E-01 | 2.86 (1.27 - 6.41) | 5.72E-02 | Peripheral and visceral vascular disease |
| DIG017 |  | N/A | 2.56 (0.89 - 7.36) | 2.50E-01 | Biliary tract disease |
| DIG025 | 2.35 (0.82 - 6.80) | 4.16E-01 | 3.16 (1.31 - 7.65) | 5.72E-02 | Other specified and unspecified gastrointestinal disorders |
| EXT020 | **4.04 (1.80 - 9.06)** | **7.80E-03** | **3.49 (1.63 - 7.44)** | **1.91E-02** | External cause codes: intent of injury, accidental/unintentional |
| EXT029 | **3.44 (1.56 - 7.59)** | **1.58E-02** | **2.98 (1.42 - 6.26)** | **3.10E-02** | External cause codes: subsequent encounter |
| INF003 | 1.96 (0.74 - 5.17) | 4.49E-01 | 1.60 (0.62 - 4.12) | 5.36E-01 | Bacterial infections |
| NEO012 | 0.34 (0.06 - 1.80) | 4.49E-01 | 0.41 (0.20 - 0.86) | 8.08E-02 | Gastrointestinal cancers - esophagus |
| NEO013 |  | N/A | 0.96 (0.42 - 2.17) | 9.15E-01 | Gastrointestinal cancers - stomach |
| NEO015 | 0.78 (0.52 - 1.19) | 5.03E-01 | 0.91 (0.63 - 1.32) | 7.15E-01 | Gastrointestinal cancers - colorectal |
| NEO017 | 0.97 (0.37 - 2.54) | 9.96E-01 | 0.80 (0.31 - 2.08) | 7.15E-01 | Gastrointestinal cancers - liver |
| NEO025 | 1.15 (0.50 - 2.67) | 8.54E-01 | 1.23 (0.56 - 2.71) | 7.15E-01 | Skin cancers - melanoma |
| NEO043 |  | N/A | 1.50 (0.71 - 3.17) | 5.20E-01 | Urinary system cancers - bladder |
| NEO048 | 0.73 (0.33 - 1.62) | 5.85E-01 | 0.84 (0.41 - 1.72) | 7.15E-01 | Nervous system cancers - brain |
| NEO051 | 0.57 (0.34 - 0.94) | 1.55E-01 | 0.74 (0.48 - 1.15) | 4.44E-01 | Endocrine system cancers - pancreas |
| NEO071 | 1.23 (0.67 - 2.26) | 6.24E-01 | 1.38 (0.79 - 2.42) | 4.95E-01 | Malignant neoplasm, unspecified |
| NVS004 | 1.25 (0.79 - 1.99) | 5.30E-01 | 1.22 (0.79 - 1.87) | 5.78E-01 | Parkinson`s disease |
| NVS006 | 0.99 (0.36 - 2.73) | 9.96E-01 | 1.44 (0.59 - 3.50) | 6.21E-01 | Other specified hereditary and degenerative nervous system conditions |
| NVS011 | **1.86 (1.37 - 2.50)** | **1.21E-03** | **2.07 (1.56 - 2.73)** | **1.06E-05** | Neurocognitive disorders |
| RSP002 | 0.63 (0.27 - 1.48) | 5.30E-01 | 0.77 (0.36 - 1.62) | 6.89E-01 | Pneumonia (except that caused by tuberculosis) |
| RSP008 | 0.76 (0.41 - 1.39) | 5.39E-01 | 1.11 (0.69 - 1.80) | 7.15E-01 | Chronic obstructive pulmonary disease and bronchiectasis |
| RSP016 | 1.42 (0.70 - 2.88) | 5.30E-01 | 1.52 (0.79 - 2.93) | 4.44E-01 | Other specified and unspecified lower respiratory disease |

HR: hazard ratio; 95% CI represents the lower and upper 95% confidence interval of HR; p.adj<sup>3</sup>: FDR-adjusted p-value.
CCSR: Clinical Classifications Software Refined, a method to classify disease categories.
1. Post-acute Mortality measures the risk of death by setting the follow-up start date as 30 days after the date of first positive SARS-CoV-2 test. 2. Overall Mortality measures the risk of death by setting the follow-up start date as the date of first positive SARS-CoV-2 test. 3. FDR correction was separately performed for each COVID-19 exposure cohort; Statistically significant associations (p.adj < 0.05) were labelled in bold face. 4. N/A indicates that the analysis for target outcome cannot proceed due to insufficient number (<5) of events in either the COVID-19 exposure or reference cohort. 5. Elevated neoplasm mortality, especially among hospitalized patients without prior cancer history, should be interpreted with caution, as this finding is more likely attributable to detection bias (unmasking of pre-existing undiagnosed malignancies) rather than representing a direct post-infection sequela of COVID-19. See Discussion for detailed interpretation.

The proportion of significant associations was 42.9% (12/28), and 10.7% (3/28) for post-acute death outcomes *with*, and *without* a prior history, respectively (Fig. S3). Corresponding proportions for overall mortality were 54.5% (18/33) and 18.2% (6/33). Detailed results are elaborated in Supplementary_text and Table_S4.

### Mortality risks of individual outcomes for hospitalized COVID-19

Compared to the reference population, hospitalized COVID-19 significantly elevated post-acute mortality for 73.0% (27/37) of outcomes encompassing multiple systems (Table_2b); the proportion was higher in the overall-mortality analysis (36/42, 85.7%). Among post-acute deaths *with* and *without* a pre-existing diagnosis, 91.7% (22/24) and 71.4% (10/14) of disorders exhibited greater mortality risk (Table_S4). However, we cautioned that mortality without a prior history, particularly for neoplasms, might reflect detection bias (i.e., unmasking of pre-existing subclinical malignancies during hospitalization) rather than truly new-onset diseases. These associations should not be interpreted as COVID-19-driven sequelae and are mechanistically distinct from other outcomes such as circulatory or respiratory mortality. For overall mortality, the proportions of conditions with increased risk were 87.1% (27/31) for those with a prior history and 90.5% (19/21) for those without (Table_S4; see Supplementary_text).

### Mortality risks of individual outcomes for non-hospitalized COVID-19

Two external-cause outcomes and neurocognitive disorders were significantly associated with increased post-acute (3/22) and overall (3/29) mortality for non-hospitalized COVID-19 (Table_2c). Similar patterns were observed for deaths *with* prior history (post-acute: 3/9; overall: 4/15; Table_S4).

For deaths *without* pre-existing diagnoses, only neurocognitive disorders remained significant for post-acute mortality. For overall mortality, the list expanded to also include peripheral/visceral vascular disease, biliary tract disorders, and other gastrointestinal disorders (Table_S4; see Supplementary_text).

### Subgroup analysis

Significant heterogeneity in post-acute mortality was observed across subgroups (Table_3, Fig. 2). All-cause mortality risk was significantly greater in individuals with advanced age (>65 years), atrial fibrillation (AF), chronic kidney disease (CKD), or hypertension (HTN) than in their risk-factor-absent counterparts.

**Figure 2.**
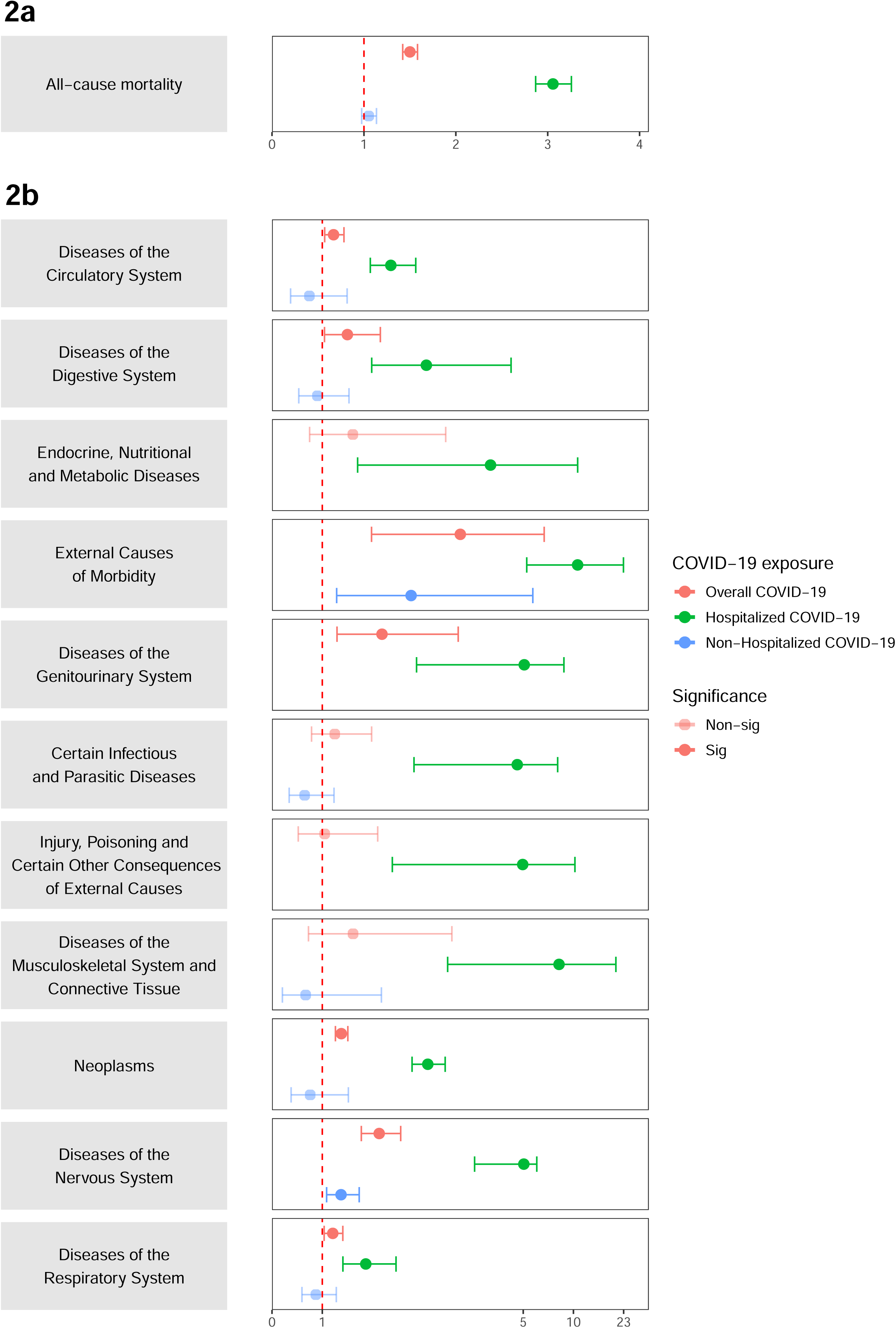
Significant Differences in Associations between Post-acute Mortality Risks in Organ Systems and Overall COVID-19 Across Subgroups. **Legend:** Comparisons showing significant differences (pval.adj_RHR_ < 0.05) in Hazard ratios (HR) for mortality with (risk factor present) and without (risk factor absent) specific characteristics are presented. The red dashed line represents the line of no effect (hazard ratio (HR) = 1). X-axis indicates the HR of post-acute mortality after COVID-19 infection. Y-axis indicates each CCSR disease category. Red and blue color respectively represent association results in the risk factor present and absent subgroups. Solid bars indicate significance, and transparent bars indicate non-significant associations for each subgroup. We only present the results if the number of events ≥5 for COVID-19 and uninfected individuals in both subgroups. Advanced age represents >65 years old, and Middle age refers to 50-65 years old

**Table 3.** Significant Differences in Associations between Post-Acute Mortality Risks in Organ Systems and Overall COVID-19 Across Subgroups.

| Outcome | HR(CI)<sub>present</sub> | p.adj<sub>present</sub> | Sig<sub>present</sub> | HR(CI)<sub>absent</sub> | p.adj<sub>absent</sub> | Sig<sub>absent</sub> | RHR | p.adj<sub>RHR</sub>* | Comparison | Organ System Description |
| --- | --- | --- | --- | --- | --- | --- | --- | --- | --- | --- |
| All-cause mortality | 1.61<br>(1.52 - 1.70) | 3.84E-59 | *** | 1.15<br>(0.98 - 1.36) | 1.94E-01 |  | **1.40** | **1.18E-03** | Advanced age<sup>1</sup> vs.<br>Middle age<sup>2</sup> |  |
| All-cause mortality | 1.69<br>(1.50 - 1.89) | 2.69E-18 | *** | 1.43<br>(1.34 - 1.52) | 2.35E-29 | *** | **1.18** | **3.55E-02** | AF vs. AF-free |  |
| All-cause mortality | 1.80<br>(1.63 - 1.97) | 4.86E-33 | *** | 1.34<br>(1.26 - 1.43) | 2.13E-17 | *** | **1.34** | **6.31E-06** | CKD vs. CKD-free |  |
| All-cause mortality | 1.59<br>(1.48 - 1.69) | 8.92E-42 | *** | 1.31<br>(1.19 - 1.44) | 2.87E-07 | *** | **1.21** | **7.52E-03** | HTN vs. HTN-free |  |
| CIR | 1.37<br>(1.16 - 1.61) | 2.39E-04 | *** | 0.51<br>(0.28 - 0.92) | 1.50E-01 |  | **2.67** | **4.61E-03** | Advanced age<sup>1</sup> vs.<br>Middle age<sup>2</sup> | Diseases of the circulatory system |
| NVS | 2.49<br>(2.04 - 3.03) | 8.44E-19 | *** | 1.12<br>(0.67 - 1.87) | 6.73E-01 |  | **2.23** | **2.33E-02** | AF vs. AF-free | Diseases of the nervous system |
| RSP | 1.89<br>(1.26 - 2.85) | 5.29E-03 | ** | 0.94<br>(0.70 - 1.25) | 6.73E-01 |  | **2.02** | **2.33E-02** | AF vs. AF-free | Diseases of the respiratory system |

1. Advanced age: >65 years old.
2. Middle age: 50-65 years old.
HR(CI)<sub>present</sub>: Hazard ratio and 95% confidence interval for the risk factor present subgroup with the examined characteristics; p.adj<sub>present</sub>: FDR-adjusted p-value for the risk factor present subgroup;
HR(CI)<sub>absent</sub>: Hazard ratio and 95% confidence interval for the risk factor absent subgroup without the examined characteristics; p.adj<sub>absent</sub>: FDR-adjusted p-value for the risk factor absent subgroup;
Sig<sub>present</sub> and Sig<sub>absent</sub>: \* indicates an FDR-adjusted p-value between 0.01 and 0.05, \*\* indicates between 0.001 and 0.01, and \*\*\* indicates a value smaller than 0.001 for the risk factor present and absent subgroups, respectively.
RHR: Ratio of hazard ratios for comparisons between the risk factor present and absent subgroups.
p.adj<sub>RHR</sub>\*: FDR-adjusted p-value for comparisons between the risk factor present and absent subgroups, where a p.adj<sub>RHR</sub> < 0.05 indicates significant differences.
Only results with significant differences (p.adj<sub>RHR</sub> < 0.05) are shown.

Older adults (>65 years) demonstrated higher post-acute circulatory mortality than middle-aged participants. AF patients experienced higher post-acute mortality from nervous and respiratory diseases than AF-free individuals.

No significant subgroup differences were found for individual CCSR outcomes after FDR correction. Subgroup analyses for overall mortality yielded largely consistent findings. A nominally decreased post-acute circulatory mortality was observed among middle-aged participants (HR: 0.51, 0.28-0.92); however, this result did not retain statistical significance after FDR correction (adjusted p =0.15). Given the large number of subgroup comparisons performed, this likely represents a chance finding. This association was also absent when the acute phase was included (0.77, 0.48-1.24). We therefore refrain from drawing substantive conclusions from this observation. Detailed results are provided in Tables_S5–S16.

### Additional analyses

- **Undocumented infections:** both QBA and the negative-test restricted reference analyses yielded generally higher HRs than the primary analysis(Table_S17-S18), indicating that primary estimates quantify the mortality risk of clinically apparent COVID-19 relative to the general UK population rather than relative to truly uninfected individuals. Undocumented infections in the reference cohort likely attenuated effect estimates, though the overall pattern of significant associations was preserved.
- **Vaccination status**: Elevated all-cause mortality was observed in both vaccinated and unvaccinated cases, with greater risk estimates in the unvaccinated group but no significant interaction with vaccination detected (Table_S19-S20).
- **Reinfection:** Estimates were similar when follow-up was extended beyond reinfection (Table_S21), suggesting the reinfection-censoring strategy had minimal impact on the results. Stratified analyses by infection number showed higher risks of all-cause, neoplastic, neurological and respiratory mortality among reinfected individuals compared to those with a single infection (Table_S22). Further details are presented in Supplementary_text.

## Discussion

This UKBB cohort study examined the risks of all-cause and cause-specific mortality across 12 organ systems and 47 individual disease categories for up to 35 months following COVID-19. SARS-CoV-2 infection was associated with increased post-acute mortality due to circulatory, digestive, genitourinary, neurological, and respiratory disorders, as well as external causes and neoplasms. The risk was particularly pronounced in individuals who had been hospitalized for COVID-19. Overall-mortality analyses (including both acute and post-acute phases) showed broader significance with larger effect estimates. Furthermore, analyses stratified by patient history and comorbidities revealed that these mortality risks are significantly influenced by pre-existing conditions and demographic factors.

An important interpretive consideration is that the reference cohort, defined by the absence of documented SARS-CoV-2 infection, likely included individuals with unrecorded infections given the high cumulative UK prevalence during the study period [17]. Our findings therefore reflect the mortality risk of clinically apparent COVID-19 relative to the general UK population during the pandemic, rather than relative to strictly uninfected individuals. Both QBA and negative-test restricted reference analyses confirmed that this contamination biases estimates toward the null, indicating that true infection-attributable mortality risks are likely larger than reported.

Previous studies [7,8] reported excess mortality after the emergence of SARS-CoV-2, most prevalently involving respiratory, cardiovascular, neurological disorders, and cancers. However, since these estimates were based on population-level aggregates without adjustment for individual confounders, they can be sensitive to the choice of pre-pandemic reference period and may reflect healthcare overloads or socioeconomic disruptions rather than COVID-19’s unique effects [18]. In contrast, this study directly assessed mortality following SARS-CoV-2 infection using individual-level data and Cox regression, with a primary focus on the post-acute phase (beyond 30 days). We reported increased mortality for previously identified conditions, but also uncovered new associations with other disorders such as digestive, genitourinary diseases, and external injuries.

Consistent with existing literature [5], we observed elevated all-cause mortality in COVID-19 survivors, with the highest HR among those requiring hospitalization. Post-acute all-cause mortality did not reach significance in the non-hospitalized cohort, but it was significant when considering overall mortality from infection onset. Cause-specific analyses further showed that hospitalized COVID-19 individuals experienced increased post-acute and overall mortality across a broader spectrum of organ systems and CCSR disorders than the non-hospitalized group, demonstrating a clear linkage between initial COVID-19 severity and subsequent mortality burden.

Our work addresses a critical gap in prior research, which primarily investigated symptoms or hospitalizations (but not mortality) as COVID-19 sequelae [5,6]. While Mendelian randomization studies support potential causal associations between hospitalized COVID-19 and post-COVID-19 syndromes across multiple systems [19–21], this method is less suited to examine post-COVID-19 mortality due to methodological constraints, for example the difficulty in obtaining genetic instruments for mortality outcomes and the inability to capture the full temporal evolution of post-infection risk. Many previous epidemiology or MR studies [22,23] are also limited to single systems or a restricted number of pre-specified conditions. In contrast, our study provides a systematic and comprehensive assessment of post-infection mortality using CCSR classification, avoiding any arbitrary exclusions or preferential selections and thus enabling the identification of a wider array of high-risk outcomes. Additionally, subgroup analyses were conducted to explore differences in post-acute/overall effects across subpopulations.

The mechanisms driving the excess mortalities are likely multifactorial. The observed increase in respiratory-related post-acute mortality aligns with previous studies suggesting non-COVID pneumonia as a common COVID-19 sequela [24]. The elevated post-acute cardiovascular mortality may be attributable to direct infiltration of SARS-CoV-2, autoimmune dysregulation, and inflammatory response within cardiac tissues [25]. A recent study [10] also revealed increased 12-month risks of all-cause, respiratory, and cardiovascular mortality in hospitalized COVID-19 patients, though respiratory/cardiovascular conditions accounted for only 20.5% of deaths. This corroborates our observation that SARS-CoV-2’s impact extends beyond these two systems. For instance, we found increased post-acute mortality for neurological disorders, consistent with research demonstrating a persistent two-year post-infection risk of neurocognitive deficits [9].

We also identified increased mortality from conditions less frequently associated with COVID-19. For example, we observed elevated post-acute genitourinary mortality, which could be mediated through coronavirus binding to ACE2 receptors in the urinary tract, inducing cellular damage and increasing susceptibility to infection [26]. Besides, renal complications were reported among COVID-19 survivors [27], potentially resulting from direct viral invasion, persistent tubular/microvascular injury from systemic inflammation and coagulopathy, and podocyte damage. Our finding of elevated post-acute digestive mortality aligns with emerging evidence of viral RNA persistence in gut tissues and sustained systemic inflammation, contributing to gastrointestinal/hepatobiliary manifestations of long COVID [28]. Furthermore, the increased post-acute mortality from external causes, primarily accidental injuries, highlights significant secondary consequences of COVID-19. Post-infection muscle weakness, fatigue, and cognitive dysfunction may impair physical coordination and risk perception [24]. COVID-19 or its clinical treatment (e.g., glucocorticoids) may also accelerate bone loss and osteoporosis progression, raising the likelihood of fractures and fatal injuries [29].

Post-acute mortality from neoplasms was elevated; however, interpretation differs by prior cancer status. Among patients *with* a prior cancer history, the elevated mortality may represent a true sequela, mediated by COVID-19-induced immune dysregulation that exacerbates pre-existing malignancies. Viral infection may trigger excessive inflammatory cytokine release, fostering a pro-tumorigenic environment [30]. Moreover, SARS-CoV-2 may deplete critical cancer-fighting immune cells such as CD4+/CD8+ T cells, impairing immune surveillance and worsening prognosis in cancer patients following infection.

In contrast, elevated cancer mortality in patients *without* prior diagnoses, observed exclusively in hospitalized cases, is more plausibly attributable to detection bias. Intensive clinical evaluation during hospitalization may have incidentally uncovered pre-existing but undiagnosed malignancies, or the physiological stress of severe infection may have accelerated the clinical presentation of subclinical cancers [31]. Given the biological implausibility of de novo carcinogenesis within the study timeframe, these findings should be distinguished from deaths attributable to post-acute sequelae of COVID-19.

While mortality risks appeared less pronounced in individuals without prior history of the examined outcomes, several non-neoplastic post-acute associations remained significant and are more biologically plausible as direct consequences of SARS-CoV-2-mediated pathology. For example, peripheral/visceral vascular disease may arise from SARS-CoV-2 invasion of vascular epithelial cells via ACE2 receptors, triggering intravascular thrombotic events and endothelial dysfunction [32]. Unlike the neoplasm findings discussed above, these associations may reflect established post-infection mechanisms and support a genuine sequela interpretation.

Comparing the post-acute and overall mortality analyses provided key insights. Compared with the post-acute findings, overall-mortality analyses yielded broader significance and generally larger estimates across all COVID-19 cohorts. Several associations (e.g., heart failure and bronchiectasis) were substantially attenuated in the post-acute analysis, suggesting these mortality risks are primarily concentrated within the first 30 days after infection. Conversely, the persistence of many associations after the 30-day landmark indicates that SARS-CoV-2’s impact on mortality extends well beyond the acute period.

Our subgroup analysis indicated that older adults with chronic conditions (AF, CKD, or HTN) were more vulnerable to deleterious consequences of SARS-CoV-2 with elevated all-cause and circulatory mortality. Moreover, AF patients demonstrated increased post-acute/overall mortality for both neurological and respiratory diseases, potentially suggesting a synergistic prothrombotic effect, as both AF and COVID-19 may induce thromboembolic complications like ischemic stroke and pulmonary embolism, contributing to the corresponding organ dysfunction and observed mortality patterns [33]. Importantly, although at a lower magnitude than their risk-factor-present counterparts, elevated post-acute/overall all-cause mortality was observed in certain risk-factor-absent groups, highlighting the multifactorial nature of post-COVID-19 mortality.

Although unvaccinated cases showed a broader risk spectrum than vaccinated individuals, directionally consistent with evidence suggesting that COVID-19 vaccination may attenuate post-acute sequelae [34], our data cannot confirm this given the lack of statistically significant interaction. Mortality risks were also higher among reinfected individuals, though this warrants cautious interpretation given limited statistical power due to sparse events, and the possibility that reinfected groups represent a more immunologically vulnerable population. As our study was not designed to evaluate the effects of vaccination or reinfection on post-COVID-19 mortality, further investigations are needed.

### Strengths and limitations

This study possesses unique strengths. First, it leveraged the breadth and depth of UKBB electronic health record system to establish a large cohort with sufficient sample size and follow-up. Instead of focusing on symptoms/hospitalizations, we analyzed mortality as the most severe post-infection consequence. Secondly, our primary 30-day landmark design helped distinguish post-acute effects from acute-phase mortality, with overall-mortality analyses provided for comparison. This dual analytical approach enables identification of both immediate and delayed mortality risks. Also, the contribution of SARS-CoV-2 to deaths was assessed via regression using individual-level data rather than excess-mortality estimates, and causes of post-COVID mortality were systematically categorized into a comprehensive range of 12 organ systems and 47 individual disorders using CCSR. In addition, analyses stratified by infection severity, prior history, and demographic/comorbidity factors further revealed heterogeneity in post-acute mortality risks.

Our work has several limitations. First, the UKBB cohort comprises primarily older, white, healthier individuals with higher educational attainment [35], which may limit generalizability of the observed mortality to other populations. However, the pathophysiological mechanisms underlying post-acute sequelae of COVID-19 are likely broadly applicable across diverse populations. Therefore, the fundamental insights derived from our findings remain highly relevant beyond the study cohort. Second, as an observational study, causality cannot be established between COVID-19 and post-acute mortality risks. While we adjusted for a comprehensive set of covariates, unmeasured confounding cannot be completely excluded. Thirdly, our results represent average mortality risks observed during the study period (31-Jan-2020 to 19-Dec-2022), and longer-term effects beyond this timeframe remain uncertain. Fourth, cause-of-death classification based on ICD-10 codes may be subject to misclassification, and the small number of events for certain outcomes may result in limited statistical power. Fifth, given the high cumulative UK COVID-19 prevalence during the study period [17], the reference cohort likely included individuals with undocumented infections. As discussed above, this shifts the comparative baseline such that our estimates reflect clinically apparent COVID-19 versus the general population rather than strictly uninfected individuals. Both QBA and negative-test restricted reference-group analyses confirmed that true infection-attributable risks are likely larger, while the overall pattern of significant associations remained unchanged. Lastly, due to restricted data availability during analysis, UKBB vaccination records including booster doses were incomplete. Hence, vaccination status was treated only as a binary covariate and its effects on post-COVID mortality could not be examined in detail.

### Clinical implications

This study demonstrates that clinically apparent COVID-19 is associated with elevated post-acute all-cause and cause-specific mortality across multiple organ systems relative to the general population without known infection, with hospitalized infections showing significantly increased risks across a broader spectrum of outcomes. However, elevated cancer mortality among patients without prior diagnoses may reflect detection bias rather than direct post-infection pathology, and should be interpreted distinctly from other sequelae-driven mortality. Overall-mortality analyses further supported these patterns with generally larger effect estimates. This underscores the critical need for preventive strategies to mitigate COVID-19 severity through early treatment. Notably, even non-hospitalized infection is linked to increased post-acute deaths from neurological disorders and external-cause injuries, and overall-mortality analysis suggests modestly elevated all-cause mortality. Given the huge number of infections worldwide, and that our estimates might be conservative relative to truly uninfected individuals, the absolute mortality burden imposed by COVID-19 is likely substantial.

In the post-pandemic era, public health policies should prioritize the management of post-COVID-19 complications by allocating healthcare resources for long-term follow-up and risk assessment of COVID-19 survivors, especially older adults and those with pre-existing high-risk comorbidities. Future research is crucial to investigate the underlying mechanisms, identify effective interventions to ameliorate the mortality burden, and examine the long-term trajectory of mortality risks tied to post-COVID-19 outcomes.

## Statements and Declarations

### Ethics Approval Statement

The UK Biobank study has received ethical approval from the NHS National Research Ethics Service North West (16/NW/0274). Individual consents were obtained by the UK Biobank. The current study was conducted under the project number 28732. Only de-identified data was accessed, and no attempts were made to identify any individual participants in this study.

### Consent to publish

Not applicable.

### Availability of data and materials

UK Biobank (UKBB) data was used for the current study. Access to UKBB data is restricted to researchers who have made prior applications. Detailed instructions on how to apply for access to the database can be found on the UK Biobank homepage (https://www.ukbiobank.ac.uk/enable-your-research/apply-for-access). All supplementary tables are available at the journal’s website and at https://drive.google.com/drive/folders/1ay_8h84m-dmGpHz-Bp_5IIpzEZwWxE6Z?usp=drive_link.

### Authors’ contributions

**RZ:** Writing – review & editing, Writing – original draft, Visualization, Validation, Software, Resources, Methodology, Formal analysis. **YX:** Writing – review & editing, Visualization, Validation, Software, Formal analysis. **JQ:** Writing – review & editing, Visualization, Validation, Formal analysis. **HCS:** Writing – review & editing, Writing – original draft, Validation, Supervision, Project administration, Funding acquisition, Data curation, Conceptualization. All authors read and approved the final manuscript.

## Data Availability

UK Biobank (UKBB) data was used for the current study. Access to UKBB data is restricted to researchers who have made prior applications. Detailed instructions on how to apply for access to the database can be found on the UK Biobank homepage.

https://drive.google.com/drive/folders/1ay_8h84m-dmGpHz-Bp_5IIpzEZwWxE6Z?usp=drive_link

## Acknowledgements

We gratefully acknowledge support from the National Natural Science Foundation of China (Grant-81971706), the Lo-Kwee-Seong Biomedical Research Fund, and the Joint Laboratory of Bioresources and Molecular Research of Common Diseases of the Kunming Institute of Zoology and The Chinese University of Hong Kong, China. The authors have used Gemini-2.0-Flash to correct grammatical errors and improve the overall readability, and the original draft was written by the authors without the help of Gemini. Special thanks to Prof. Pak Sham for data access.

## Figure titles and Legends

**Supplementary Figure 1.** An Overview of Analytic Workflow. **Legend**: **Cohort establishment:** UKBB participants were classified into overall COVID-19 exposure and reference cohorts. This overall exposure cohort was further divided into hospitalized and non-hospitalized exposure cohorts. For our primary analysis, samples who died within the 30-days acute period after COVID-19 infection were excluded for all the cohorts. **Outcome definition:** The primary (composite) outcomes were: (1) all-cause mortality and (2) cause-specific mortality grouped by 12 organ systems; Secondary (single) outcome included cause-specific mortality grouped by 47 individual CCSR disorders, which were further stratified by the presence or absence of a prior history of the same condition **Statistical analysis:** In our primary analysis, adjusted Cox regression models were employed to assess post-acute mortality risks of overall COVID-19 exposure compared to the reference. Additional analyses included (1) stratification of the exposure cohort into hospitalized and non-hospitalized cohorts; (2) stratification of single CCSR outcomes by prior history of the same condition; and (3) subgroup comparisons based on the presence or absence of specific risk factors. For further details, please refer to the Method sections in the main text

**Supplementary Figure 2.** Associations Between All-Cause and Cause-Specific Overall Mortality Risks in Different Organ Systems and Overall/Hospitalized/Non-hospitalized COVID-19. **Legend:** The vertical red dashed line represents the line of no effect (hazard ratio (HR) = 1). X-axis indicates the HR of overall mortality after COVID-19 infection. Y-axis indicates each COVID-19 disease severity. Confidence intervals are also shown in the figure. Solid bars indicate significance, and transparent bars indicate non-significant associations. We only present the results if the number of events ≥5 for both COVID-19 exposure and reference cohort

**Supplementary Figure 3.** Associations Between Cause-Specific Post-acute Mortality Risks in Single Disorders (Stratified by Prior History) and Overall COVID-19. **Legend:** The 1st, 2nd and 3rd column respectively represent the results for any death records, death with a prior history, and death without a prior history of the same condition. The red dashed line represents the line of no effect (hazard ratio (HR) = 1). X-axis indicates the HR of post-acute mortality after COVID-19 infection. Y-axis indicates each CCSR disease category. Solid bars indicate significance, and transparent bars indicate non-significant associations. We only present the results if the number of events ≥5 for both COVID-19 exposure and reference cohort. Single CCSR disorders are grouped by organ systems. CIR: circulatory system diseases; DIG: digestive system diseases; END: endocrine, nutritional and metabolic diseases; EXT: external causes of morbidity; GEN: genitourinary system diseases; INF: certain infectious and parasitic diseases; NEO: neoplasms; NVS: nervous system diseases; RSP: respiratory system diseases

**Supplementary Figure 4.** Associations Between Cause-Specific Overall Mortality Risks in Single Disorders (Stratified by Prior History) and Overall COVID-19. **Legend:** The 1st, 2nd and 3rd column respectively represent the results for any death records, death with a prior history, and death without a prior history of the same condition. The red dashed line represents the line of no effect (hazard ratio (HR) = 1). X-axis indicates the HR of overall mortality after COVID-19 infection. Y-axis indicates each CCSR disease category. Solid bars indicate significance, and transparent bars indicate non-significant associations. We only present the results if the number of events ≥ 5 for both COVID-19 exposure and reference cohort. Single CCSR disorders are grouped by organ systems. CIR: circulatory system diseases; DIG: digestive system diseases; END: endocrine, nutritional and metabolic diseases; EXT: external causes of morbidity; GEN: genitourinary system diseases; INF: certain infectious and parasitic diseases; NEO: neoplasms; NVS: nervous system diseases; RSP: respiratory system diseases

