## Supplementary figures and images for "Association of COVID-19 with risks of all-cause and cause-specific mortality post-infection: A UK Biobank cohort study"

### Supplementary Figure 1

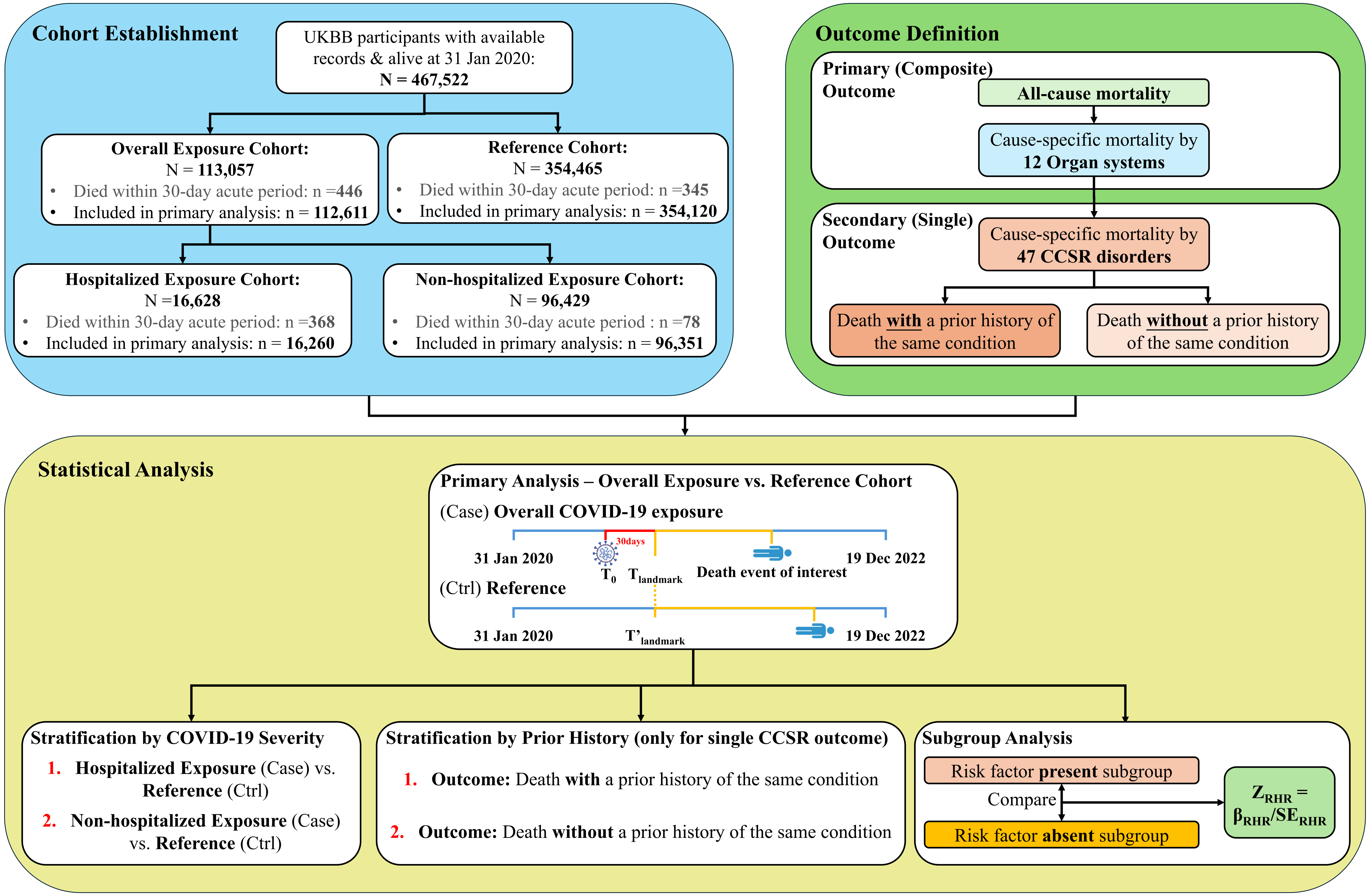

### Supplementary Figure 2

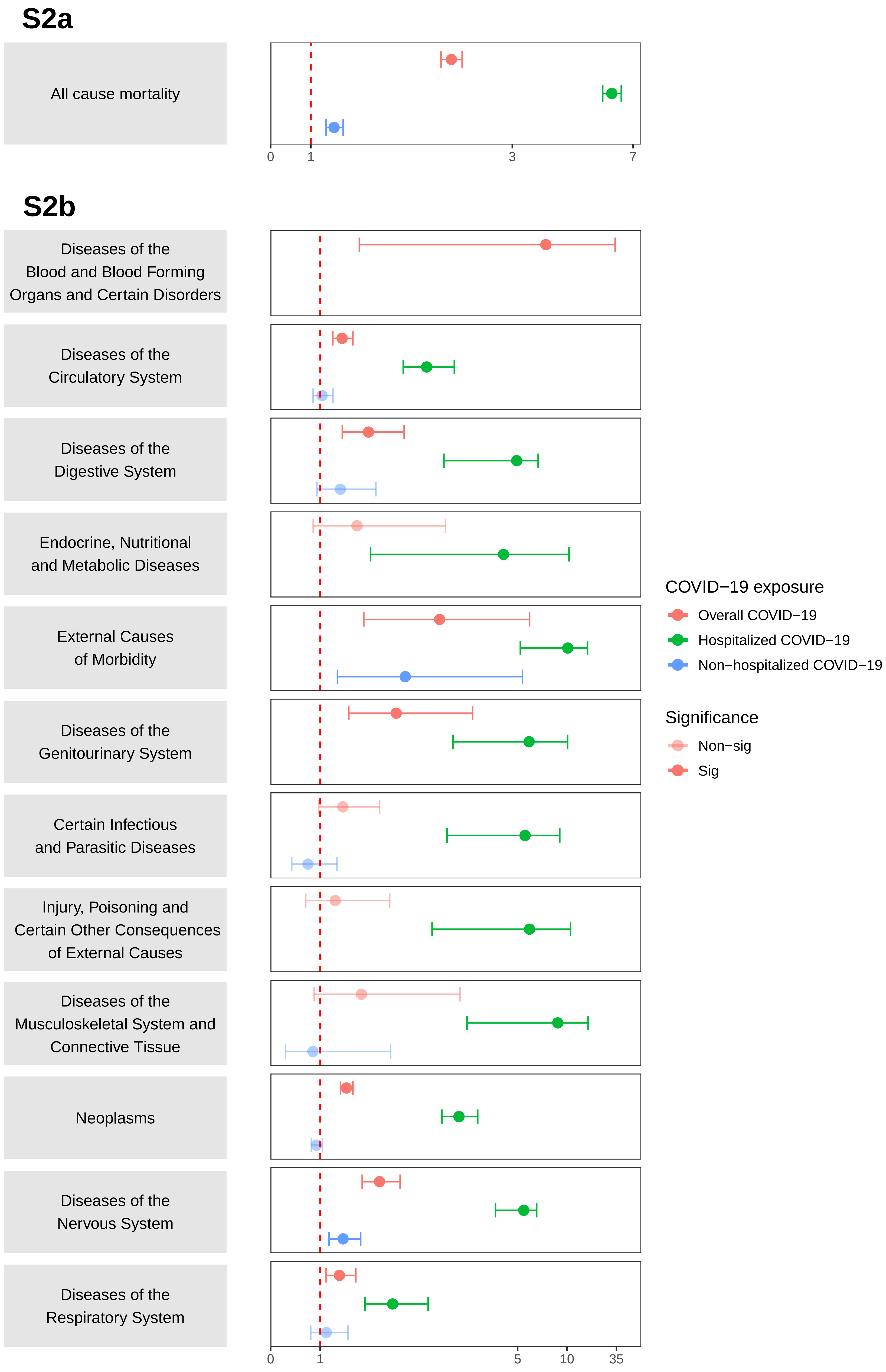

### Supplementary Figure 3

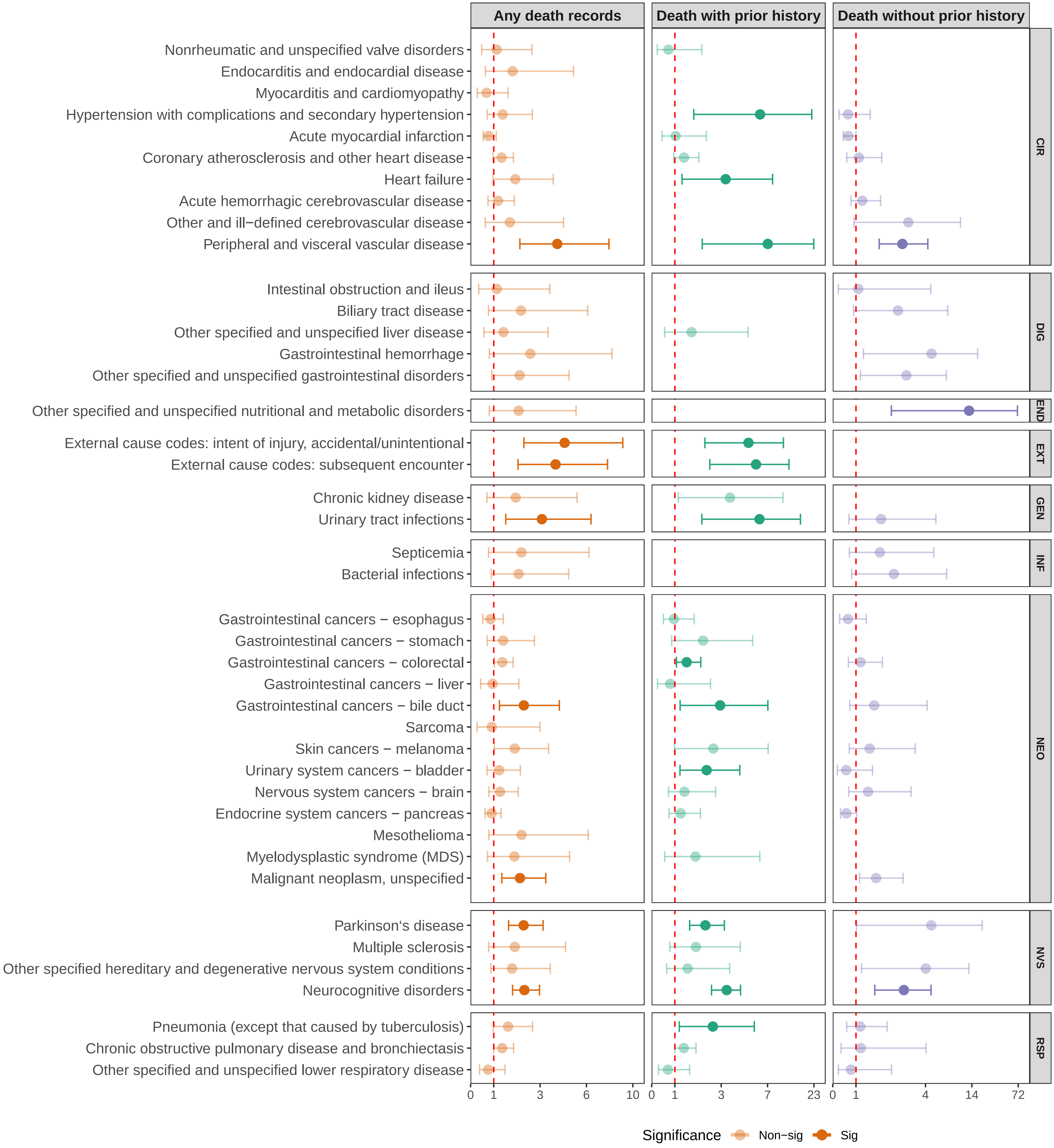

### Supplementary Figure 4

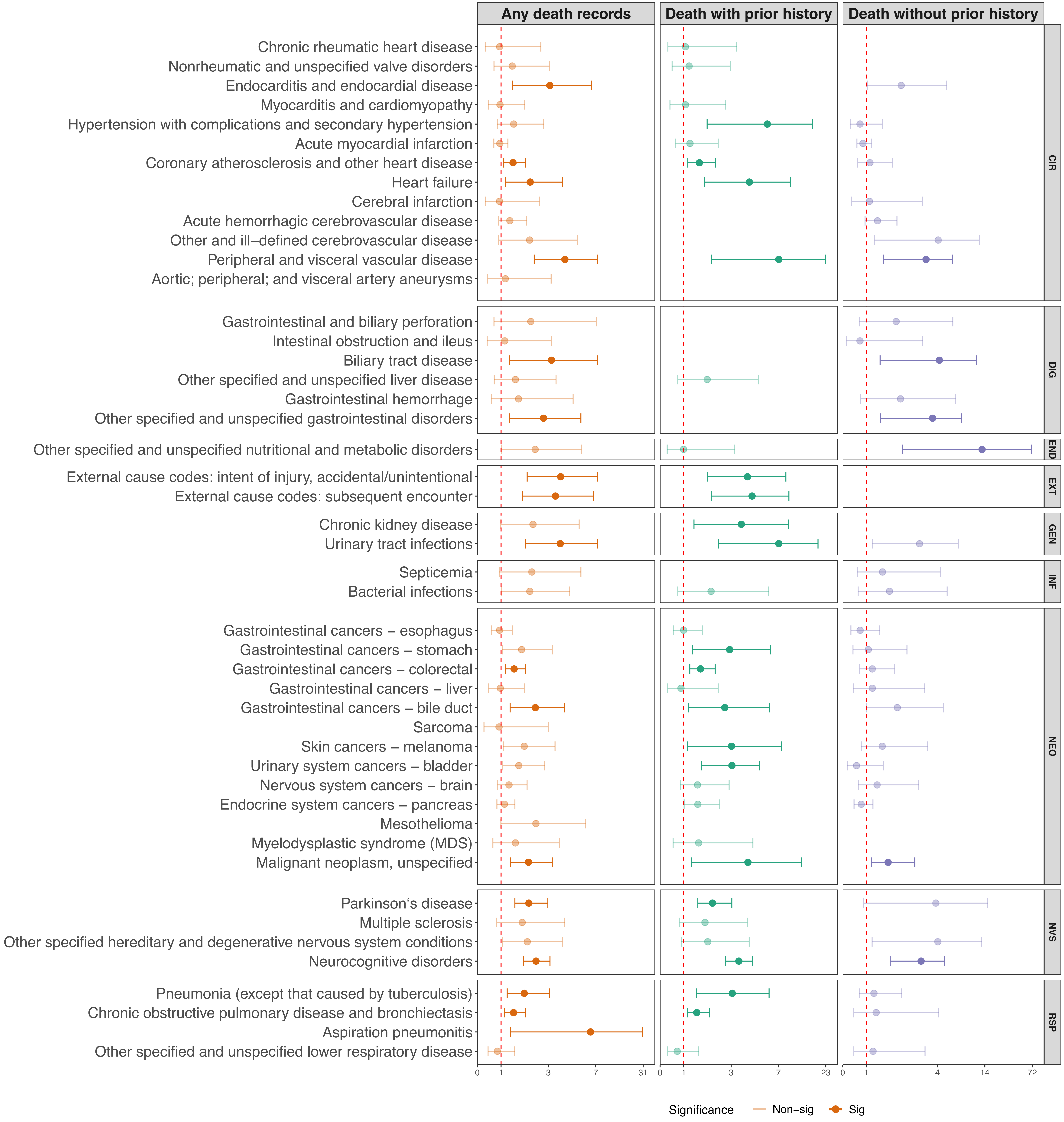
