## Supplementary Text for "Association of COVID-19 with risks of all-cause and cause-specific mortality post-infection: A UK Biobank cohort study"

**Supplementary Methods**	**2-9**

**Full results**	**10-19**

**References**	**20**

***Supplementary Methods***

**Data Sources**

Participants’ baseline demographic characteristics and clinical information were accessed from UKBB. As the primary study outcome, mortality records linked to UKBB participants were obtained from national death registries. This mortality data incorporates the corresponding date and causes of death. Primary causes of death were considered as outcomes to prevent confounding from secondary conditions that might obscure the true COVID-19-mortality relationship. Pre-existing (ICD-10 coded) comorbidities, were extracted from multiple sources within the UKBB, including baseline assessments, follow-up questionnaires, Hospital Episode Statistics (HES) data, and General Practitioner (GP) records. Exposure to SARS-CoV-2 infection was defined using COVID-19 testing data from the UKBB portal: specifically, any positive PCR test result or any hospital inpatient record associated with diagnosis code U07.1 during the study follow-up period. Any COVID-19 tests, clinical events, or deaths occurring after the study end date were excluded from the analysis.

**Outcomes**

Given that other contributory causes of death in UKBB mortality records might reflect complex comorbidities not directly attributable to COVID-19 exposure, our analysis focused solely on the primary cause of death. These ICD-10 primary cause codes were subsequently categorized into clinically relevant groups based on the Healthcare Cost and Utilization Project (HCUP) Clinical Classifications Software Refined (CCSR) for ICD-10-CM, version 2022.1 [1].

The primary study outcomes were post-acute mortality, defined as deaths occurring ≥31 days after SARS-CoV-2 infection, which included: (1) all-cause mortality and (2) cause-specific mortality, grouped by HCUP CCSR body system categories. For each body system, mortality was defined by the presence of at least one contributing sequela within that system. Secondary outcomes included post-acute mortality due to individual CCSR-defined disease categories. As a complementary analysis, we also evaluated overall mortality across the entire follow-up period (i.e., including deaths within the acute phase of days 0-30) using the same outcome definitions.

To ensure adequate statistical power, analyses were restricted to outcomes with ≥5 death events in both the COVID-19 exposure and reference cohorts. This filtering step yielded up to 12 composite (organ-system) outcomes and 47 single (CCSR-defined disease) categories for inclusion in the analysis (Table_S1). As the minimum event threshold was applied separately in the post-acute and overall mortality analyses, the exact number of eligible outcomes may differ slightly between these two analytic frameworks. In addition, each single CCSR outcome was stratified by pre-existing diagnosis of the same condition, based on records before *T_0_*, creating subgroups of deaths with and without a relevant prior history. This stratification was conducted in both the post-acute and overall analyses.

**Covariates**

To adjust for baseline differences between the COVID-19 exposure and reference cohorts, a comprehensive set of covariates that may confound the associations between SARS-CoV-2 infection and post-infection outcomes were selected for statistical analysis [2]. These comprised: sociodemographic characteristics (age, sex, ethnicity, and the Townsend Deprivation Index, with higher values indicating greater area-level material deprivation); lifestyle and anthropometric measures (smoking status, Body Mass Index [BMI], and waist circumference); biochemical markers (lipid and glucose levels, haemoglobin A1c [HbA1c]); immunological factors (autoimmune diseases, immunodeficiency, prior immunosuppressant use); pre-existing medical conditions across various systems, including cardiovascular (atrial fibrillation [AF], coronary artery disease [CAD], hypertension [HTN], and stroke), respiratory (chronic obstructive pulmonary disease [COPD] and pneumonia), renal (chronic kidney disease [CKD]), metabolic (type 2 diabetes mellitus [T2DM]), neurological (dementia), and oncologic (cancer); COVID-19 vaccination status; and indicators of overall health status in the prior year (number of non-cancer illnesses, GP-prescribed medications, and hospital admissions). Missing covariate values were imputed using the random forest-based *missRanger* package in R [3].

**Statistical Analyses**

Proportional hazards Cox regression was employed to model the time to death event of interest. Primary analyses were conducted for post-acute mortality using follow-up starting at the 30-day landmark (T_landmark_/T’_landmark_). Overall mortality analyses (including days 0-30) across the full follow-up period were additionally performed. Descriptive statistics are presented for both the COVID-19 exposure and reference cohorts. Continuous variables are reported as mean (standard deviation), and categorical variables as frequencies (percentages). To mitigate potential convergence issues due to the large number of covariates, we employed a variable selection procedure based on the method proposed by Zhao et al. [4]: after including key standard variables (age, sex, number of hospitalizations and medications within the past year), univariate testing was performed on the remaining covariates. Predictors demonstrating a nominally significant association with the outcome (p < 0.05) were then incorporated into the final multivariable Cox regression model. All statistical analyses were performed using R (version 3.6.1). The false discovery rate (FDR) was used to control for multiple testing, where FDR-adjusted p-values < 0.05 were considered statistically significant. The overall analytic workflow is illustrated in Fig.S1.

Risks of death due to composite outcomes within each body system

HRs for all-cause mortality and death due to *at least one sequela in each body system* were calculated for the COVID-19 exposure cohort, compared with the reference cohort in the post-acute period; as the reference may include undocumented mild or asymptomatic infections, these estimates reflect mortality risk associated with clinically apparent COVID-19 relative to the general population without known infection. The overall exposure cohort was then divided into two mutually exclusive groups: hospitalized and non-hospitalized COVID-19, to analyze mortality risks associated with each post-infection outcome, respectively. Corresponding overall-mortality estimates are reported in the supplementary material.

Risks of mortality due to individual diseases

To estimate HRs for post-acute cause-specific mortality due to each individual CCSR disease outcome, analyses were repeated by comparing the overall, hospitalized, and non-hospitalized COVID-19 cohorts to the reference cohort. Within these COVID-19 cohorts, we also performed stratified analysis to examine each death outcome in patients with and without a prior history of the corresponding CCSR sequela, as described previously in the Outcomes section. The overall mortality analysis for single CCSR outcomes was repeated and reported in the supplementary material.

Subgroup analysis

To assess the heterogeneity of the association between overall COVID-19 infection and mortality (for both composite and single outcomes) across different subgroups, stratified analyses were conducted for both overall and post-acute mortality. We compared individuals *with* (risk-factor-present group) and *without* (risk-factor-absent group) each of the following demographic or comorbidity characteristics: age (advanced age [>65] vs. middle age [50-65]), sex (male vs. female), history of AF, CAD, CKD, COPD, dementia, heart failure, HTN, renal failure, stroke, and T2DM. For each characteristic, we estimated the ratio of hazard ratios (RHR) between the risk-factor-present and risk-factor-absent subgroups by calculating the difference in log-hazard ratios $(\beta_{RHR}= \beta_{present}- \beta_{absent})$, with a standard error estimated as ${SE}_{RHR}=\sqrt{variance\left( \beta_{present} \right)+variance\left( \beta_{absent} \right)}$. We then used a z-statistic $(z= \beta_{RHR}/{SE}_{RHR})$ to assess the significance of the RHR (obtained as $\exp(\beta_{RHR})$) between subgroups. Analyses were not performed if the subgrouping factor itself was also a component of the primary/secondary outcome of interest.

Additional analysis

To evaluate the robustness of the primary post-acute organ-system mortality findings in the overall COVID-19 exposure cohort, several additional analyses were conducted to address factors that could potentially influence effect estimates. For all additional analyses, the same outcome eligibility criteria, landmark design, covariate selection strategy, and FDR correction approach were applied as in the primary analysis.

*Accounting for contamination by undocumented infection*

As described above, the reference cohort was defined by the absence of documented SARS-CoV-2 infection rather than confirmed seronegative status, and may therefore include individuals with undocumented infections. The primary estimates thus reflect the mortality risk of clinically apparent COVID-19 relative to the general population without known infection, with undocumented infections in the reference potentially attenuating effect estimates toward the null. To address this concern, we performed a quantitative bias analysis (QBA) to estimate the magnitude of such contamination by undocumented infection. Let *π* denote the contamination rate (i.e., the proportion of the reference cohort who were truly infected but undocumented), and *γ* denote the mortality HR among these undocumented infections relative to the truly uninfected population. The mortality rate of the reference cohort (*λ_ref_*) can be modeled as a mixture of the undocumented infected and truly uninfected individuals:

$$\begin{aligned} \lambda_{ref} =&\pi\times\lambda_{undocumented} + \left( 1-\pi\right)\times\lambda_{uninfected} \# \\ =&\pi\times\gamma\times\lambda_{uninfected} + \left( 1-\pi\right)\times\lambda_{uninfected} \\ =&\left[ \pi\times\gamma+ \left( 1-\pi\right) \right] \times\lambda_{uninfected}\#\left( 1 \right) \end{aligned}$$

In the primary analysis, the original HR for COVID-19 infection was estimated by comparing mortality between the exposure and reference cohorts. Substituting Equation $\left( 1 \right)$:

$$\begin{aligned} Original HR=&\frac{\lambda_{exp}}{\lambda_{ref}}\# \\ =&\frac{\lambda_{exp}}{\left[ \pi\times\gamma+ \left( 1-\pi\right) \right] \times\lambda_{uninfected}}&\#\left( 2 \right) \end{aligned}$$

Based on Equation $\left( 2 \right)$, the bias-corrected HR (*HR_corrected_*) for the exposure cohort versus truly uninfected individuals was calculated as:

$$\begin{aligned} {HR}_{corrected}=&\frac{\lambda_{exp}}{\lambda_{uninfected}}\# \\ =&\frac{Original HR \times\left[ \pi\times\gamma+ \left( 1-\pi\right) \right] \times\lambda_{uninfected}}{\lambda_{uninfected}} \\ =&Original HR \times\left[ \pi\times\gamma+ \left( 1-\pi\right) \right]\#\left( 3 \right) \end{aligned}$$

where $\left[ \pi\times\gamma+ \left( 1-\pi\right) \right]$ represents the correction factor.

We evaluated two prespecified scenarios:

- In the “moderate” scenario, we assumed *π* = 0.20, derived by subtracting the observed COVID-19 prevalence in our primary analysis (112,611 / [112,611 + 354,120] ≈ 25%) from the UK cumulative SARS-CoV-2 infection prevalence of ~45% by December 2021 (the onset of the fourth COVID-19 wave in the UK, after which cumulative incidence increased rapidly) [5]; for *γ*, we used the HR of post-acute all-cause mortality for non-hospitalized COVID-19 versus the reference cohort (*γ* = 1.05), reflecting the assumption that most undocumented infections were mild or asymptomatic.
- In the “extreme” scenario, we assumed *π* = 0.45, based on a cumulative UK prevalence of ~70% by mid-2022 [5]; *γ* was set to the HR of post-acute all-cause mortality for overall COVID-19 versus the reference cohort (*γ* = 1.50), representing the assumption that undocumented infections carried the same average mortality risks as all documented infections regardless of severity.

As a complementary analysis, we repeated the primary analysis using a negative-test restricted reference cohort, defined as participants who had undergone SARS-CoV-2 testing during the follow-up period but never received a positive result (N = 169,400), thereby providing a more stringent reference group with reduced likelihood of undocumented infections.

*Accounting for vaccination status*

To examine the potential modifying effect of COVID-19 vaccination on post-acute mortality, we performed two analyses. First, we stratified the overall COVID-19 exposure cohort by vaccination status (receipt of any vaccine dose(s) versus no vaccination) and separately estimated HRs for each stratum relative to the reference cohort.

Second, we formally tested for effect modification by including a multiplicative interaction term between COVID-19 exposure status and vaccination status in the Cox regression model for each composite outcome.

*Accounting for reinfection-based censoring*

To assess the sensitivity of our findings to the censoring of follow-up at reinfection, we repeated the analysis with follow-up extended beyond the date of reinfection, allowing deaths occurring after subsequent SARS-CoV-2 infections to be captured.

Furthermore, we evaluated whether post-acute mortality risks varied according to the number of SARS-CoV-2 infections during the study period as an exploratory analysis. The COVID-19 exposure cohort was stratified into three mutually exclusive groups: single infection, one reinfection, and two or more reinfections. For all groups, follow-up commenced on the date of the first SARS-CoV-2 infection. Separate Cox regression models were then fitted for each subgroup relative to the same reference cohort.

***Full results***

***Overview***

The COVID-19 exposure cohort included 113,057 participants (61,971 [54.8%] females), with a median follow-up of 274 days (maximum 969 days). This cohort was further divided into 16,628 individuals in the hospitalized COVID-19 group (8,207 [49.4%] females; median follow-up 305 days; maximum 969 days) and 96,429 in the non-hospitalized group (53,764 [55.8%] females; median follow-up 268 days; maximum 921 days). The reference cohort included 354,465 participants without known COVID-19 infection (197,147 [55.6%] females), with a median follow-up of 365 days (maximum 998 days).

For the primary post-acute mortality analysis, we excluded participants (overall COVID-19: 446; reference: 345) who died within the 30-day landmark, yielding samples of 112,611 COVID-19 (median follow-up: 247 days; maximum 939 days) and 354,120 reference participants (median follow-up: 335 days; maximum 968 days). The COVID-19 cohort comprised 16,260 hospitalized (median follow-up: 292 days; maximum 939 days) and 96,351 non-hospitalized (median follow-up: 241 days; maximum 891 days) individuals. During follow-up, 4,942 (4.4%) COVID-19 exposure cohort participants experienced reinfection (hospitalized: 1,812 [11.1%]; non-hospitalized: 3,130 [3.2%]). In the reference cohort, 169,400 (47.8%) participants received SARS-CoV-2 testing but never tested positive. Detailed demographic and clinical characteristics for each cohort are presented in Table_S2.

As summarized in Table_S3a, total deaths in the post-acute period amounted to 2,888 (2.56%) in the overall COVID-19 exposure cohort, 2,382 (14.65%) in the hospitalized COVID-19 group, 506 (0.53%) in the non-hospitalized group and 5,317 (1.50%) in the reference cohort. Neoplasms, circulatory, neurological, and respiratory disorders were consistently the most prevalent causes of death across all cohorts. For overall mortality (including days 0-30), the leading death-cause categories remained the same as in the post-acute analysis; the corresponding distributions are provided in Table_S3b.

**Mortality risks of composite outcomes within each body system**

Risks of all-cause and cause-specific mortality stratified by organ system are presented in Fig.1, Fig.S2, and Table_1. In the primary post-acute analysis, compared to the reference group without known infection, all-cause mortality was significantly elevated (FDR-adjusted p < 0.05) in the overall COVID-19 (HR: 1.50, 95% confidence interval (CI): 1.42-1.58) and hospitalized COVID-19 (HR: 3.06, 95% CI: 2.87-3.26) groups, but not in the non-hospitalized COVID-19 (HR: 1.05, 95% CI: 0.98-1.14) group (Fig.1a, Table_1). In the overall-mortality analysis, significantly increased all-cause deaths were observed in the overall (HR: 2.39, 95% CI: 2.29-2.50; subsequent values are presented in the same format), hospitalized (6.29, 5.99-6.61), and non-hospitalized COVID-19 (1.23, 1.15-1.32) cohorts (Fig.S2a, Table_1).

For organ-system outcomes, all significant associations identified between clinically apparent COVID-19 infection (overall, hospitalized, and non-hospitalized) and death demonstrated increased risks (HR >1) of cause-specific mortality (Fig.1b, Fig.S2b, Table_1). Notably, the hospitalized COVID-19 group exhibited significantly increased mortality risks in all 11 examined organ systems in both the post-acute and overall-mortality analyses. Within the overall COVID-19 exposure cohort, significantly elevated post-acute mortality risk was identified in 7 out of 11 (63.6%) organ systems, including diseases of the circulatory system (HR: 1.22, 95% CI: 1.05-1.43), digestive system (1.50, 1.04-2.16), external causes (3.75, 1.98-7.09), genitourinary system (2.19, 1.29-3.71), neoplasms (1.38, 1.26-1.51), nervous system (2.13, 1.78-2.56), and respiratory system (1.21, 1.04-1.41). For overall mortality, significant associations in the overall COVID-19 exposure cohort were observed for 8 out of 12 (66.7%) organ systems, additionally including diseases of the blood and blood-forming organs (HR: 7.85, 95% CI: 1.80-34.33). For the non-hospitalized cohort, 2 out of 7 (28.6%) body systems were found significant in the post-acute analysis, with increased risks of death from external causes (HR: 2.77, 95% CI: 1.29-5.96) and diseases of the nervous system (1.37, 1.09-1.74). These same two organ systems remained significant (2/7, 28.6%) in the overall-mortality analysis.

**Mortality risks of individual CCSR outcomes in the overall COVID-19 cohort**

Fig.S3 and Table_2a summarize the associations between clinically apparent COVID-19 infection and single CCSR outcomes in the post-acute analysis. Among 42 disorders investigated, COVID-19 infection significantly increased mortality risk for 8 (19.0%) conditions. These significant outcomes, ranked from the largest to smallest HR per system, included:

- Circulatory: Peripheral/visceral vascular disease (HR: 4.11, 95% CI: 2.13-7.94)
- External-cause injuries: Accidental/unintentional intent of injury (4.58, 2.30-9.14), and subsequent encounter of external cause codes (4.00, 2.05-7.82).
- Genitourinary: Urinary tract infection (UTI) (3.11, 1.52-6.39).
- Neoplasm: Bile duct cancer (2.30, 1.24-4.25) and unspecified malignant neoplasm (2.13, 1.35-3.37).
- Neurological: Neurocognitive disorders (2.32, 1.81-2.97), and Parkinson’s disease (2.29, 1.64-3.19).

In the overall-mortality analysis (Fig.S4, Table_2a), more single CCSR conditions (17 out of 47, 36.2%) were significantly associated with overall COVID-19, with generally larger effect estimates in magnitude. Additional significance was identified for circulatory disorders (endocarditis/endocardial disease [HR: 3.12, 95% CI: 1.47-6.62], heart failure [2.23, 1.18-4.22], and coronary atherosclerosis/other heart disease [1.51, 1.12-2.03]), digestive disorders (biliary tract disease [3.26, 1.36-7.83] and other gastrointestinal disorders [2.79, 1.36-5.75]), neoplasms (colorectal cancer [1.55, 1.18-2.03]), and respiratory disorders (aspiration pneumonitis [6.56, 1.41-30.51], pneumonia [1.98, 1.26-3.11], and COPD/bronchiectasis [1.53, 1.15-2.04]).

When restricting the outcome to post-acute death *with* a prior history of the examined CCSR disease (Fig.S3, and full results in Table_S4), the pattern of significant associations (12 out of 28, 42.9%) was broadly similar to that of the primary analysis (regardless of any prior history). Five disorders were newly identified as associated with higher mortality risk: hypertension with complications/secondary hypertension, heart failure, colorectal cancer, bladder cancer, and pneumonia. In the overall-mortality analysis, a larger number of associations reached significance (18 out of 33, 54.5%), including coronary atherosclerosis/other heart disease, CKD, COPD/bronchiectasis, and cancers of the stomach and skin (Table_S4).

For post-acute death *without* a prior history, the proportion of significant associations with elevated risks decreased to 10.7% (3 out of 28; Fig.S3, and full results in Table_S4). Three outcomes were significantly associated with increased mortality: peripheral/visceral vascular disease (HR: 3.00, 95% CI: 2.00-4.50), other specified/unspecified nutritional and metabolic disorders (13.40, 2.53-71.07), and neurocognitive disorders (3.07, 1.81-5.20). The overall-mortality analysis further identified significant associations (6 out of 33, 18.2%) for biliary tract disease, other gastrointestinal disorders, and unspecified malignant neoplasm (Table_S4).

**Mortality risks of individual outcomes in the hospitalized COVID-19 cohort**

Compared to the reference cohort, hospitalized COVID-19 infection significantly elevated post-acute mortality risk for 73.0% (27 out of 37) of the examined CCSR outcomes (Table_2b). These outcomes encompassed multiple organ systems, including circulatory, digestive, endocrine, genitourinary, nervous, and respiratory disorders, as well as external-cause injuries, infectious diseases, and neoplasms. In the overall-mortality analysis, a greater proportion of significant outcomes (36 out of 42, 85.7%; Table_2b) accompanied by generally higher HRs were observed.

Stratifying deaths within the hospitalized COVID-19 cohort by prior history of the outcome revealed consistent patterns. Among post-acute deaths *with* a pre-existing diagnosis, 91.7% (22 out of 24; Table_S4) of CCSR disorders were significantly associated with greater mortality risk; the corresponding proportion for overall mortality was 87.1% (27 out of 31; Table_S4). For post-acute deaths *without* a prior history (Table_S4), hospitalized COVID-19 infection was associated with increased mortality for 71.4% (10 out of 14) of conditions. Elevated neoplasm mortality in this subgroup should be interpreted with caution, as this likely reflects detection bias (unmasking of pre-existing undiagnosed malignancies during clinical workup) rather than de novo malignancies caused by COVID-19. Such associations are mechanistically distinct from other outcomes such as circulatory or respiratory mortality. The overall-mortality analysis showed an even higher proportion of significant associations (19 out of 21, 90.5%; Table_S4) for outcomes without pre-existing diagnoses.

**Mortality risks of individual outcomes in the non-hospitalized COVID-19 cohort**

Table_2c indicates that a small proportion of CCSR disorders (3 out of 22, 13.6%) were significantly associated with increased post-acute mortality in individuals with non-hospitalized COVID-19 infection (HR >1). These included accidental/unintentional intent of injury (HR: 4.04, 95% CI: 1.80-9.06), subsequent encounter of external cause codes (3.44, 1.56-7.59), and neurocognitive disorders (1.86, 1.37-2.50). These three outcomes also remained significant in the overall-mortality analysis (3 out of 29, 10.3%; Table_2c).

Analysis of post-acute deaths *with* a prior history (Table_S4) exhibited the same set of significant CCSR disorders (3 out of 9, 33.3%); for overall mortality, one additional significant outcome was identified (4 out of 15, 26.7%; Table_S4): hypertension with complications/secondary hypertension (7.60, 2.22-26.01). Among deaths *without* pre-existing diagnoses (Table_S4), only neurocognitive disorders (2.77, 1.43-5.38) remained significantly associated with increased mortality risk (1 out of 8, 12.5%); the overall-mortality analysis showed a higher proportion (4 out of 10, 40.0%), with peripheral/visceral vascular diseases (3.29, 1.36-7.95), biliary tract disease (4.44, 1.31-15.06), and other gastrointestinal disorders (3.92, 1.46-10.51) also reaching significance (Table_S4).

**Subgroup analysis**

With the detailed results for each characteristic listed in Table_S5-S16, post-acute mortality risks showing significant differences between risk-factor-present and risk-factor-absent subgroups (FDR-adjusted p < 0.05) are summarized for composite organ-system outcomes in Table_3 and Fig.2.

- Although SARS-CoV-2 infection was associated with increased post-acute all-cause mortality (HR >1) in both individuals with and without advanced age (>65 years), AF, CKD, or HTN history, the magnitude of this risk was significantly greater (RHR >1) in the risk-factor-present groups.
- Older adults (>65 years) demonstrated a significantly higher risk of post-acute mortality from circulatory diseases compared to their middle-aged counterparts (50-65 years).
- Patients with AF history experienced higher post-acute mortality risks from nervous system and respiratory diseases following COVID-19 compared to those without AF.

For individual CCSR disease categories, subgroup analysis revealed no statistically significant differences after FDR correction (Table_S5b-S16b).

Subgroup analyses for overall mortality yielded largely consistent findings (Table_S5c-S16c, Table_S5d-S16d). Advanced age, CKD, and HTN similarly amplified the COVID-19-associated all-cause mortality risk, and older adults again showed greater circulatory disease mortality. AF history was associated with higher mortality from nervous system and respiratory diseases, consistent with the post-acute results. At the individual CCSR level, no subgroup differences reached statistical significance after FDR correction, although trends toward stronger associations were observed for heart failure mortality among individuals with AF (Table_S5d) and for hypertension mortality among those with CKD (Table_S7d) (FDR-adjusted p < 0.10).

Notably, post-acute circulatory mortality among middle-aged participants showed a point estimate below 1 (HR: 0.51, 0.28-0.92), though this did not retain significance after FDR correction (adjusted p = 0.15). Given the large number of subgroup comparisons performed, this likely represents a chance finding. This seemingly protective association was also absent when the acute phase was included in the overall mortality analysis (HR: 0.77, 0.48-1.24), further supporting a spurious result. We therefore refrain from drawing substantive conclusions from this observation.

**Additional analysis**

*Accounting for contamination by undocumented infection*

After adjusting for potential contamination of the reference cohort by undocumented infections, the QBA showed consistently higher effect estimates across all composite outcomes compared to the primary analysis (Table_S17). Under both the moderate scenario (*π* = 0.20, *λ* = 1.05; correction factor = 1.01) and the extreme scenario (*π* = 0.45, *λ* = 1.50; correction factor = 1.225), bias-corrected HRs were uniformly amplified; for example, the HR_corrected_ for all-cause mortality increased from 1.50 to 1.52 and 1.84, respectively. These findings suggest that undocumented infections in the reference cohort attenuated estimates toward the null, and that true mortality risks relative to strictly uninfected individuals are likely larger than reported.

Use of a negative-test restricted reference group further supported this interpretation, yielding the same set of significant outcomes as in the primary analysis (Table_S18). With the exception of external causes, for which the HR decreased from 3.75 to 3.59, effect estimates were modestly higher for all-cause mortality (HR: 1.82, 95% CI: 1.73-1.91), circulatory diseases (1.26, 1.09-1.47), digestive diseases (1.65, 1.18-2.30), genitourinary diseases (2.39, 1.45-3.92), neoplasms (1.45, 1.33-1.58), nervous system diseases (2.15, 1.80-2.57), and respiratory diseases (1.31, 1.05-1.64). Together, these results indicate that the primary estimates, which quantify the mortality risk of clinically apparent COVID-19 relative to the general population, are conservative relative to truly uninfected individuals, while the overall pattern of significant associations remained unchanged.

*Accounting for vaccination status*

In stratified analyses, both vaccinated (HR: 1.48, 95% CI: 1.37-1.60) and unvaccinated (1.66, 1.54-1.79) COVID-19 cases exhibited significantly elevated post-acute all-cause mortality relative to the reference cohort (Table_S19). Among vaccinated participants, significant associations were observed for neoplasms (1.35, 1.19-1.53), nervous system diseases (2.12, 1.61-2.79), and respiratory diseases (1.22, 1.04-1.43). The unvaccinated group showed higher risk estimates across most outcomes, with additional significant associations identified for circulatory diseases (1.33, 1.06-1.68), endocrine diseases (5.10, 1.87-13.92), external causes (7.88, 3.08-20.13), and genitourinary diseases (4.24, 2.24-8.03).

Despite these differences in point estimates, no statistically significant interaction between COVID-19 infection and vaccination status was detected for any composite organ-system outcome after FDR correction (Table_S20), providing no strong evidence that vaccination modified the associations between SARS-CoV-2 infection and post-acute mortality.

*Accounting for reinfection-based censoring*

When follow-up was extended beyond the date of reinfection, the resulting estimates were nearly identical to those from the primary analysis that censored follow-up at reinfection (Table_S21). The HR for all-cause mortality was 1.52 (95% CI: 1.44-1.61), compared with 1.50 (1.42-1.58) in the primary analysis, and the same organ systems reached statistical significance in both analyses. These findings suggest that the primary results were not materially affected by the reinfection-censoring strategy.

Stratification by number of SARS-CoV-2 infections revealed an overall increase in post-acute mortality risks with reinfection (Table_S22). Compared with the reference cohort, all-cause mortality was significantly elevated among those with a single infection (HR: 1.47, 95% CI: 1.39-1.55), one reinfection (3.42, 2.72-4.29), and two or more reinfections (3.10, 2.33-4.13). Similar stepwise increases were observed for neoplasms (single infection: 1.36, 1.24-1.49; one reinfection: 2.25, 1.51-3.35; two or more reinfections: 5.06, 2.98-8.60), nervous system diseases (2.02, 1.67-2.44; 6.20, 3.40-11.28; 11.66, 4.69-28.96), and respiratory diseases (1.17, 1.02-1.35; 3.83, 1.56-9.36; 12.43, 5.05-30.63). Circulatory mortality was significantly elevated for single infection (1.21, 1.03-1.42) and two or more reinfections (2.84, 1.22-6.61), but not for one reinfection. Analyses of other organ systems among reinfected groups did not meet eligibility criteria because of sparse events.
