## Supplementary Table 1 to 22 for "Association of COVID-19 with risks of all-cause and cause-specific mortality post-infection: A UK Biobank cohort study"

| **Supplementary Table 1. List of Primary and Secondary Outcomes Included in the Study.** | | | |
| --- | --- | --- | --- |
| **Primary (Composite) Outcomes*** | | **Secondary (Single) Outcomes*** | |
| **3-Character Abbreviation** | **Organ System Description** | **CCSR Category** | **CCSR Category Description** |
| All-cause mortality | |  |  |
| BLD | Diseases of the blood and blood forming organs  and certain disorders involving the immune mechanism |  |  |
| CIR | Diseases of the circulatory system | CIR001 | Chronic rheumatic heart disease |
|  |  | CIR003 | Nonrheumatic and unspecified valve disorders |
|  |  | CIR004 | Endocarditis and endocardial disease |
|  |  | CIR005 | Myocarditis and cardiomyopathy |
|  |  | CIR008 | Hypertension with complications and secondary hypertension |
|  |  | CIR009 | Acute myocardial infarction |
|  |  | CIR011 | Coronary atherosclerosis and other heart disease |
|  |  | CIR019 | Heart failure |
|  |  | CIR020 | Cerebral infarction |
|  |  | CIR021 | Acute hemorrhagic cerebrovascular disease |
|  |  | CIR024 | Other and ill-defined cerebrovascular disease |
|  |  | CIR026 | Peripheral and visceral vascular disease |
|  |  | CIR029 | Aortic; peripheral; and visceral artery aneurysms |
| DIG | Diseases of the digestive system | DIG006 | Gastrointestinal and biliary perforation |
|  |  | DIG012 | Intestinal obstruction and ileus |
|  |  | DIG017 | Biliary tract disease |
|  |  | DIG019 | Other specified and unspecified liver disease |
|  |  | DIG021 | Gastrointestinal hemorrhage |
|  |  | DIG025 | Other specified and unspecified gastrointestinal disorders |
| END | Endocrine nutritional and metabolic diseases | END016 | Other specified and unspecified nutritional and metabolic disorders |
| EXT | External cause codes | EXT020 | External cause codes: intent of injury, accidental/unintentional |
|  |  | EXT029 | External cause codes: subsequent encounter |
| GEN | Diseases of the genitourinary system | GEN003 | Chronic kidney disease |
|  |  | GEN004 | Urinary tract infections |
| INF | Certain infectious and parasitic diseases | INF002 | Septicemia |
|  |  | INF003 | Bacterial infections |
| INJ | Injury poisoning and certain other consequences of external causes |  |  |
| MUS | Diseases of the musculoskeletal system and connective tissue |  |  |
| NEO | Neoplasms | NEO012 | Gastrointestinal cancers - esophagus |
|  |  | NEO013 | Gastrointestinal cancers - stomach |
|  |  | NEO015 | Gastrointestinal cancers - colorectal |
|  |  | NEO017 | Gastrointestinal cancers - liver |
|  |  | NEO018 | Gastrointestinal cancers - bile duct |
|  |  | NEO024 | Sarcoma |
|  |  | NEO025 | Skin cancers - melanoma |
|  |  | NEO043 | Urinary system cancers - bladder |
|  |  | NEO048 | Nervous system cancers - brain |
|  |  | NEO051 | Endocrine system cancers - pancreas |
|  |  | NEO067 | Mesothelioma |
|  |  | NEO068 | Myelodysplastic syndrome (MDS) |
|  |  | NEO071 | Malignant neoplasm, unspecified |
| NVS | Diseases of the nervous system | NVS004 | Parkinson`s disease |
|  |  | NVS005 | Multiple sclerosis |
|  |  | NVS006 | Other specified hereditary and degenerative nervous system conditions |
|  |  | NVS011 | Neurocognitive disorders |
| RSP | Diseases of the respiratory system | RSP002 | Pneumonia (except that caused by tuberculosis) |
|  |  | RSP008 | Chronic obstructive pulmonary disease and bronchiectasis |
|  |  | RSP010 | Aspiration pneumonitis |
|  |  | RSP016 | Other specified and unspecified lower respiratory disease |
| * Analyses were restricted to the composite and single outcomes with ≥5 death events in both the COVID-19 exposure and reference cohorts. | | | |

| **Supplementary Table 2. Characteristics of the UKBB participants in the COVID-19 Exposure and Reference Cohort.** | | | | | |
| --- | --- | --- | --- | --- | --- |
|  | | **Overall COVID-19** | **Hospitalized^1^ COVID-19** | **Non-hospitalized COVID-19** | **Reference cohort^2^** |
| **Post-acute Mortality^3^** | **N included** | 112,611 | 16,260 | 96,351 | 354,120 |
|  |  |  |  |  | *(169,400 [47.8%] undergone SARS-CoV-2 test & never had positive records)* |
|  | **N of death before 30-day landmark** | 446 | 368 | 78 | 345 |
|  | **Median follow-up length (days)** | 247 | 292 | 241 | 335 |
|  | **Max. follow-up length (days)** | 939 | 939 | 891 | 968 |
|  | **N reinfected within the follow-up** | 4,942 (4.4%) | 1,812 (11.1%) | 3,130 (3.2%) | N/A |
|  | 1^st^ reinfection | 4,422 | 1,461 | 2,961 |  |
|  | 2^nd^ or more reinfection | 520 | 351 | 169 |  |
| **Overall Mortality^4^** | **Total N** | 113,057 | 16,628 | 96,429 | 354,465 |
|  | **Median follow-up length (days)** | 274 | 305 | 268 | 365 |
|  | **Max. follow-up length (days)** | 969 | 969 | 921 | 998 |
| **Age^5^** | | 66.23 (8.20) | 68.77 (8.62) | 65.79 (8.04) | 68.65 (7.97) |
| **Sex^6^ (%)** | |  |  |  |  |
| Male | | 51,086 (45.2%) | 8,421 (50.6%) | 42,665 (44.2%) | 157,318 (44.4%) |
| Female | | 61,971 (54.8%) | 8,207 (49.4%) | 53,764 (55.8%) | 197,147 (55.6%) |
| **Ethnicity (%)** | |  |  |  |  |
| White | | 106,633 (94.3%) | 15,299 (92.0%) | 91,334 (94.7%) | 333,325 (94.0%) |
| Non-White | | 6,424 (5.7%) | 1,329 (8.0%) | 5,095 (5.3%) | 21,140 (6.0%) |
| **Townsend Deprivation Index** | | -1.38 (3.00) | -0.73 (3.27) | -1.49 (2.93) | -1.32 (3.09) |
| **Smoking status (%)** | |  |  |  |  |
| Never | | 64,007 (56.6%) | 8,110 (48.8%) | 55,897 (58.0%) | 197,685 (55.8%) |
| Previous | | 39,090 (34.6%) | 6,399 (38.5%) | 32,691 (34.0%) | 120,410 (34.0%) |
| Current | | 9,960 (8.8%) | 2,119 (12.7%) | 7.841 (8.0%) | 36,370 (10.2%) |
| **BMI** | | 27.47 (4.82) | 28.73 (5.39) | 27.26 (4.68) | 27.34 (4.73) |
| **Waist circumference** | | 90.30 (13.45) | 94.25 (14.45) | 89.62 (13.15) | 89.91 (13.21) |
| **HDL** | | 1.54 (0.43) | 1.44 (0.43) | 1.56 (0.43) | 1.54 (0.44) |
| **LDL** | | 2.09 (1.04) | 2.20 (1.17) | 2.07 (1.02) | 2.28 (1.15) |
| **Triglycerides** | | 1.55 (0.94) | 1.69 (1.03) | 1.52 (0.92) | 1.57 (0.97) |
| **Glucose** | | 5.24 (1.52) | 5.54 (2.10) | 5.18 (1.39) | 5.25 (1.48) |
| **HbA1c** | | 32.04 (11.35) | 33.88 (12.72) | 31.72 (11.07) | 32.63 (11.17) |
| **Haemoglobin** | | 13.92 (1.31) | 13.71 (1.51) | 13.96 (1.27) | 13.97 (1.29) |
| **Systemic autoimmune diseases (%)** | | 4,552 (4.0%) | 1,066 (6.4%) | 3,486 (3.6%) | 12,393 (3.5%) |
| **Organ-specific autoimmune diseases (%)** | | 10,608 (9.4%) | 1,720 (10.3%) | 8,888 (9.2%) | 28,924 (8.2%) |
| **Immunodeficiency (%)** | | 1,109 (1.0%) | 269 (1.6%) | 840 (0.9%) | 2,717 (0.8%) |
| **Immunosuppressant use (%)** | | 3,263 (2.9%) | 800 (4.8%) | 2,463 (2.6%) | 7,243 (2.0%) |
| **AF history(%)** | | 7,307 (6.5%) | 2,083 (12.5%) | 5,224 (5.4%) | 22,910 (6.5%) |
| **CAD history(%)** | | 17,570 (15.5%) | 3,898 (23.4%) | 13,672 (14.2%) | 53,449 (15.1%) |
| **CKD history (%)** | | 17,414 (15.4%) | 3,674 (22.1%) | 13,740 (14.2%) | 54,346 (15.3%) |
| **COPD history(%)** | | 6,565 (5.8%) | 2,067 (12.4%) | 4,498 (4.7%) | 19,937 (5.6%) |
| **Cancer history (%)** | | 7,440 (6.6%) | 1,378 (8.3%) | 6,062 (6.3%) | 25,217 (7.1%) |
| **Dementia history (%)** | | 3,914 (3.5%) | 1,189 (7.2%) | 2,725 (2.8%) | 8,975 (2.5%) |
| **HTN history (%)** | | 42,600 (37.7%) | 8,644 (52.0%) | 33,956 (35.2%) | 140,183 (39.5%) |
| **Non-COVID Pneumonia history (%)** | | 10,637 (9.4%) | 2,658 (16.0%) | 7,979 (8.3%) | 29,205 (8.2%) |
| **Stroke history (%)** | | 5,771 (5.1%) | 1,904 (11.5%) | 3,867 (4.0%) | 17,954 (5.1%) |
| **T2DM history (%)** | | 9,677 (8.6%) | 2,857 (17.2%) | 6,820 (7.1%) | 29,180 (8.2%) |
| **Vaccination status (%)** | |  |  |  |  |
| Receiving any doses | | 101,569 (89.8%) | 11,230 (67.5%) | 90,339 (93.7%) | 279,978 (79.0%) |
| No vaccination | | 11,488 (10.2%) | 5,398 (32.5%) | 6,090 (6.3%) | 74,487 (21.0%) |
| **Number of non-cancer illness^7^** | | 1.78 (1.83) | 2.35 (2.17) | 1.69 (1.75) | 1.81 (1.82) |
| **Number of GP medications prescribed^7^** | | 4.56 (4.76) | 6.16 (6.54) | 4.29 (4.32) | 4.06 (4.58) |
| **Number of hospitalizations^7^** | | 0.54 (2.44) | 1.22 (5.12) | 0.42 (1.53) | 0.44 (1.78) |
| Demographic/clinical characteristics were selected based on potential relevance to COVID-19 and its complications. 1. Hospitalized COVID-19 cohort refers to UKBB participants with an associated hospitalization record due to U07.1, the ICD-10 codes for fatal (laboratory-confirmed) COVID-19 infection. 2. The reference cohort includes all UKBB participants without a known diagnosis or those tested negative for COVID-19, regardless of hospitalization status. 3. Post-acute Mortality measures the risk of death by setting the follow-up start date as 30 days after the date of first positive SARS-CoV-2 test. 4. Overall Mortality measures the risk of death by setting the follow-up start date as the date of first positive SARS-CoV-2 test. 5. Continuous variables (without (%) following) are presented as mean (standard deviation). 6. Categorical variables (with (%) following) are presented as frequencies (percentages). 7. Number of non-cancer illness, GP medications prescribed and hospitalization in the past year were measured. | | | | | |

| **Supplementary Table 3. Total Number (Percentage) of Death Causes Stratified by Composite Outcomes in Different Cohorts.** | | | | |
| --- | --- | --- | --- | --- |
| ***a. Distribution of Death Causes of Post-acute Mortality^1^*** | | | | |
|  | **Overall COVID-19 (n = 112,611)** | **Hospitalized^2^ COVID-19 (n = 16,260)** | **Non-hospitalized COVID-19 (n = 96,351)** | **Reference cohort^3^ (n = 354,120)** |
| All-cause mortality | 2888 (2.56%) | 2382 (14.65%) | 506 (0.53%) | 5317 (1.50%) |
| (**CIR**) Diseases of the circulatory system | 206 (0.18%) | 132 (0.81%) | 74 (0.08%) | 907 (0.26%) |
| (**DIG**) Diseases of the digestive system | 43 (0.04%) | 28 (0.17%) | 15 (0.02%) | 146 (0.04%) |
| (**END**) Endocrine nutritional and metabolic diseases | 10 (0.01%) | 7 (0.04%) | 3 (0.003%) | 34 (0.01%) |
| (**EXT**) External cause codes | 18 (0.02%) | 12 (0.07%) | 6 (0.01%) | 51 (0.01%) |
| (**GEN**) Diseases of the genitourinary system | 26 (0.02%) | 23 (0.14%) | 3 (0.003%) | 54 (0.02%) |
| (**INF**) Certain infectious and parasitic diseases | 28 (0.02%) | 19 (0.12%) | 9 (0.01%) | 104 (0.03%) |
| (**INJ**) Injury poisoning and certain other consequences of external causes | 13 (0.01%) | 10 (0.06%) | 3 (0.003%) | 55 (0.02%) |
| (**MUS**) Diseases of the musculoskeletal system and connective tissue | 13 (0.01%) | 8 (0.05%) | 5 (0.01%) | 29 (0.01%) |
| (**NEO**) Neoplasms | 665 (0.59%) | 462 (2.84%) | 203 (0.21%) | 2414 (0.68%) |
| (**NVS**) Diseases of the nervous system | 218 (0.19%) | 122 (0.75%) | 96 (0.10%) | 314 (0.09%) |
| (**RSP**) Diseases of the respiratory system | 107 (0.10%) | 80 (0.49%) | 27 (0.03%) | 338 (0.10%) |
| ***b. Distribution of Death Causes of Overall Mortality^4^*** | | | | |
|  | **Overall COVID-19 (n = 113,057)** | **Hospitalized^2^ COVID-19 (n = 16,628)** | **Non-hospitalized COVID-19 (n = 96,429)** | **Reference cohort^3^ (n = 354,465)** |
| All-cause mortality | 3,334 (2.95%) | 2,750 (16.54%) | 584 (0.61%) | 5662 (1.60%) |
| (**BLD**) Diseases of the blood and blood forming organs  and certain disorders involving the immune mechanism | 5 (0.004%) | 4 (0.02%) | 1 (0.001%) | 7 (0.002%) |
| (**CIR**) Diseases of the circulatory system | 269 (0.24%) | 190 (1.14%) | 79 (0.08%) | 961 (0.27%) |
| (**DIG**) Diseases of the digestive system | 63 (0.06%) | 48 (0.29%) | 15 (0.02%) | 159 (0.04%) |
| (**END**) Endocrine nutritional and metabolic diseases | 12 (0.01%) | 9 (0.05%) | 3 (0.003%) | 36 (0.01%) |
| (**EXT**) External cause codes | 20 (0.02%) | 10 (0.06%) | 10 (0.01%) | 55 (0.02%) |
| (**GEN**) Diseases of the genitourinary system | 33 (0.03%) | 30 (0.18%) | 3 (0.003%) | 58 (0.02%) |
| (**INF**) Certain infectious and parasitic diseases | 36 (0.03%) | 27 (0.16%) | 9 (0.01%) | 112 (0.03%) |
| (**INJ**) Injury poisoning and certain other consequences of external causes | 17 (0.02%) | 14 (0.08%) | 3 (0.003%) | 58 (0.02%) |
| (**MUS**) Diseases of the musculoskeletal system and connective tissue | 15 (0.01%) | 10 (0.06%) | 5 (0.01%) | 31 (0.01%) |
| (**NEO**) Neoplasms | 812 (0.72%) | 602 (3.62%) | 210 (0.22%) | 2528 (0.71%) |
| (**NVS**) Diseases of the nervous system | 247 (0.22%) | 148 (0.89%) | 99 (0.10%) | 340 (0.10%) |
| (**RSP**) Diseases of the respiratory system | 129 (0.11%) | 101 (0.61%) | 28 (0.03%) | 355 (0.10%) |
| 1. Post-acute Mortality measures the risk of death by setting the follow-up start date as 30 days after the date of first positive SARS-CoV-2 test. 2. Hospitalized COVID-19 cohort refers to UKBB participants with an associated hospitalization record due to U07.1, the ICD-10 codes for fatal (laboratory-confirmed) COVID-19 infection. 3. The reference cohort includes all UKBB participants without a known diagnosis or those tested negative for COVID-19, regardless of hospitalization status. 4. Overall Mortality measures the risk of death by setting the follow-up start date as the date of first positive SARS-CoV-2 test. | | | | |

**Supplementary Table 4. Associations Between Cause-Specific Mortality Risks in Single CCSR Disorders With/Without Prior History and Overall/Hospitalized/Non-hospitalized COVID-19 Using Cox Regression Model.**

| **Analysis** | **Death Status** | **Outcome** | **Post-acute Mortality^1^** | | **Overall Mortality^2^** | | **CCSR Category Description** |
| --- | --- | --- | --- | --- | --- | --- | --- |
|  |  |  | **HR (95% CI)** | **p.adj^3^** | **HR (95% CI)** | **p.adj^3^** |  |
| Overall COVID-19 vs. reference | Death with a prior history | CIR001 | N/A^4^ | | 1.07 (0.33 - 3.47) | 9.64E-01 | Chronic rheumatic heart disease |
|  |  | CIR003 | 0.72 (0.24 - 2.16) | 6.26E-01 | 1.23 (0.51 - 2.97) | 7.63E-01 | Nonrheumatic and unspecified valve disorders |
|  |  | CIR005 | N/A | | 1.08 (0.42 - 2.77) | 9.64E-01 | Myocarditis and cardiomyopathy |
|  |  | CIR008 | **6.35 (1.82 - 22.25)** | **1.53E-02** | **6.05 (1.98 - 18.45)** | **4.39E-03** | Hypertension with complications and secondary hypertension |
|  |  | CIR009 | 1.03 (0.45 - 2.36) | 9.41E-01 | 1.26 (0.65 - 2.45) | 5.94E-01 | Acute myocardial infarction |
|  |  | CIR011 | 1.39 (0.95 - 2.03) | 1.50E-01 | **1.66 (1.17 - 2.35)** | **1.09E-02** | Coronary atherosclerosis and other heart disease |
|  |  | CIR019 | **3.38 (1.31 - 8.74)** | **3.68E-02** | **4.54 (1.87 - 10.98)** | **3.01E-03** | Heart failure |
|  |  | CIR026 | **7.07 (2.18 - 22.94)** | **5.20E-03** | **7.07 (2.18 - 22.94)** | **3.79E-03** | Peripheral and visceral vascular disease |
|  |  | DIG019 | 1.72 (0.56 - 5.31) | 4.39E-01 | 1.99 (0.75 - 5.29) | 2.27E-01 | Other specified and unspecified liver disease |
|  |  | END016 | N/A | | 0.99 (0.30 - 3.30) | 9.89E-01 | Other specified and unspecified nutritional and metabolic disorders |
|  |  | EXT020 | **5.35 (2.29 - 12.48)** | **7.24E-04** | **4.38 (2.02 - 9.51)** | **1.07E-03** | External cause codes: intent of injury, accidental/unintentional |
|  |  | EXT029 | **6.01 (2.51 - 14.38)** | **5.33E-04** | **4.77 (2.16 - 10.53)** | **7.66E-04** | External cause codes: subsequent encounter |
|  |  | GEN003 | 3.76 (1.15 - 12.33) | 6.21E-02 | **3.87 (1.43 - 10.44)** | **1.62E-02** | Chronic kidney disease |
|  |  | GEN004 | **6.30 (2.16 - 18.35)** | **4.11E-03** | **7.10 (2.48 - 20.35)** | **1.27E-03** | Urinary tract infections |
|  |  | INF003 | N/A | | 2.15 (0.75 - 6.18) | 2.18E-01 | Bacterial infections |
|  |  | NEO012 | 0.96 (0.50 - 1.83) | 9.40E-01 | 1.00 (0.56 - 1.78) | 9.89E-01 | Gastrointestinal cancers - esophagus |
|  |  | NEO013 | 2.22 (0.86 - 5.71) | 1.63E-01 | **2.94 (1.36 - 6.34)** | **1.40E-02** | Gastrointestinal cancers - stomach |
|  |  | NEO015 | **1.51 (1.07 - 2.12)** | **4.40E-02** | **1.71 (1.26 - 2.33)** | **2.68E-03** | Gastrointestinal cancers - colorectal |
|  |  | NEO017 | 0.80 (0.25 - 2.54) | 7.54E-01 | 0.88 (0.32 - 2.45) | 9.18E-01 | Gastrointestinal cancers - liver |
|  |  | NEO018 | **2.95 (1.23 - 7.11)** | **4.34E-02** | **2.73 (1.19 - 6.23)** | **3.27E-02** | Gastrointestinal cancers - bile duct |
|  |  | NEO025 | 2.66 (0.98 - 7.20) | 1.01E-01 | **3.04 (1.17 - 7.91)** | **4.09E-02** | Skin cancers - melanoma |
|  |  | NEO043 | **2.37 (1.22 - 4.60)** | **3.68E-02** | **3.07 (1.74 - 5.41)** | **7.66E-04** | Urinary system cancers - bladder |
|  |  | NEO048 | 1.41 (0.73 - 2.76) | 4.31E-01 | 1.58 (0.86 - 2.91) | 2.10E-01 | Nervous system cancers - brain |
|  |  | NEO051 | 1.25 (0.74 - 2.10) | 4.74E-01 | 1.59 (1.01 - 2.51) | 7.37E-02 | Endocrine system cancers - pancreas |
|  |  | NEO068 | 1.89 (0.56 - 6.33) | 4.31E-01 | 1.63 (0.55 - 4.84) | 4.94E-01 | Myelodysplastic syndrome (MDS) |
|  |  | NEO071 | N/A | | **4.42 (1.31 - 14.87)** | **3.27E-02** | Malignant neoplasm, unspecified |
|  |  | NVS004 | **2.32 (1.64 - 3.27)** | **2.70E-05** | **2.21 (1.60 - 3.06)** | **3.03E-05** | Parkinson`s disease |
|  |  | NVS005 | 1.91 (0.78 - 4.65) | 2.43E-01 | 1.90 (0.82 - 4.38) | 2.07E-01 | Multiple sclerosis |
|  |  | NVS006 | 1.55 (0.64 - 3.73) | 4.38E-01 | 2.01 (0.90 - 4.52) | 1.45E-01 | Other specified hereditary and degenerative nervous system conditions |
|  |  | NVS011 | **3.47 (2.58 - 4.67)** | **4.75E-15** | **3.65 (2.76 - 4.83)** | **2.96E-18** | Neurocognitive disorders |
|  |  | RSP002 | **2.64 (1.19 - 5.86)** | **4.34E-02** | **3.09 (1.54 - 6.20)** | **4.39E-03** | Pneumonia (except that caused by tuberculosis) |
|  |  | RSP008 | 1.39 (1.00 - 1.91) | 9.46E-02 | **1.55 (1.14 - 2.09)** | **1.09E-02** | Chronic obstructive pulmonary disease and bronchiectasis |
|  |  | RSP016 | 0.70 (0.30 - 1.64) | 4.74E-01 | 0.72 (0.32 - 1.64) | 5.51E-01 | Other specified and unspecified lower respiratory disease |
|  | Death without a prior history | CIR004 | N/A | | 2.47 (1.03 - 5.91) | 1.39E-01 | Endocarditis and endocardial disease |
|  |  | CIR008 | 0.66 (0.27 - 1.61) | 4.62E-01 | 0.73 (0.32 - 1.67) | 5.76E-01 | Hypertension with complications and secondary hypertension |
|  |  | CIR009 | 0.67 (0.45 - 1.00) | 1.59E-01 | 0.85 (0.60 - 1.21) | 5.59E-01 | Acute myocardial infarction |
|  |  | CIR011 | 1.13 (0.60 - 2.11) | 7.59E-01 | 1.15 (0.63 - 2.10) | 7.26E-01 | Coronary atherosclerosis and other heart disease |
|  |  | CIR020 | N/A | | 1.13 (0.38 - 3.36) | 8.56E-01 | Cerebral infarction |
|  |  | CIR021 | 1.27 (0.79 - 2.06) | 4.37E-01 | 1.47 (0.94 - 2.29) | 2.35E-01 | Acute hemorrhagic cerebrovascular disease |
|  |  | CIR024 | 3.26 (0.92 - 11.56) | 1.72E-01 | 4.14 (1.34 - 12.83) | 6.42E-02 | Other and ill-defined cerebrovascular disease |
|  |  | CIR026 | **3.00 (2.00 - 4.50)** | **2.94E-06** | **3.51 (1.71 - 7.21)** | **9.98E-03** | Peripheral and visceral vascular disease |
|  |  | DIG006 | N/A | | 2.26 (0.70 - 7.24) | 3.78E-01 | Gastrointestinal and biliary perforation |
|  |  | DIG012 | 1.10 (0.24 - 5.14) | 9.03E-01 | 0.73 (0.16 - 3.37) | 7.26E-01 | Intestinal obstruction and ileus |
|  |  | DIG017 | 2.81 (0.90 - 8.81) | 1.78E-01 | **4.38 (1.57 - 12.16)** | **3.07E-02** | Biliary tract disease |
|  |  | DIG021 | 5.31 (1.32 - 21.31) | 8.52E-02 | 2.44 (0.76 - 7.82) | 3.15E-01 | Gastrointestinal hemorrhage |
|  |  | DIG025 | 3.17 (1.19 - 8.49) | 8.52E-02 | **3.79 (1.59 - 9.03)** | **2.13E-02** | Other specified and unspecified gastrointestinal disorders |
|  |  | END016 | 1**3.40 (2.53 - 71.07)** | **2.14E-02** | **13.40 (2.53 - 71.07)** | **2.13E-02** | Other specified and unspecified nutritional and metabolic disorders |
|  |  | GEN004 | 2.08 (0.70 - 6.24) | 3.54E-01 | 3.24 (1.25 - 8.41) | 6.42E-02 | Urinary tract infections |
|  |  | INF002 | 2.03 (0.71 - 5.79) | 3.54E-01 | 1.68 (0.61 - 4.62) | 5.35E-01 | Septicemia |
|  |  | INF003 | 2.64 (0.81 - 8.57) | 2.31E-01 | 1.97 (0.65 - 6.02) | 4.52E-01 | Bacterial infections |
|  |  | NEO012 | 0.65 (0.29 - 1.44) | 4.37E-01 | 0.73 (0.34 - 1.56) | 5.75E-01 | Gastrointestinal cancers - esophagus |
|  |  | NEO013 | N/A | | 1.08 (0.43 - 2.71) | 8.70E-01 | Gastrointestinal cancers - stomach |
|  |  | NEO015 | 1.20 (0.67 - 2.14) | 6.55E-01 | 1.24 (0.71 - 2.19) | 5.76E-01 | Gastrointestinal cancers - colorectal |
|  |  | NEO017 | N/A | | 1.25 (0.45 - 3.46) | 7.26E-01 | Gastrointestinal cancers - liver |
|  |  | NEO018 | 1.78 (0.73 - 4.35) | 3.54E-01 | 2.30 (1.01 - 5.24) | 1.39E-01 | Gastrointestinal cancers - bile duct |
|  |  | NEO025 | 1.59 (0.71 - 3.56) | 4.26E-01 | 1.67 (0.78 - 3.58) | 3.93E-01 | Skin cancers - melanoma |
|  |  | NEO043 | 0.58 (0.20 - 1.71) | 4.37E-01 | 0.58 (0.20 - 1.71) | 5.35E-01 | Urinary system cancers - bladder |
|  |  | NEO048 | 1.52 (0.69 - 3.38) | 4.37E-01 | 1.45 (0.66 - 3.20) | 5.59E-01 | Nervous system cancers - brain |
|  |  | NEO051 | 0.59 (0.33 - 1.03) | 1.72E-01 | 0.78 (0.47 - 1.27) | 5.35E-01 | Endocrine system cancers - pancreas |
|  |  | NEO071 | 1.87 (1.15 - 3.03) | 7.79E-02 | **1.91 (1.20 - 3.04)** | **3.40E-02** | Malignant neoplasm, unspecified |
|  |  | NVS004 | 5.26 (1.02 - 26.96) | 1.59E-01 | 3.93 (0.89 - 17.42) | 1.97E-01 | Parkinson`s disease |
|  |  | NVS006 | 4.06 (1.24 - 13.36) | 8.52E-02 | 4.06 (1.24 - 13.36) | 7.67E-02 | Other specified hereditary and degenerative nervous system conditions |
|  |  | NVS011 | **3.07 (1.81 - 5.20)** | **4.31E-04** | **3.31 (2.00 - 5.46)** | **1.02E-04** | Neurocognitive disorders |
|  |  | RSP002 | 1.18 (0.60 - 2.35) | 7.31E-01 | 1.32 (0.70 - 2.49) | 5.73E-01 | Pneumonia (except that caused by tuberculosis) |
|  |  | RSP008 | 1.21 (0.36 - 4.10) | 7.90E-01 | 1.41 (0.47 - 4.26) | 6.63E-01 | Chronic obstructive pulmonary disease and bronchiectasis |
|  |  | RSP016 | 0.78 (0.24 - 2.53) | 7.59E-01 | 1.28 (0.47 - 3.47) | 7.26E-01 | Other specified and unspecified lower respiratory disease |
| Hospitalized COVID-19  vs. reference | Death with a prior history | CIR003 | 0.99 (0.26 - 3.78) | 9.87E-01 | 1.40 (0.42 - 4.59) | 5.83E-01 | Nonrheumatic and unspecified valve disorders |
|  |  | CIR005 | N/A^4^ | | 2.51 (0.87 - 7.22) | 9.71E-02 | Myocarditis and cardiomyopathy |
|  |  | CIR008 | N/A | | **8.27 (2.14 - 31.89)** | **2.89E-03** | Hypertension with complications and secondary hypertension |
|  |  | CIR009 | 2.02 (0.84 - 4.89) | 1.22E-01 | **2.79 (1.39 - 5.61)** | **4.92E-03** | Acute myocardial infarction |
|  |  | CIR011 | **2.54 (1.60 - 4.01)** | **1.41E-04** | **3.20 (2.12 - 4.83)** | **9.64E-08** | Coronary atherosclerosis and other heart disease |
|  |  | CIR019 | **5.54 (1.82 - 16.85)** | **3.58E-03** | **8.79 (3.26 - 23.68)** | **3.03E-05** | Heart failure |
|  |  | CIR026 | **16.94 (4.86 - 59.06)** | **2.16E-05** | **16.94 (4.86 - 59.06)** | **1.70E-05** | Peripheral and visceral vascular disease |
|  |  | DIG019 | N/A | | **4.56 (1.46 - 14.19)** | **1.06E-02** | Other specified and unspecified liver disease |
|  |  | END016 | N/A | | 2.91 (0.78 - 10.83) | 1.17E-01 | Other specified and unspecified nutritional and metabolic disorders |
|  |  | EXT020 | **16.54 (6.65 - 41.16)** | **7.81E-09** | **13.44 (5.81 - 31.10)** | **5.82E-09** | External cause codes: intent of injury, accidental/unintentional |
|  |  | EXT029 | **18.18 (7.10 - 46.52)** | **7.81E-09** | **14.31 (6.07 - 33.76)** | **5.82E-09** | External cause codes: subsequent encounter |
|  |  | GEN003 | **6.98 (1.90 - 25.65)** | **4.49E-03** | **7.44 (2.52 - 21.95)** | **4.29E-04** | Chronic kidney disease |
|  |  | GEN004 | **12.80 (4.35 - 37.60)** | **1.07E-05** | **15.45 (5.31 - 44.97)** | **1.50E-06** | Urinary tract infections |
|  |  | INF003 | N/A | | **5.15 (1.62 - 16.31)** | **6.60E-03** | Bacterial infections |
|  |  | NEO012 | **2.57 (1.22 - 5.40)** | **1.38E-02** | **2.81 (1.46 - 5.40)** | **2.69E-03** | Gastrointestinal cancers - esophagus |
|  |  | NEO013 | **7.60 (2.74 - 21.09)** | **1.70E-04** | **10.74 (4.73 - 24.40)** | **5.02E-08** | Gastrointestinal cancers - stomach |
|  |  | NEO015 | **3.94 (2.53 - 6.13)** | **7.81E-09** | **5.15 (3.51 - 7.54)** | **4.32E-16** | Gastrointestinal cancers - colorectal |
|  |  | NEO017 | N/A | | 1.75 (0.53 - 5.78) | 3.74E-01 | Gastrointestinal cancers - liver |
|  |  | NEO018 | **10.96 (4.18 - 28.75)** | **3.95E-06** | **9.81 (4.01 - 24.02)** | **1.56E-06** | Gastrointestinal cancers - bile duct |
|  |  | NEO025 | **7.21 (2.37 - 21.90)** | **7.49E-04** | **8.46 (2.97 - 24.10)** | **1.03E-04** | Skin cancers - melanoma |
|  |  | NEO043 | **8.46 (4.19 - 17.06)** | **9.71E-09** | **11.16 (6.20 - 20.08)** | **6.45E-15** | Urinary system cancers - bladder |
|  |  | NEO048 | **4.68 (2.11 - 10.36)** | **2.26E-04** | **5.89 (2.91 - 11.92)** | **1.91E-06** | Nervous system cancers - brain |
|  |  | NEO051 | **2.51 (1.25 - 5.04)** | **1.18E-02** | **3.88 (2.17 - 6.93)** | **9.56E-06** | Endocrine system cancers - pancreas |
|  |  | NEO068 | **5.20 (1.45 - 18.64)** | **1.29E-02** | **4.13 (1.28 - 13.35)** | **2.02E-02** | Myelodysplastic syndrome (MDS) |
|  |  | NEO071 | N/A | | **21.03 (5.77 - 76.62)** | **8.29E-06** | Malignant neoplasm, unspecified |
|  |  | NVS004 | **6.97 (4.63 - 10.51)** | **1.96E-19** | **6.70 (4.57 - 9.82)** | **3.12E-21** | Parkinson`s disease |
|  |  | NVS005 | **7.32 (2.72 - 19.66)** | **1.47E-04** | **7.56 (3.00 - 19.10)** | **3.13E-05** | Multiple sclerosis |
|  |  | NVS006 | **4.77 (1.67 - 13.63)** | **4.49E-03** | **6.09 (2.28 - 16.28)** | **4.54E-04** | Other specified hereditary and degenerative nervous system conditions |
|  |  | NVS011 | **7.11 (4.86 - 10.39)** | **1.06E-22** | **8.44 (5.98 - 11.93)** | **3.02E-32** | Neurocognitive disorders |
|  |  | RSP002 | **6.03 (2.49 - 14.62)** | **1.41E-04** | **6.90 (3.20 - 14.89)** | **1.91E-06** | Pneumonia (except that caused by tuberculosis) |
|  |  | RSP008 | **2.29 (1.60 - 3.27)** | **1.38E-05** | **2.70 (1.94 - 3.75)** | **1.24E-08** | Chronic obstructive pulmonary disease and bronchiectasis |
|  | Death without a prior history | CIR004 | N/A | | **7.37 (2.78 - 19.56)** | **1.83E-04** | Endocarditis and endocardial disease |
|  |  | CIR008 | N/A | | 1.52 (0.55 - 4.23) | 4.23E-01 | Hypertension with complications and secondary hypertension |
|  |  | CIR009 | 1.43 (0.85 - 2.43) | 2.12E-01 | **2.02 (1.30 - 3.14)** | **3.51E-03** | Acute myocardial infarction |
|  |  | CIR011 | **2.52 (1.13 - 5.65)** | **3.43E-02** | **2.72 (1.27 - 5.82)** | **1.40E-02** | Coronary atherosclerosis and other heart disease |
|  |  | CIR020 | N/A | | **3.40 (1.07 - 10.81)** | **4.15E-02** | Cerebral infarction |
|  |  | CIR021 | 1.28 (0.55 - 2.98) | 6.15E-01 | **2.36 (1.24 - 4.50)** | **1.34E-02** | Acute hemorrhagic cerebrovascular disease |
|  |  | CIR024 | **5.71 (1.40 - 23.34)** | **2.37E-02** | **7.93 (2.36 - 26.62)** | **1.88E-03** | Other and ill-defined cerebrovascular disease |
|  |  | CIR026 | **4.62 (1.73 - 12.36)** | **5.86E-03** | **7.44 (3.35 - 16.52)** | **4.42E-06** | Peripheral and visceral vascular disease |
|  |  | DIG017 | N/A | | **10.40 (3.34 - 32.34)** | **1.83E-04** | Biliary tract disease |
|  |  | DIG025 | N/A | | **5.61 (1.96 - 16.05)** | **2.76E-03** | Other specified and unspecified gastrointestinal disorders |
|  |  | GEN004 | N/A | | **10.16 (3.66 - 28.21)** | **3.65E-05** | Urinary tract infections |
|  |  | NEO012 | 2.00 (0.76 - 5.21) | 2.01E-01 | 2.40 (0.99 - 5.81) | 5.40E-02 | Gastrointestinal cancers - esophagus |
|  |  | NEO015^5^ | **2.53 (1.27 - 5.05)** | **1.63E-02** | **2.77 (1.43 - 5.37)** | **3.99E-03** | Gastrointestinal cancers - colorectal |
|  |  | NEO018^5^ | **6.32 (2.39 - 16.69)** | **9.33E-04** | **8.65 (3.67 - 20.41)** | **4.42E-06** | Gastrointestinal cancers - bile duct |
|  |  | NEO025 | N/A | | **3.29 (1.16 - 9.39)** | **3.00E-02** | Skin cancers - melanoma |
|  |  | NEO048^5^ | **3.45 (1.32 - 9.01)** | **1.97E-02** | **3.30 (1.27 - 8.58)** | **1.90E-02** | Nervous system cancers - brain |
|  |  | NEO051 | 1.18 (0.57 - 2.42) | 6.58E-01 | **1.95 (1.09 - 3.51)** | **3.00E-02** | Endocrine system cancers - pancreas |
|  |  | NEO071^5^ | **10.14 (5.61 - 18.35)** | **2.66E-13** | **11.05 (6.26 - 19.51)** | **2.57E-15** | Malignant neoplasm, unspecified |
|  |  | NVS006 | **7.97 (2.11 - 30.08)** | **5.86E-03** | **7.97 (2.11 - 30.08)** | **3.81E-03** | Other specified hereditary and degenerative nervous system conditions |
|  |  | NVS011 | **4.56 (2.22 - 9.37)** | **2.61E-04** | **5.66 (2.95 - 10.85)** | **1.90E-06** | Neurocognitive disorders |
|  |  | RSP002 | **3.61 (1.57 - 8.31)** | **5.86E-03** | **4.35 (2.05 - 9.21)** | **3.21E-04** | Pneumonia (except that caused by tuberculosis) |
| Non-hospitalized COVID-19 vs. reference | Death with a prior history | CIR008 | N/A^4^ | | **7.60 (2.22 - 26.01)** | **9.79E-03** | Hypertension with complications and secondary hypertension |
|  |  | CIR009 | N/A | | 1.16 (0.47 - 2.85) | 8.45E-01 | Acute myocardial infarction |
|  |  | CIR011 | 0.75 (0.42 - 1.34) | 4.36E-01 | 1.19 (0.75 - 1.90) | 7.41E-01 | Coronary atherosclerosis and other heart disease |
|  |  | CIR019 | N/A | | 3.16 (1.03 - 9.67) | 1.16E-01 | Heart failure |
|  |  | EXT020 | **4.01 (1.41 - 11.45)** | **2.83E-02** | **3.26 (1.27 - 8.36)** | **4.53E-02** | External cause codes: intent of injury, accidental/unintentional |
|  |  | EXT029 | **4.31 (1.49 - 12.48)** | **2.83E-02** | **3.63 (1.39 - 9.46)** | **3.36E-02** | External cause codes: subsequent encounter |
|  |  | NEO015 | 0.88 (0.56 - 1.41) | 6.03E-01 | 1.10 (0.74 - 1.65) | 8.43E-01 | Gastrointestinal cancers - colorectal |
|  |  | NEO043 | N/A | | 1.85 (0.82 - 4.17) | 3.20E-01 | Urinary system cancers - bladder |
|  |  | NEO048 | N/A | | 0.91 (0.37 - 2.24) | 8.96E-01 | Nervous system cancers - brain |
|  |  | NEO051 | 0.82 (0.41 - 1.64) | 6.03E-01 | 1.11 (0.62 - 1.97) | 8.45E-01 | Endocrine system cancers - pancreas |
|  |  | NVS004 | 1.26 (0.79 - 2.01) | 4.36E-01 | 1.23 (0.80 - 1.91) | 6.11E-01 | Parkinson`s disease |
|  |  | NVS006 | N/A | | 1.36 (0.48 - 3.86) | 8.22E-01 | Other specified hereditary and degenerative nervous system conditions |
|  |  | NVS011 | **2.40 (1.67 - 3.46)** | **1.92E-05** | **2.65 (1.90 - 3.70)** | **1.79E-07** | Neurocognitive disorders |
|  |  | RSP008 | 0.63 (0.32 - 1.26) | 4.34E-01 | 0.99 (0.58 - 1.70) | 9.81E-01 | Chronic obstructive pulmonary disease and bronchiectasis |
|  |  | RSP016 | 1.60 (0.68 - 3.75) | 4.36E-01 | 1.74 (0.78 - 3.91) | 3.57E-01 | Other specified and unspecified lower respiratory disease |
|  | Death without a prior history | CIR009 | 0.43 (0.15 - 1.26) | 2.78E-01 | 0.61 (0.39 - 0.96) | 7.45E-02 | Acute myocardial infarction |
|  |  | CIR021 | 1.43 (0.84 - 2.43) | 3.33E-01 | 1.51 (0.91 - 2.51) | 1.50E-01 | Acute hemorrhagic cerebrovascular disease |
|  |  | CIR026 | 2.46 (0.85 - 7.10) | 2.78E-01 | **3.29 (1.36 - 7.95)** | **3.03E-02** | Peripheral and visceral vascular disease |
|  |  | DIG017 | N/A | | **4.44 (1.31 - 15.06)** | **4.58E-02** | Biliary tract disease |
|  |  | DIG025 | N/A | | **3.92 (1.46 - 10.51)** | **3.03E-02** | Other specified and unspecified gastrointestinal disorders |
|  |  | NEO015 | 0.58 (0.23 - 1.48) | 3.80E-01 | 0.57 (0.22 - 1.44) | 2.83E-01 | Gastrointestinal cancers - colorectal |
|  |  | NEO025 | 1.15 (0.42 - 3.12) | 8.83E-01 | 1.05 (0.39 - 2.81) | 9.28E-01 | Skin cancers - melanoma |
|  |  | NEO051 | 0.41 (0.19 - 0.90) | 1.17E-01 | 0.52 (0.26 - 1.04) | 9.88E-02 | Endocrine system cancers - pancreas |
|  |  | NEO071 | 1.02 (0.53 - 1.95) | 9.55E-01 | 1.15 (0.63 - 2.12) | 7.09E-01 | Malignant neoplasm, unspecified |
|  |  | NVS011 | **2.77 (1.43 - 5.38)** | **2.34E-02** | **3.04 (1.63 - 5.68)** | **5.26E-03** | Neurocognitive disorders |
| HR: hazard ratio; 95% CI represents the lower and upper 95% confidence interval of HR; p.adj^3^: FDR-adjusted p-value. CCSR: Clinical Classifications Software Refined, a method to classify disease categories.  1. Post-acute Mortality measures the risk of death by setting the follow-up start date as 30 days after the date of first positive SARS-CoV-2 test. 2. Overall Mortality measures the risk of death by setting the follow-up start date as the date of first positive SARS-CoV-2 test. 3. FDR correction was separately performed for each COVID-19 exposure cohort; Statistically significant associations (p.adj < 0.05) were labelled in bold face. 4. N/A indicates that the analysis for target outcome cannot proceed due to insufficient number (<5) of events in either the COVID-19 exposure or reference cohort.  5. Elevated neoplasm mortality, especially among hospitalized patients without prior cancer history, should be interpreted with caution, as this finding is more likely attributable to detection bias (unmasking of pre-existing undiagnosed malignancies) rather than representing a direct post-infection sequela of COVID-19. See Discussion for detailed interpretation. | | | | | | | |

| **Supplementary Table 5a. Differences in Associations between Post-acute Mortality Risks in Organ Systems and Overall COVID-19 in Comparisons of Subgroup (AF vs. AF-free).** | | | | | | | | | | |
| --- | --- | --- | --- | --- | --- | --- | --- | --- | --- | --- |
| **Outcome** | **HR(CI)_present_** | **p.adj_present_** | **Sig_present_** | **HR(CI)_absent_** | **p.adj_absent_** | **Sig_absent_** | **RHR** | **p.adj_RHR_** | **Comparison** | **Organ System Description** |
| All-cause mortality | 1.69  (1.50 - 1.89) | 2.69E-18 | *** | 1.43  (1.34 - 1.52) | 2.35E-29 | *** | 1.18 | 3.55E-02 | AF vs. AF-free |  |
| DIG | 1.85  (1.02 - 3.35) | 4.97E-02 | * | 1.33  (0.82 - 2.14) | 4.94E-01 |  | 1.39 | 6.34E-01 |  | Diseases of the digestive system |
| EXT | 5.05  (1.32 - 19.31) | 2.41E-02 | * | 3.36  (1.62 - 6.96) | 3.08E-03 | ** | 1.50 | 7.70E-01 |  | External cause codes |
| GEN | 3.23  (1.47 - 7.07) | 5.38E-03 | ** | 1.43  (0.67 - 3.06) | 5.65E-01 |  | 2.25 | 2.89E-01 |  | Diseases of the genitourinary system |
| INJ | 1.40  (0.47 - 4.14) | 5.45E-01 |  | 1.38  (0.60 - 3.17) | 5.98E-01 |  | 1.01 | 9.84E-01 |  | Injury poisoning and certain other consequences of external causes |
| NEO | 1.43  (1.13 - 1.80) | 5.29E-03 | ** | 1.35  (1.23 - 1.49) | 3.88E-09 | *** | 1.06 | 7.70E-01 |  | Neoplasms |
| NVS | 2.49  (2.04 - 3.03) | 8.44E-19 | *** | 1.12  (0.67 - 1.87) | 6.73E-01 |  | 2.23 | 2.33E-02 |  | Diseases of the nervous system |
| RSP | 1.89  (1.26 - 2.85) | 5.29E-03 | ** | 0.94  (0.70 - 1.25) | 6.73E-01 |  | 2.02 | 2.33E-02 |  | Diseases of the respiratory system |
| **Supplementary Table 5b. Differences in Associations between Post-acute Mortality Risks in Specific Disorders and Overall COVID-19 in Comparisons of Subgroup (AF vs. AF-free).** | | | | | | | | | | |
| **Outcome** | **HR(CI)_present_** | **p.adj_present_** | **Sig_present_** | **HR(CI)_absent_** | **p.adj_absent_** | **Sig_absent_** | **RHR** | **p.adj_RHR_** | **Comparison** | **CCSR Category Description** |
| CIR008 | 2.35  (0.82 - 6.71) | 2.11E-01 |  | 0.67  (0.27 - 1.68) | 7.88E-01 |  | 3.50 | 3.27E-01 | AF vs. AF-free | Hypertension with complications and secondary hypertension |
| CIR009 | 0.96  (0.52 - 1.75) | 8.84E-01 |  | 0.68  (0.44 - 1.05) | 2.69E-01 |  | 1.41 | 3.69E-01 |  | Acute myocardial infarction |
| CIR011 | 1.69  (0.99 - 2.88) | 1.33E-01 |  | 1.04  (0.69 - 1.56) | 8.82E-01 |  | 1.63 | 3.27E-01 |  | Coronary atherosclerosis and other heart disease |
| CIR021 | 1.62  (0.63 - 4.15) | 4.17E-01 |  | 0.94  (0.55 - 1.61) | 8.82E-01 |  | 1.72 | 3.69E-01 |  | Acute hemorrhagic cerebrovascular disease |
| GEN004 | 5.51  (1.77 - 17.17) | 3.24E-02 | * | 1.95  (0.65 - 5.81) | 5.82E-01 |  | 2.83 | 3.27E-01 |  | Urinary tract infections |
| NEO015 | 0.86  (0.32 - 2.27) | 8.43E-01 |  | 1.39  (1.02 - 1.90) | 1.89E-01 |  | 0.62 | 3.69E-01 |  | Gastrointestinal cancers - colorectal |
| NEO048 | 2.33  (0.79 - 6.88) | 2.11E-01 |  | 1.16  (0.66 - 2.02) | 8.62E-01 |  | 2.01 | 3.69E-01 |  | Nervous system cancers - brain |
| NVS011 | 1.38  (0.72 - 2.65) | 4.17E-01 |  | 2.58  (1.97 - 3.38) | 5.30E-11 | *** | 0.53 | 3.27E-01 |  | Neurocognitive disorders |
| RSP002 | 2.32  (1.09 - 4.97) | 1.33E-01 |  | 1.06  (0.52 - 2.14) | 8.82E-01 |  | 2.20 | 3.27E-01 |  | Pneumonia (except that caused by tuberculosis) |
| RSP008 | 1.83  (1.02 - 3.32) | 1.33E-01 |  | 1.15  (0.78 - 1.67) | 8.03E-01 |  | 1.60 | 3.27E-01 |  | Chronic obstructive pulmonary disease and bronchiectasis |
| **Supplementary Table 5c. Differences in Associations between Overall Mortality Risks in Organ Systems and Overall COVID-19 in Comparisons of Subgroup (AF vs. AF-free).** | | | | | | | | | | |
| **Outcome** | **HR(CI)_present_** | **p.adj_present_** | **Sig_present_** | **HR(CI)_absent_** | **p.adj_absent_** | **Sig_absent_** | **RHR** | **p.adj_RHR_** | **Comparison** | **CCSR Category Description** |
| All-cause mortality | 2.58  (2.35 - 2.84) | 1.56E-84 | *** | 2.31  (2.20 - 2.43) | 3.74E-237 | *** | 1.12 | 1.24E-01 | AF vs. AF-free |  |
| DIG | 2.11  (1.21 - 3.67) | 1.43E-02 | * | 1.97  (1.34 - 2.89) | 1.41E-03 | ** | 1.07 | 8.72E-01 |  | Diseases of the digestive system |
| EXT | 3.85  (1.09 - 13.64) | 5.23E-02 |  | 3.36  (1.71 - 6.61) | 1.41E-03 | ** | 1.15 | 8.72E-01 |  | External cause codes |
| GEN | 3.35  (1.59 - 7.05) | 2.90E-03 | ** | 2.06  (1.08 - 3.91) | 5.48E-02 |  | 1.63 | 7.27E-01 |  | Diseases of the genitourinary system |
| INF | 1.04  (0.40 - 2.70) | 9.35E-01 |  | 1.51  (0.95 - 2.39) | 1.38E-01 |  | 0.69 | 8.72E-01 |  | Certain infectious and parasitic diseases |
| INJ | 1.31  (0.49 - 3.48) | 6.59E-01 |  | 1.75  (0.84 - 3.67) | 1.71E-01 |  | 0.74 | 8.72E-01 |  | Injury poisoning and certain other consequences of external causes |
| MUS | 1.43  (0.40 - 5.11) | 6.59E-01 |  | 2.12  (0.86 - 5.20) | 1.46E-01 |  | 0.68 | 8.72E-01 |  | Diseases of the musculoskeletal system and connective tissue |
| NEO | 1.55  (1.25 - 1.91) | 1.48E-04 | *** | 1.52  (1.39 - 1.66) | 3.39E-19 | *** | 1.02 | 8.72E-01 |  | Neoplasms |
| NVS | 2.58  (2.14 - 3.11) | 9.77E-23 | *** | 1.23  (0.77 - 1.96) | 4.20E-01 |  | 2.10 | 2.10E-02 |  | Diseases of the nervous system |
| RSP | 2.19  (1.50 - 3.20) | 1.48E-04 | *** | 1.07  (0.82 - 1.39) | 6.40E-01 |  | 2.06 | 2.10E-02 |  | Diseases of the respiratory system |
| **Supplementary Table 5d. Differences in Associations between Overall Mortality Risks in Specific Disorders and Overall COVID-19 in Comparisons of Subgroup (AF vs. AF-free).** | | | | | | | | | | |
| **Outcome** | **HR(CI)_present_** | **p.adj_present_** | **Sig_present_** | **HR(CI)_absent_** | **p.adj_absent_** | **Sig_absent_** | **RHR** | **p.adj_RHR_** | **Comparison** | **CCSR Category Description** |
| CIR003 | 1.28  (0.36 - 4.60) | 8.07E-01 |  | 2.13  (0.82 - 5.51) | 2.76E-01 |  | 0.60 | 6.09E-01 | AF vs. AF-free | Nonrheumatic and unspecified valve disorders |
| CIR004 | 3.00  (0.94 - 9.60) | 9.36E-02 |  | 3.06  (1.14 - 8.18) | 6.90E-02 |  | 0.98 | 9.80E-01 |  | Endocarditis and endocardial disease |
| CIR008 | 3.11  (1.18 - 8.20) | 4.99E-02 | * | 0.73  (0.32 - 1.71) | 6.18E-01 |  | 4.24 | 1.90E-01 |  | Hypertension with complications and secondary hypertension |
| CIR009 | 0.93  (0.53 - 1.65) | 8.15E-01 |  | 0.95  (0.66 - 1.37) | 7.84E-01 |  | 0.98 | 9.80E-01 |  | Acute myocardial infarction |
| CIR011 | 2.12  (1.32 - 3.40) | 9.42E-03 | ** | 1.10  (0.74 - 1.63) | 6.83E-01 |  | 1.93 | 1.90E-01 |  | Coronary atherosclerosis and other heart disease |
| CIR019 | 5.12  (1.95 - 13.42) | 7.26E-03 | ** | 0.64  (0.22 - 1.89) | 6.09E-01 |  | 8.01 | 8.02E-02 |  | Heart failure |
| CIR021 | 1.74  (0.75 - 4.01) | 2.40E-01 |  | 1.18  (0.73 - 1.91) | 6.18E-01 |  | 1.47 | 5.76E-01 |  | Acute hemorrhagic cerebrovascular disease |
| CIR026 | 2.62  (0.84 - 8.18) | 1.30E-01 |  | 4.86  (2.37 - 9.94) | 8.09E-05 | *** | 0.54 | 5.76E-01 |  | Peripheral and visceral vascular disease |
| GEN004 | 5.51  (1.77 - 17.17) | 1.30E-02 | * | 3.43  (1.38 - 8.54) | 2.61E-02 | * | 1.61 | 6.09E-01 |  | Urinary tract infections |
| NEO015 | 1.11  (0.50 - 2.45) | 8.15E-01 |  | 1.59  (1.19 - 2.12) | 6.53E-03 | ** | 0.70 | 5.76E-01 |  | Gastrointestinal cancers - colorectal |
| NEO039 | 3.86  (1.78 - 8.37) | 7.26E-03 | ** | 2.25  (1.60 - 3.16) | 2.56E-05 | *** | 1.72 | 3.72E-01 |  | Male reproductive system cancers - prostate |
| NEO043 | 3.24  (1.03 - 10.19) | 8.19E-02 |  | 1.33  (0.77 - 2.31) | 4.92E-01 |  | 2.43 | 3.72E-01 |  | Urinary system cancers - bladder |
| NEO048 | 2.78  (0.98 - 7.89) | 8.66E-02 |  | 1.17  (0.69 - 1.98) | 6.44E-01 |  | 2.38 | 3.72E-01 |  | Nervous system cancers - brain |
| NVS011 | 1.80  (1.01 - 3.21) | 8.19E-02 |  | 2.71  (2.10 - 3.50) | 4.19E-13 | *** | 0.66 | 3.72E-01 |  | Neurocognitive disorders |
| RSP002 | 2.66  (1.29 - 5.49) | 2.18E-02 | * | 1.42  (0.78 - 2.59) | 4.49E-01 |  | 1.87 | 3.72E-01 |  | Pneumonia (except that caused by tuberculosis) |
| RSP008 | 2.13  (1.24 - 3.63) | 1.87E-02 | * | 1.25  (0.87 - 1.78) | 4.49E-01 |  | 1.71 | 3.72E-01 |  | Chronic obstructive pulmonary disease and bronchiectasis |
| HR(CI)_present_ : Hazard ratio and 95% confidence inteval for the risk factor present subgroup with the examined characteristics; p.adj_present_: FDR-adjusted p-value for the risk factor present subgroup; HR(CI)_absent_: Hazard ratio and 95% confidence inteval for the risk factor absent subgroup without the examined characteristics; p.adj_absent_: FDR-adjusted p-value for the risk factor absent subgroup; Sig_present_ and Sig_absent_: * indicates an FDR-adjusted p-value between 0.01 and 0.05, ** indicates between 0.001 and 0.01, and *** indicates a value smaller than 0.001 for the risk factor present and absent subgroups, respectively. RHR: Ratio of hazard ratios for comparisons between the risk factor present and absent subgroups. p.adj_RHR_*: FDR-adjusted p-value for comparisons between the risk factor present and absent subgroups, where a p.adj_RHR_ < 0.05 indicates significant differences.  CCSR: Clinical Classifications Software Refined, a method to classify disease categories. | | | | | | | | | | |

| **Supplementary Table 6a. Differences in Associations between Post-acute Mortality Risks in Organ Systems and Overall COVID-19 in Comparisons of Subgroup (CAD vs. CAD-free).** | | | | | | | | | | |
| --- | --- | --- | --- | --- | --- | --- | --- | --- | --- | --- |
| **Outcome** | **HR(CI)_present_** | **p.adj_present_** | **Sig_present_** | **HR(CI)_absent_** | **p.adj_absent_** | **Sig_absent_** | **RHR** | **p.adj_RHR_** | **Comparison** | **Organ System Description** |
| All-cause mortality | 1.46  (1.33 - 1.60) | 3.83E-14 | *** | 1.49  (1.39 - 1.59) | 4.43E-31 | *** | 0.98 | 8.25E-01 | CAD vs. CAD-free |  |
| DIG | 1.56  (0.89 - 2.73) | 1.56E-01 |  | 1.41  (0.87 - 2.31) | 2.65E-01 |  | 1.11 | 8.25E-01 |  | Diseases of the digestive system |
| GEN | 3.58  (1.50 - 8.54) | 8.59E-03 | ** | 1.69  (0.85 - 3.35) | 2.65E-01 |  | 2.12 | 5.25E-01 |  | Diseases of the genitourinary system |
| INF | 1.18  (0.54 - 2.59) | 6.73E-01 |  | 1.32  (0.74 - 2.38) | 4.68E-01 |  | 0.90 | 8.25E-01 |  | Certain infectious and parasitic diseases |
| INJ | 1.39  (0.48 - 4.03) | 6.21E-01 |  | 0.81  (0.34 - 1.91) | 6.27E-01 |  | 1.72 | 6.99E-01 |  | Injury poisoning and certain other consequences of external causes |
| NEO | 1.29  (1.09 - 1.54) | 8.59E-03 | ** | 1.45  (1.31 - 1.61) | 8.96E-12 | *** | 0.89 | 5.25E-01 |  | Neoplasms |
| NVS | 1.76  (1.19 - 2.60) | 8.59E-03 | ** | 2.40  (1.94 - 2.96) | 1.49E-15 | *** | 0.73 | 5.25E-01 |  | Diseases of the nervous system |
| RSP | 1.42  (1.01 - 2.02) | 7.39E-02 |  | 1.09  (0.78 - 1.50) | 6.27E-01 |  | 1.31 | 5.25E-01 |  | Diseases of the respiratory system |
| **Supplementary Table 6b. Differences in Associations between Post-acute Mortality Risks in Specific Disorders and Overall COVID-19 in Comparisons of Subgroup (CAD vs. CAD-free).** | | | | | | | | | | |
| **Outcome** | **HR(CI)_present_** | **p.adj_present_** | **Sig_present_** | **HR(CI)_absent_** | **p.adj_absent_** | **Sig_absent_** | **RHR** | **p.adj_RHR_** | **Comparison** | **CCSR Category Description** |
| CIR009 | 0.55  (0.30 - 0.99) | 2.34E-01 |  | 0.84  (0.54 - 1.32) | 6.36E-01 |  | 0.65 | 9.19E-01 | CAD vs. CAD-free | Acute myocardial infarction |
| CIR019 | 1.55  (0.52 - 4.64) | 6.48E-01 |  | 1.87  (0.74 - 4.70) | 4.56E-01 |  | 0.83 | 9.19E-01 |  | Heart failure |
| CIR021 | 1.30  (0.59 - 2.85) | 6.94E-01 |  | 1.17  (0.66 - 2.08) | 6.36E-01 |  | 1.11 | 9.19E-01 |  | Acute hemorrhagic cerebrovascular disease |
| GEN004 | 8.65  (2.14 - 34.94) | 3.69E-02 | * | 1.60  (0.62 - 4.12) | 5.46E-01 |  | 5.40 | 7.50E-01 |  | Urinary tract infections |
| NEO012 | 0.80  (0.31 - 2.12) | 7.61E-01 |  | 0.85  (0.48 - 1.51) | 6.36E-01 |  | 0.94 | 9.19E-01 |  | Gastrointestinal cancers - esophagus |
| NEO013 | 1.79  (0.60 - 5.32) | 4.91E-01 |  | 1.27  (0.53 - 3.02) | 6.36E-01 |  | 1.41 | 9.19E-01 |  | Gastrointestinal cancers - stomach |
| NEO015 | 1.17  (0.58 - 2.38) | 7.61E-01 |  | 1.47  (1.06 - 2.04) | 7.57E-02 |  | 0.80 | 9.19E-01 |  | Gastrointestinal cancers - colorectal |
| NEO025 | 2.49  (0.62 - 9.98) | 4.21E-01 |  | 1.58  (0.76 - 3.30) | 4.82E-01 |  | 1.58 | 9.19E-01 |  | Skin cancers - melanoma |
| NEO051 | 1.00  (0.43 - 2.35) | 9.93E-01 |  | 0.86  (0.57 - 1.30) | 6.36E-01 |  | 1.16 | 9.19E-01 |  | Endocrine system cancers - pancreas |
| NEO071 | 1.77  (0.61 - 5.10) | 4.91E-01 |  | 2.18  (1.31 - 3.64) | 1.37E-02 | * | 0.81 | 9.19E-01 |  | Malignant neoplasm, unspecified |
| NVS004 | 1.77  (0.91 - 3.41) | 3.42E-01 |  | 2.85  (1.93 - 4.20) | 8.75E-07 | *** | 0.62 | 9.19E-01 |  | Parkinson`s disease |
| NVS011 | 1.92  (1.11 - 3.31) | 1.44E-01 |  | 2.45  (1.85 - 3.25) | 8.53E-09 | *** | 0.78 | 9.19E-01 |  | Neurocognitive disorders |
| RSP002 | 1.82  (0.85 - 3.92) | 3.70E-01 |  | 1.66  (0.85 - 3.24) | 4.02E-01 |  | 1.10 | 9.19E-01 |  | Pneumonia (except that caused by tuberculosis) |
| RSP008 | 1.41  (0.88 - 2.25) | 3.74E-01 |  | 1.24  (0.82 - 1.89) | 5.46E-01 |  | 1.13 | 9.19E-01 |  | Chronic obstructive pulmonary disease and bronchiectasis |
| RSP016 | 0.88  (0.34 - 2.25) | 8.39E-01 |  | 0.80  (0.30 - 2.16) | 6.58E-01 |  | 1.10 | 9.19E-01 |  | Other specified and unspecified lower respiratory disease |
| **Supplementary Table 6c. Differences in Associations between Overall Mortality Risks in Organ Systems and Overall COVID-19 in Comparisons of Subgroup (CAD vs. CAD-free).** | | | | | | | | | | |
| **Outcome** | **HR(CI)_present_** | **p.adj_present_** | **Sig_present_** | **HR(CI)_absent_** | **p.adj_absent_** | **Sig_absent_** | **RHR** | **p.adj_RHR_** | **Comparison** | **CCSR Category Description** |
| All-cause mortality | 1.42  (1.16 - 1.74) | 2.10E-03 | ** | 1.36  (1.12 - 1.66) | 4.27E-03 | ** | 1.04 | 7.63E-01 | CAD vs. CAD-free |  |
| DIG | 1.76  (1.05 - 2.95) | 4.69E-02 | * | 2.04  (1.37 - 3.04) | 9.98E-04 | *** | 0.86 | 7.29E-01 |  | Diseases of the digestive system |
| EXT | 2.37  (0.74 - 7.59) | 1.87E-01 |  | 3.61  (1.80 - 7.22) | 8.80E-04 | *** | 0.66 | 6.94E-01 |  | External cause codes |
| GEN | 4.43  (2.07 - 9.51) | 5.88E-04 | *** | 1.78  (0.94 - 3.37) | 9.99E-02 |  | 2.49 | 4.89E-01 |  | Diseases of the genitourinary system |
| INF | 1.16  (0.55 - 2.42) | 7.03E-01 |  | 1.65  (1.00 - 2.73) | 7.71E-02 |  | 0.70 | 6.94E-01 |  | Certain infectious and parasitic diseases |
| INJ | 1.73  (0.69 - 4.34) | 2.70E-01 |  | 1.05  (0.48 - 2.28) | 9.08E-01 |  | 1.66 | 6.94E-01 |  | Injury poisoning and certain other consequences of external causes |
| NEO | 1.40  (1.19 - 1.64) | 4.09E-04 | *** | 1.63  (1.48 - 1.79) | 3.19E-22 | *** | 0.86 | 4.89E-01 |  | Neoplasms |
| NVS | 1.82  (1.27 - 2.61) | 2.61E-03 | ** | 2.45  (2.01 - 3.00) | 6.98E-18 | *** | 0.74 | 4.89E-01 |  | Diseases of the nervous system |
| RSP | 1.66  (1.20 - 2.30) | 3.75E-03 | ** | 1.24  (0.92 - 1.67) | 1.67E-01 |  | 1.34 | 4.89E-01 |  | Diseases of the respiratory system |
| **Supplementary Table 6d. Differences in Associations between Overall Mortality Risks in Specific Disorders and Overall COVID-19 in Comparisons of Subgroup (CAD vs. CAD-free).** | | | | | | | | | | |
| **Outcome** | **HR(CI)_present_** | **p.adj_present_** | **Sig_present_** | **HR(CI)_absent_** | **p.adj_absent_** | **Sig_absent_** | **RHR** | **p.adj_RHR_** | **Comparison** | **CCSR Category Description** |
| CIR003 | 1.62  (0.57 - 4.60) | 4.49E-01 |  | 2.49  (0.89 - 6.98) | 1.85E-01 |  | 0.65 | 8.95E-01 | CAD vs. CAD-free | Nonrheumatic and unspecified valve disorders |
| CIR004 | 10.05  (2.55 - 39.58) | 1.20E-02 | * | 1.50  (0.53 - 4.23) | 5.28E-01 |  | 6.70 | 2.88E-01 |  | Endocarditis and endocardial disease |
| CIR008 | 2.29  (0.85 - 6.11) | 1.92E-01 |  | 0.88  (0.39 - 1.99) | 8.20E-01 |  | 2.60 | 7.23E-01 |  | Hypertension with complications and secondary hypertension |
| CIR009 | 0.76  (0.47 - 1.24) | 3.80E-01 |  | 1.00  (0.67 - 1.51) | 9.88E-01 |  | 0.76 | 8.95E-01 |  | Acute myocardial infarction |
| CIR019 | 2.44  (0.95 - 6.30) | 1.47E-01 |  | 1.65  (0.67 - 4.05) | 3.82E-01 |  | 1.48 | 8.95E-01 |  | Heart failure |
| CIR021 | 1.77  (0.87 - 3.59) | 2.06E-01 |  | 1.27  (0.76 - 2.14) | 4.77E-01 |  | 1.39 | 8.95E-01 |  | Acute hemorrhagic cerebrovascular disease |
| CIR026 | 3.16  (1.02 - 9.84) | 1.25E-01 |  | 4.62  (2.23 - 9.56) | 2.34E-04 | *** | 0.68 | 8.95E-01 |  | Peripheral and visceral vascular disease |
| DIG017 | 9.77  (1.00 - 95.65) | 1.25E-01 |  | 2.74  (0.96 - 7.79) | 1.64E-01 |  | 3.57 | 8.95E-01 |  | Biliary tract disease |
| GEN004 | 12.38  (3.16 - 48.50) | 7.58E-03 | ** | 2.13  (0.89 - 5.09) | 1.85E-01 |  | 5.81 | 2.88E-01 |  | Urinary tract infections |
| INF003 | 2.23  (0.72 - 6.95) | 2.42E-01 |  | 2.38  (0.81 - 7.03) | 2.03E-01 |  | 0.94 | 9.67E-01 |  | Bacterial infections |
| NEO012 | 1.02  (0.44 - 2.35) | 9.71E-01 |  | 0.89  (0.52 - 1.53) | 7.58E-01 |  | 1.14 | 8.95E-01 |  | Gastrointestinal cancers - esophagus |
| NEO013 | 2.27  (0.87 - 5.94) | 1.92E-01 |  | 1.77  (0.86 - 3.64) | 2.03E-01 |  | 1.28 | 8.95E-01 |  | Gastrointestinal cancers - stomach |
| NEO015 | 1.60  (0.85 - 3.00) | 2.25E-01 |  | 1.57  (1.17 - 2.12) | 1.01E-02 | * | 1.01 | 9.67E-01 |  | Gastrointestinal cancers - colorectal |
| NEO018 | 4.87  (1.41 - 16.82) | 7.71E-02 |  | 1.73  (0.88 - 3.40) | 2.03E-01 |  | 2.82 | 7.23E-01 |  | Gastrointestinal cancers - bile duct |
| NEO025 | 4.09  (1.06 - 15.81) | 1.25E-01 |  | 1.62  (0.80 - 3.28) | 2.64E-01 |  | 2.53 | 8.68E-01 |  | Skin cancers - melanoma |
| NEO039 | 2.05  (1.12 - 3.78) | 9.36E-02 |  | 2.55  (1.76 - 3.69) | 5.42E-06 | *** | 0.81 | 8.95E-01 |  | Male reproductive system cancers - prostate |
| NEO043 | 0.62  (0.21 - 1.80) | 4.49E-01 |  | 2.56  (1.47 - 4.45) | 4.12E-03 | ** | 0.24 | 2.88E-01 |  | Urinary system cancers - bladder |
| NEO048 | 1.12  (0.39 - 3.19) | 9.04E-01 |  | 1.45  (0.86 - 2.42) | 2.52E-01 |  | 0.77 | 8.95E-01 |  | Nervous system cancers - brain |
| NEO051 | 1.05  (0.47 - 2.34) | 9.38E-01 |  | 1.15  (0.81 - 1.65) | 5.28E-01 |  | 0.91 | 9.10E-01 |  | Endocrine system cancers - pancreas |
| NEO071 | 1.77  (0.61 - 5.10) | 3.85E-01 |  | 2.23  (1.38 - 3.59) | 4.12E-03 | ** | 0.79 | 8.95E-01 |  | Malignant neoplasm, unspecified |
| NVS004 | 1.60  (0.86 - 2.98) | 2.25E-01 |  | 2.67  (1.85 - 3.85) | 2.09E-06 | *** | 0.60 | 7.23E-01 |  | Parkinson`s disease |
| NVS011 | 2.10  (1.26 - 3.51) | 3.66E-02 | * | 2.59  (1.98 - 3.39) | 1.05E-10 | *** | 0.81 | 8.95E-01 |  | Neurocognitive disorders |
| RSP002 | 2.28  (1.12 - 4.62) | 9.36E-02 |  | 1.79  (0.99 - 3.26) | 1.64E-01 |  | 1.27 | 8.95E-01 |  | Pneumonia (except that caused by tuberculosis) |
| RSP008 | 1.56  (1.01 - 2.42) | 1.25E-01 |  | 1.44  (0.98 - 2.11) | 1.67E-01 |  | 1.09 | 8.95E-01 |  | Chronic obstructive pulmonary disease and bronchiectasis |
| RSP016 | 0.81  (0.32 - 2.08) | 7.55E-01 |  | 1.13  (0.47 - 2.67) | 8.20E-01 |  | 0.72 | 8.95E-01 |  | Other specified and unspecified lower respiratory disease |
| HR(CI)_present_ : Hazard ratio and 95% confidence inteval for the risk factor present subgroup with the examined characteristics; p.adj_present_: FDR-adjusted p-value for the risk factor present subgroup; HR(CI)_absent_: Hazard ratio and 95% confidence inteval for the risk factor absent subgroup without the examined characteristics; p.adj_absent_: FDR-adjusted p-value for the risk factor absent subgroup; Sig_present_ and Sig_absent_: * indicates an FDR-adjusted p-value between 0.01 and 0.05, ** indicates between 0.001 and 0.01, and *** indicates a value smaller than 0.001 for the risk factor present and absent subgroups, respectively. RHR: Ratio of hazard ratios for comparisons between the risk factor present and absent subgroups. p.adj_RHR_*: FDR-adjusted p-value for comparisons between the risk factor present and absent subgroups, where a p.adj_RHR_ < 0.05 indicates significant differences.  CCSR: Clinical Classifications Software Refined, a method to classify disease categories. | | | | | | | | | | |

| **Supplementary Table 7a. Differences in Associations between Post-acute Mortality Risks in Organ Systems and Overall COVID-19 in Comparisons of Subgroup (CKD vs. CKD-free).** | | | | | | | | | | |
| --- | --- | --- | --- | --- | --- | --- | --- | --- | --- | --- |
| **Outcome** | **HR(CI)_present_** | **p.adj_present_** | **Sig_present_** | **HR(CI)_absent_** | **p.adj_absent_** | **Sig_absent_** | **RHR** | **p.adj_RHR_** | **Comparison** | **Organ System Description** |
| All-cause mortality | 1.80  (1.63 - 1.97) | 4.86E-33 | *** | 1.34  (1.26 - 1.43) | 2.13E-17 | *** | 1.34 | 6.31E-06 | CKD vs. CKD-free |  |
| CIR | 1.48  (1.16 - 1.87) | 2.80E-03 | ** | 0.98  (0.79 - 1.21) | 8.36E-01 |  | 1.51 | 6.47E-02 |  | Diseases of the circulatory system |
| DIG | 1.74  (0.96 - 3.13) | 1.06E-01 |  | 1.33  (0.83 - 2.13) | 3.19E-01 |  | 1.31 | 6.49E-01 |  | Diseases of the digestive system |
| INF | 1.18  (0.54 - 2.58) | 7.55E-01 |  | 1.30  (0.73 - 2.32) | 4.33E-01 |  | 0.91 | 8.64E-01 |  | Certain infectious and parasitic diseases |
| MUS | 1.88  (0.60 - 5.83) | 3.68E-01 |  | 2.15  (0.76 - 6.05) | 2.37E-01 |  | 0.87 | 8.64E-01 |  | Diseases of the musculoskeletal system and connective tissue |
| NEO | 1.61  (1.35 - 1.91) | 2.49E-07 | *** | 1.26  (1.13 - 1.40) | 5.27E-05 | *** | 1.28 | 6.47E-02 |  | Neoplasms |
| NVS | 2.60  (1.81 - 3.75) | 7.96E-07 | *** | 1.82  (1.47 - 2.27) | 2.57E-07 | *** | 1.43 | 2.01E-01 |  | Diseases of the nervous system |
| RSP | 1.06  (0.72 - 1.57) | 7.55E-01 |  | 1.37  (1.02 - 1.83) | 7.35E-02 |  | 0.78 | 5.02E-01 |  | Diseases of the respiratory system |
| **Supplementary Table 7b. Differences in Associations between Post-acute Mortality Risks in Specific Disorders and Overall COVID-19 in Comparisons of Subgroup (CKD vs. CKD-free).** | | | | | | | | | | |
| **Outcome** | **HR(CI)_present_** | **p.adj_present_** | **Sig_present_** | **HR(CI)_absent_** | **p.adj_absent_** | **Sig_absent_** | **RHR** | **p.adj_RHR_** | **Comparison** | **CCSR Category Description** |
| CIR008 | 3.97  (1.46 - 10.79) | 2.78E-02 | * | 0.59  (0.22 - 1.61) | 4.56E-01 |  | 6.67 | 1.36E-01 | CKD vs. CKD-free | Hypertension with complications and secondary hypertension |
| CIR009 | 0.77  (0.42 - 1.42) | 4.91E-01 |  | 0.69  (0.45 - 1.07) | 2.68E-01 |  | 1.11 | 9.05E-01 |  | Acute myocardial infarction |
| CIR011 | 1.53  (1.00 - 2.35) | 1.13E-01 |  | 1.15  (0.69 - 1.89) | 7.31E-01 |  | 1.34 | 5.43E-01 |  | Coronary atherosclerosis and other heart disease |
| CIR021 | 2.05  (0.86 - 4.89) | 1.87E-01 |  | 0.94  (0.54 - 1.63) | 9.30E-01 |  | 2.19 | 5.08E-01 |  | Acute hemorrhagic cerebrovascular disease |
| GEN004 | 5.72  (1.71 - 19.15) | 2.47E-02 | * | 2.04  (0.78 - 5.34) | 2.90E-01 |  | 2.80 | 5.08E-01 |  | Urinary tract infections |
| NEO013 | 3.31  (1.05 - 10.43) | 1.10E-01 |  | 0.94  (0.38 - 2.32) | 9.58E-01 |  | 3.51 | 4.91E-01 |  | Gastrointestinal cancers - stomach |
| NEO015 | 1.06  (0.58 - 1.96) | 8.41E-01 |  | 1.53  (1.09 - 2.14) | 7.58E-02 |  | 0.70 | 5.43E-01 |  | Gastrointestinal cancers - colorectal |
| NEO043 | 2.00  (0.64 - 6.22) | 3.33E-01 |  | 1.02  (0.53 - 1.95) | 9.63E-01 |  | 1.97 | 5.43E-01 |  | Urinary system cancers - bladder |
| NEO048 | 1.39  (0.56 - 3.47) | 5.08E-01 |  | 1.33  (0.74 - 2.39) | 4.56E-01 |  | 1.05 | 9.55E-01 |  | Nervous system cancers - brain |
| NEO051 | 1.49  (0.76 - 2.94) | 3.33E-01 |  | 0.74  (0.47 - 1.15) | 3.24E-01 |  | 2.03 | 4.91E-01 |  | Endocrine system cancers - pancreas |
| NEO071 | 2.67  (1.06 - 6.75) | 1.10E-01 |  | 1.70  (0.99 - 2.91) | 1.68E-01 |  | 1.57 | 5.43E-01 |  | Malignant neoplasm, unspecified |
| NVS004 | 3.74  (1.83 - 7.66) | 2.38E-03 | ** | 2.14  (1.44 - 3.16) | 1.20E-03 | ** | 1.75 | 5.08E-01 |  | Parkinson`s disease |
| NVS006 | 3.62  (0.94 - 13.93) | 1.23E-01 |  | 1.57  (0.65 - 3.83) | 4.56E-01 |  | 2.30 | 5.43E-01 |  | Other specified hereditary and degenerative nervous system conditions |
| NVS011 | 2.63  (1.59 - 4.35) | 2.38E-03 | ** | 2.01  (1.50 - 2.69) | 5.20E-05 | *** | 1.31 | 5.43E-01 |  | Neurocognitive disorders |
| RSP002 | 1.61  (0.73 - 3.56) | 3.33E-01 |  | 1.66  (0.87 - 3.16) | 2.83E-01 |  | 0.97 | 9.55E-01 |  | Pneumonia (except that caused by tuberculosis) |
| RSP008 | 1.21  (0.72 - 2.04) | 5.08E-01 |  | 1.55  (1.04 - 2.30) | 1.18E-01 |  | 0.78 | 5.62E-01 |  | Chronic obstructive pulmonary disease and bronchiectasis |
| **Supplementary Table 7c. Differences in Associations between Overall Mortality Risks in Organ Systems and Overall COVID-19 in Comparisons of Subgroup (CKD vs. CKD-free).** | | | | | | | | | | |
| **Outcome** | **HR(CI)_present_** | **p.adj_present_** | **Sig_present_** | **HR(CI)_absent_** | **p.adj_absent_** | **Sig_absent_** | **RHR** | **p.adj_RHR_** | **Comparison** | **CCSR Category Description** |
| All-cause mortality | 2.80  (2.59 - 3.02) | 5.37E-148 | *** | 2.17  (2.05 - 2.29) | 2.93E-172 | *** | 1.29 | 1.23E-06 | CKD vs. CKD-free |  |
| CIR | 1.71  (1.38 - 2.12) | 2.67E-06 | *** | 1.17  (0.97 - 1.42) | 1.32E-01 |  | 1.45 | 5.63E-02 |  | Diseases of the circulatory system |
| DIG | 2.26  (1.35 - 3.76) | 3.53E-03 | ** | 1.81  (1.21 - 2.70) | 6.23E-03 | ** | 1.25 | 6.93E-01 |  | Diseases of the digestive system |
| EXT | 3.83  (1.09 - 13.50) | 6.12E-02 |  | 3.21  (1.63 - 6.34) | 1.52E-03 | ** | 1.19 | 9.78E-01 |  | External cause codes |
| INF | 1.44  (0.73 - 2.84) | 3.72E-01 |  | 1.45  (0.86 - 2.46) | 1.66E-01 |  | 0.99 | 9.78E-01 |  | Certain infectious and parasitic diseases |
| INJ | 1.08  (0.40 - 2.91) | 8.77E-01 |  | 1.69  (0.81 - 3.54) | 1.66E-01 |  | 0.64 | 6.93E-01 |  | Injury poisoning and certain other consequences of external causes |
| MUS | 1.95  (0.68 - 5.59) | 3.07E-01 |  | 2.09  (0.79 - 5.52) | 1.66E-01 |  | 0.93 | 9.78E-01 |  | Diseases of the musculoskeletal system and connective tissue |
| NEO | 1.69  (1.44 - 1.98) | 5.41E-10 | *** | 1.42  (1.29 - 1.57) | 4.10E-12 | *** | 1.19 | 2.03E-01 |  | Neoplasms |
| NVS | 2.61  (1.85 - 3.69) | 1.80E-07 | *** | 1.98  (1.61 - 2.42) | 2.09E-10 | *** | 1.32 | 3.51E-01 |  | Diseases of the nervous system |
| RSP | 1.18  (0.81 - 1.70) | 4.26E-01 |  | 1.60  (1.22 - 2.08) | 1.46E-03 | ** | 0.74 | 3.51E-01 |  | Diseases of the respiratory system |
| **Supplementary Table 7d. Differences in Associations between Overall Mortality Risks in Specific Disorders and Overall COVID-19 in Comparisons of Subgroup (CKD vs. CKD-free).** | | | | | | | | | | |
| **Outcome** | **HR(CI)_present_** | **p.adj_present_** | **Sig_present_** | **HR(CI)_absent_** | **p.adj_absent_** | **Sig_absent_** | **RHR** | **p.adj_RHR_** | **Comparison** | **CCSR Category Description** |
| CIR003 | 1.50  (0.56 - 4.05) | 4.60E-01 |  | 2.58  (0.84 - 7.91) | 2.07E-01 |  | 0.58 | 5.93E-01 | CKD vs. CKD-free | Nonrheumatic and unspecified valve disorders |
| CIR008 | 4.59  (1.87 - 11.27) | 4.61E-03 | ** | 0.61  (0.23 - 1.63) | 4.56E-01 |  | 7.49 | 6.14E-02 |  | Hypertension with complications and secondary hypertension |
| CIR009 | 0.98  (0.59 - 1.63) | 9.34E-01 |  | 0.88  (0.59 - 1.30) | 6.67E-01 |  | 1.12 | 8.11E-01 |  | Acute myocardial infarction |
| CIR011 | 1.63  (1.10 - 2.42) | 3.00E-02 | * | 1.39  (0.88 - 2.21) | 2.55E-01 |  | 1.17 | 7.14E-01 |  | Coronary atherosclerosis and other heart disease |
| CIR021 | 2.33  (1.05 - 5.21) | 6.18E-02 |  | 1.15  (0.70 - 1.89) | 6.98E-01 |  | 2.03 | 4.54E-01 |  | Acute hemorrhagic cerebrovascular disease |
| DIG017 | 7.10  (1.67 - 30.16) | 1.92E-02 | * | 2.33  (0.72 - 7.52) | 2.55E-01 |  | 3.05 | 5.38E-01 |  | Biliary tract disease |
| GEN004 | 7.68  (2.44 - 24.20) | 3.51E-03 | ** | 2.65  (1.09 - 6.45) | 9.53E-02 |  | 2.90 | 4.54E-01 |  | Urinary tract infections |
| NEO012 | 0.61  (0.21 - 1.76) | 4.41E-01 |  | 1.09  (0.65 - 1.82) | 7.87E-01 |  | 0.56 | 5.38E-01 |  | Gastrointestinal cancers - esophagus |
| NEO013 | 4.11  (1.60 - 10.55) | 9.78E-03 | ** | 1.17  (0.52 - 2.59) | 7.82E-01 |  | 3.53 | 3.17E-01 |  | Gastrointestinal cancers - stomach |
| NEO015 | 1.25  (0.73 - 2.13) | 4.60E-01 |  | 1.76  (1.29 - 2.40) | 1.92E-03 | ** | 0.71 | 5.38E-01 |  | Gastrointestinal cancers - colorectal |
| NEO018 | 6.15  (1.60 - 23.65) | 1.92E-02 | * | 2.00  (1.04 - 3.81) | 9.58E-02 |  | 3.08 | 4.54E-01 |  | Gastrointestinal cancers - bile duct |
| NEO039 | 3.25  (1.74 - 6.06) | 2.15E-03 | ** | 2.25  (1.58 - 3.20) | 6.50E-05 | *** | 1.45 | 5.38E-01 |  | Male reproductive system cancers - prostate |
| NEO043 | 2.75  (1.10 - 6.85) | 5.30E-02 |  | 1.18  (0.64 - 2.15) | 6.98E-01 |  | 2.33 | 4.54E-01 |  | Urinary system cancers - bladder |
| NEO048 | 1.42  (0.59 - 3.44) | 4.60E-01 |  | 1.50  (0.87 - 2.58) | 2.55E-01 |  | 0.95 | 9.31E-01 |  | Nervous system cancers - brain |
| NEO051 | 1.52  (0.80 - 2.86) | 2.77E-01 |  | 1.03  (0.70 - 1.51) | 8.86E-01 |  | 1.48 | 5.38E-01 |  | Endocrine system cancers - pancreas |
| NEO071 | 3.07  (1.31 - 7.20) | 2.11E-02 | * | 1.66  (1.00 - 2.78) | 1.20E-01 |  | 1.84 | 5.38E-01 |  | Malignant neoplasm, unspecified |
| NVS004 | 2.85  (1.47 - 5.53) | 6.64E-03 | ** | 2.10  (1.46 - 3.03) | 4.85E-04 | *** | 1.36 | 5.59E-01 |  | Parkinson`s disease |
| NVS006 | 8.46  (2.28 - 31.45) | 6.02E-03 | ** | 1.57  (0.65 - 3.83) | 4.56E-01 |  | 5.38 | 3.17E-01 |  | Other specified hereditary and degenerative nervous system conditions |
| NVS011 | 2.81  (1.74 - 4.53) | 4.62E-04 | *** | 2.17  (1.65 - 2.87) | 8.75E-07 | *** | 1.29 | 5.42E-01 |  | Neurocognitive disorders |
| RSP002 | 1.88  (0.89 - 3.96) | 1.44E-01 |  | 1.96  (1.12 - 3.45) | 6.72E-02 |  | 0.96 | 9.31E-01 |  | Pneumonia (except that caused by tuberculosis) |
| RSP008 | 1.34  (0.82 - 2.20) | 3.14E-01 |  | 1.76  (1.22 - 2.53) | 1.01E-02 | * | 0.76 | 5.43E-01 |  | Chronic obstructive pulmonary disease and bronchiectasis |
| HR(CI)_present_ : Hazard ratio and 95% confidence inteval for the risk factor present subgroup with the examined characteristics; p.adj_present_: FDR-adjusted p-value for the risk factor present subgroup; HR(CI)_absent_: Hazard ratio and 95% confidence inteval for the risk factor absent subgroup without the examined characteristics; p.adj_absent_: FDR-adjusted p-value for the risk factor absent subgroup; Sig_present_ and Sig_absent_: * indicates an FDR-adjusted p-value between 0.01 and 0.05, ** indicates between 0.001 and 0.01, and *** indicates a value smaller than 0.001 for the risk factor present and absent subgroups, respectively. RHR: Ratio of hazard ratios for comparisons between the risk factor present and absent subgroups. p.adj_RHR_*: FDR-adjusted p-value for comparisons between the risk factor present and absent subgroups, where a p.adj_RHR_ < 0.05 indicates significant differences.  CCSR: Clinical Classifications Software Refined, a method to classify disease categories. | | | | | | | | | | |

| **Supplementary Table 8a. Differences in Associations between Post-acute Mortality Risks in Organ Systems and Overall COVID-19 in Comparisons of Subgroup (COPD vs. COPD-free).** | | | | | | | | | | |
| --- | --- | --- | --- | --- | --- | --- | --- | --- | --- | --- |
| **Outcome** | **HR(CI)_present_** | **p.adj_present_** | **Sig_present_** | **HR(CI)_absent_** | **p.adj_absent_** | **Sig_absent_** | **RHR** | **p.adj_RHR_** | **Comparison** | **Organ System Description** |
| All-cause mortality | 1.45  (1.28 - 1.63) | 2.11E-08 | *** | 1.53  (1.44 - 1.62) | 1.24E-41 | *** | 0.95 | 8.25E-01 | COPD vs. COPD-free |  |
| CIR | 1.47  (1.06 - 2.03) | 4.10E-02 | * | 1.17  (0.98 - 1.41) | 8.19E-02 |  | 1.25 | 8.25E-01 |  | Diseases of the circulatory system |
| DIG | 1.49  (0.55 - 4.06) | 4.31E-01 |  | 1.49  (1.00 - 2.22) | 5.81E-02 |  | 1.00 | 9.97E-01 |  | Diseases of the digestive system |
| EXT | 5.61  (1.47 - 21.47) | 4.10E-02 | * | 3.20  (1.55 - 6.64) | 3.05E-03 | ** | 1.75 | 8.25E-01 |  | External cause codes |
| GEN | 2.51  (0.77 - 8.16) | 1.47E-01 |  | 2.20  (1.20 - 4.01) | 1.49E-02 | * | 1.14 | 9.95E-01 |  | Diseases of the genitourinary system |
| NEO | 1.23  (0.99 - 1.52) | 8.58E-02 |  | 1.49  (1.35 - 1.64) | 6.17E-15 | *** | 0.83 | 7.85E-01 |  | Neoplasms |
| NVS | 2.31  (1.12 - 4.76) | 4.10E-02 | * | 2.15  (1.78 - 2.60) | 6.55E-15 | *** | 1.07 | 9.95E-01 |  | Diseases of the nervous system |
| **Supplementary Table 8b. Differences in Associations between Post-acute Mortality Risks in Specific Disorders and Overall COVID-19 in Comparisons of Subgroup (COPD vs. COPD-free).** | | | | | | | | | | |
| **Outcome** | **HR(CI)_present_** | **p.adj_present_** | **Sig_present_** | **HR(CI)_absent_** | **p.adj_absent_** | **Sig_absent_** | **RHR** | **p.adj_RHR_** | **Comparison** | **CCSR Category Description** |
| CIR009 | 0.82  (0.38 - 1.80) | 7.07E-01 |  | 0.70  (0.47 - 1.04) | 1.20E-01 |  | 1.18 | 8.68E-01 | COPD vs. COPD-free | Acute myocardial infarction |
| CIR011 | 1.52  (0.80 - 2.90) | 2.63E-01 |  | 1.21  (0.83 - 1.75) | 3.57E-01 |  | 1.26 | 8.68E-01 |  | Coronary atherosclerosis and other heart disease |
| CIR026 | 6.10  (1.43 - 25.95) | 4.32E-02 | * | 2.82  (1.30 - 6.10) | 1.52E-02 | * | 2.16 | 8.68E-01 |  | Peripheral and visceral vascular disease |
| EXT020 | 7.28  (1.70 - 31.24) | 3.41E-02 | * | 4.09  (1.85 - 9.01) | 2.17E-03 | ** | 1.78 | 8.68E-01 |  | External cause codes: intent of injury, accidental/unintentional |
| EXT029 | 7.28  (1.70 - 31.24) | 3.41E-02 | * | 3.49  (1.62 - 7.50) | 4.22E-03 | ** | 2.09 | 8.68E-01 |  | External cause codes: subsequent encounter |
| NEO015 | 0.38  (0.12 - 1.18) | 1.71E-01 |  | 1.54  (1.13 - 2.09) | 1.49E-02 | * | 0.25 | 1.83E-01 |  | Gastrointestinal cancers - colorectal |
| NEO051 | 0.97  (0.33 - 2.88) | 9.57E-01 |  | 0.85  (0.57 - 1.26) | 4.12E-01 |  | 1.15 | 8.68E-01 |  | Endocrine system cancers - pancreas |
| NVS011 | 2.60  (0.95 - 7.10) | 1.42E-01 |  | 2.38  (1.84 - 3.08) | 4.42E-10 | *** | 1.09 | 8.68E-01 |  | Neurocognitive disorders |
| RSP002 | 2.22  (0.82 - 6.05) | 1.77E-01 |  | 1.61  (0.89 - 2.89) | 1.44E-01 |  | 1.38 | 8.68E-01 |  | Pneumonia (except that caused by tuberculosis) |
| **Supplementary Table 8c. Differences in Associations between Overall Mortality Risks in Organ Systems and Overall COVID-19 in Comparisons of Subgroup (COPD vs. COPD-free).** | | | | | | | | | | |
| **Outcome** | **HR(CI)_present_** | **p.adj_present_** | **Sig_present_** | **HR(CI)_absent_** | **p.adj_absent_** | **Sig_absent_** | **RHR** | **p.adj_RHR_** | **Comparison** | **CCSR Category Description** |
| All-cause mortality | 2.42  (2.19 - 2.66) | 1.54E-69 | *** | 2.41  (2.30 - 2.54) | 4.41E-262 | *** | 1.00 | 9.82E-01 | COPD vs. COPD-free |  |
| CIR | 1.64  (1.23 - 2.21) | 1.59E-03 | ** | 1.42  (1.20 - 1.67) | 3.58E-05 | *** | 1.16 | 7.67E-01 |  | Diseases of the circulatory system |
| DIG | 1.59  (0.62 - 4.03) | 3.32E-01 |  | 2.07  (1.48 - 2.90) | 3.58E-05 | *** | 0.77 | 9.58E-01 |  | Diseases of the digestive system |
| EXT | 6.73  (1.85 - 24.44) | 5.30E-03 | ** | 2.71  (1.37 - 5.38) | 4.19E-03 | ** | 2.48 | 7.67E-01 |  | External cause codes |
| GEN | 2.50  (0.91 - 6.85) | 8.76E-02 |  | 2.43  (1.40 - 4.22) | 1.93E-03 | ** | 1.03 | 9.82E-01 |  | Diseases of the genitourinary system |
| NEO | 1.43  (1.18 - 1.73) | 9.82E-04 | *** | 1.64  (1.50 - 1.79) | 6.20E-26 | *** | 0.87 | 7.67E-01 |  | Neoplasms |
| NVS | 2.99  (1.63 - 5.49) | 9.82E-04 | *** | 2.18  (1.82 - 2.61) | 7.98E-17 | *** | 1.37 | 7.67E-01 |  | Diseases of the nervous system |
| **Supplementary Table 8d. Differences in Associations between Overall Mortality Risks in Specific Disorders and Overall COVID-19 in Comparisons of Subgroup (COPD vs. COPD-free).** | | | | | | | | | | |
| **Outcome** | **HR(CI)_present_** | **p.adj_present_** | **Sig_present_** | **HR(CI)_absent_** | **p.adj_absent_** | **Sig_absent_** | **RHR** | **p.adj_RHR_** | **Comparison** | **CCSR Category Description** |
| CIR008 | 5.55  (1.38 - 22.29) | 3.61E-02 | * | 1.15  (0.57 - 2.30) | 7.03E-01 |  | 4.84 | 4.00E-01 | COPD vs. COPD-free | Hypertension with complications and secondary hypertension |
| CIR009 | 1.25  (0.64 - 2.44) | 5.86E-01 |  | 0.88  (0.62 - 1.26) | 5.66E-01 |  | 1.41 | 6.33E-01 |  | Acute myocardial infarction |
| CIR011 | 1.91  (1.06 - 3.46) | 5.23E-02 |  | 1.33  (0.94 - 1.87) | 1.57E-01 |  | 1.44 | 6.33E-01 |  | Coronary atherosclerosis and other heart disease |
| CIR026 | 6.10  (1.43 - 25.95) | 3.61E-02 | * | 3.35  (1.69 - 6.67) | 1.78E-03 | ** | 1.82 | 6.58E-01 |  | Peripheral and visceral vascular disease |
| EXT020 | 7.28  (1.70 - 31.24) | 2.42E-02 | * | 3.54  (1.69 - 7.42) | 2.14E-03 | ** | 2.06 | 6.33E-01 |  | External cause codes: intent of injury, accidental/unintentional |
| EXT029 | 7.28  (1.70 - 31.24) | 2.42E-02 | * | 3.11  (1.51 - 6.40) | 4.08E-03 | ** | 2.34 | 6.33E-01 |  | External cause codes: subsequent encounter |
| NEO013 | 3.06  (1.10 - 8.52) | 5.23E-02 |  | 1.52  (0.75 - 3.09) | 3.04E-01 |  | 2.01 | 6.33E-01 |  | Gastrointestinal cancers - stomach |
| NEO015 | 0.50  (0.18 - 1.40) | 2.49E-01 |  | 1.72  (1.30 - 2.27) | 6.74E-04 | *** | 0.29 | 4.00E-01 |  | Gastrointestinal cancers - colorectal |
| NEO039 | 4.89  (1.79 - 13.35) | 2.42E-02 | * | 2.20  (1.58 - 3.08) | 3.02E-05 | *** | 2.22 | 6.33E-01 |  | Male reproductive system cancers - prostate |
| NEO043 | 1.76  (0.55 - 5.63) | 4.16E-01 |  | 1.69  (1.00 - 2.87) | 8.06E-02 |  | 1.04 | 9.51E-01 |  | Urinary system cancers - bladder |
| NEO048 | 1.21  (0.28 - 5.18) | 8.53E-01 |  | 1.44  (0.87 - 2.39) | 2.15E-01 |  | 0.84 | 9.34E-01 |  | Nervous system cancers - brain |
| NEO051 | 1.02  (0.37 - 2.82) | 9.68E-01 |  | 1.10  (0.78 - 1.56) | 6.20E-01 |  | 0.93 | 9.44E-01 |  | Endocrine system cancers - pancreas |
| NVS004 | 5.02  (1.17 - 21.45) | 5.23E-02 |  | 2.12  (1.53 - 2.93) | 3.48E-05 | *** | 2.37 | 6.33E-01 |  | Parkinson`s disease |
| NVS011 | 3.60  (1.53 - 8.49) | 2.42E-02 | * | 2.48  (1.94 - 3.17) | 9.82E-12 | *** | 1.45 | 6.33E-01 |  | Neurocognitive disorders |
| RSP002 | 2.43  (0.93 - 6.36) | 1.04E-01 |  | 1.89  (1.13 - 3.18) | 2.76E-02 | * | 1.28 | 7.99E-01 |  | Pneumonia (except that caused by tuberculosis) |
| HR(CI)_present_ : Hazard ratio and 95% confidence inteval for the risk factor present subgroup with the examined characteristics; p.adj_present_: FDR-adjusted p-value for the risk factor present subgroup; HR(CI)_absent_: Hazard ratio and 95% confidence inteval for the risk factor absent subgroup without the examined characteristics; p.adj_absent_: FDR-adjusted p-value for the risk factor absent subgroup; Sig_present_ and Sig_absent_: * indicates an FDR-adjusted p-value between 0.01 and 0.05, ** indicates between 0.001 and 0.01, and *** indicates a value smaller than 0.001 for the risk factor present and absent subgroups, respectively. RHR: Ratio of hazard ratios for comparisons between the risk factor present and absent subgroups. p.adj_RHR_*: FDR-adjusted p-value for comparisons between the risk factor present and absent subgroups, where a p.adj_RHR_ < 0.05 indicates significant differences.  CCSR: Clinical Classifications Software Refined, a method to classify disease categories. | | | | | | | | | | |

| **Supplementary Table 9a. Differences in Associations between Post-acute Mortality Risks in Organ Systems and Overall COVID-19 in Comparisons of Subgroup (dementia vs. dementia-free).** | | | | | | | | | | |
| --- | --- | --- | --- | --- | --- | --- | --- | --- | --- | --- |
| **Outcome** | **HR(CI)_present_** | **p.adj_present_** | **Sig_present_** | **HR(CI)_absent_** | **p.adj_absent_** | **Sig_absent_** | **RHR** | **p.adj_RHR_** | **Comparison** | **Organ System Description** |
| All-cause mortality | 1.56  (1.38 - 1.77) | 1.87E-11 | *** | 1.50  (1.41 - 1.59) | 9.99E-39 | *** | 1.04 | 8.10E-01 | dementia vs. dementia-free |  |
| CIR | 1.75  (0.75 - 4.08) | 3.96E-01 |  | 1.56  (1.04 - 2.34) | 4.98E-02 | * | 1.12 | 8.10E-01 |  | Diseases of the circulatory system |
| EXT | 5.91  (1.52 - 22.97) | 3.12E-02 | * | 2.65  (1.24 - 5.66) | 2.40E-02 | * | 2.23 | 8.08E-01 |  | External cause codes |
| INF | 0.92  (0.28 - 3.02) | 9.63E-01 |  | 1.60  (0.97 - 2.63) | 7.93E-02 |  | 0.58 | 8.08E-01 |  | Certain infectious and parasitic diseases |
| NEO | 1.01  (0.68 - 1.50) | 9.63E-01 |  | 1.42  (1.30 - 1.56) | 1.82E-13 | *** | 0.71 | 5.79E-01 |  | Neoplasms |
| RSP | 1.37  (0.73 - 2.54) | 4.87E-01 |  | 1.20  (0.93 - 1.55) | 1.52E-01 |  | 1.14 | 8.10E-01 |  | Diseases of the respiratory system |
| **Supplementary Table 9b. Differences in Associations between Post-acute Mortality Risks in Specific Disorders and Overall COVID-19 in Comparisons of Subgroup (dementia vs. dementia-free).** | | | | | | | | | | |
| **Outcome** | **HR(CI)_present_** | **p.adj_present_** | **Sig_present_** | **HR(CI)_absent_** | **p.adj_absent_** | **Sig_absent_** | **RHR** | **p.adj_RHR_** | **Comparison** | **CCSR Category Description** |
| CIR009 | 1.71  (0.64 - 4.62) | 5.00E-01 |  | 0.70  (0.48 - 1.03) | 7.86E-02 |  | 2.45 | 4.55E-01 | dementia vs. dementia-free | Acute myocardial infarction |
| CIR011 | 1.34  (0.50 - 3.59) | 7.39E-01 |  | 1.27  (0.91 - 1.79) | 1.65E-01 |  | 1.05 | 9.66E-01 |  | Coronary atherosclerosis and other heart disease |
| EXT020 | 5.95  (1.53 - 23.15) | 2.37E-02 | * | 3.48  (1.51 - 8.01) | 1.18E-02 | * | 1.71 | 7.14E-01 |  | External cause codes: intent of injury, accidental/unintentional |
| EXT029 | 5.95  (1.53 - 23.15) | 2.37E-02 | * | 2.95  (1.31 - 6.62) | 1.73E-02 | * | 2.02 | 6.72E-01 |  | External cause codes: subsequent encounter |
| NVS004 | 2.38  (1.41 - 4.03) | 8.54E-03 | ** | 2.42  (1.58 - 3.71) | 3.59E-04 | *** | 0.99 | 9.66E-01 |  | Parkinson`s disease |
| NVS006 | 1.05  (0.30 - 3.62) | 9.43E-01 |  | 3.16  (1.32 - 7.58) | 1.73E-02 | * | 0.33 | 4.55E-01 |  | Other specified hereditary and degenerative nervous system conditions |
| RSP008 | 0.80  (0.32 - 2.01) | 7.39E-01 |  | 1.53  (1.10 - 2.13) | 1.73E-02 | * | 0.52 | 4.55E-01 |  | Chronic obstructive pulmonary disease and bronchiectasis |
| **Supplementary Table 9c. Differences in Associations between Overall Mortality Risks in Organ Systems and Overall COVID-19 in Comparisons of Subgroup (dementia vs. dementia-free).** | | | | | | | | | | |
| **Outcome** | **HR(CI)_present_** | **p.adj_present_** | **Sig_present_** | **HR(CI)_absent_** | **p.adj_absent_** | **Sig_absent_** | **RHR** | **p.adj_RHR_** | **Comparison** | **CCSR Category Description** |
| All-cause mortality | 2.28  (2.05 - 2.54) | 5.20E-50 | *** | 2.42  (2.31 - 2.54) | 2.08E-277 | *** | 0.94 | 5.11E-01 | dementia vs. dementia-free |  |
| CIR | 1.35  (0.87 - 2.11) | 2.94E-01 |  | 1.40  (1.21 - 1.62) | 2.05E-05 | *** | 0.97 | 9.38E-01 |  | Diseases of the circulatory system |
| DIG | 2.13  (0.96 - 4.74) | 1.48E-01 |  | 2.06  (1.46 - 2.90) | 5.77E-05 | *** | 1.03 | 9.38E-01 |  | Diseases of the digestive system |
| EXT | 5.91  (1.52 - 22.97) | 3.64E-02 | * | 2.50  (1.25 - 5.00) | 9.86E-03 | ** | 2.37 | 5.11E-01 |  | External cause codes |
| INF | 0.75  (0.24 - 2.36) | 7.15E-01 |  | 1.90  (1.22 - 2.97) | 5.98E-03 | ** | 0.39 | 3.65E-01 |  | Certain infectious and parasitic diseases |
| NEO | 1.07  (0.74 - 1.54) | 7.15E-01 |  | 1.59  (1.46 - 1.73) | 2.17E-26 | *** | 0.67 | 3.15E-01 |  | Neoplasms |
| RSP | 1.46  (0.81 - 2.64) | 2.94E-01 |  | 1.39  (1.10 - 1.76) | 5.98E-03 | ** | 1.05 | 9.38E-01 |  | Diseases of the respiratory system |
| **Supplementary Table 9d. Differences in Associations between Overall Mortality Risks in Specific Disorders and Overall COVID-19 in Comparisons of Subgroup (dementia vs. dementia-free).** | | | | | | | | | | |
| **Outcome** | **HR(CI)_present_** | **p.adj_present_** | **Sig_present_** | **HR(CI)_absent_** | **p.adj_absent_** | **Sig_absent_** | **RHR** | **p.adj_RHR_** | **Comparison** | **CCSR Category Description** |
| CIR009 | 1.43  (0.57 - 3.60) | 6.65E-01 |  | 0.89  (0.64 - 1.24) | 4.93E-01 |  | 1.61 | 5.65E-01 | dementia vs. dementia-free | Acute myocardial infarction |
| CIR011 | 1.34  (0.50 - 3.59) | 7.12E-01 |  | 1.46  (1.07 - 2.00) | 2.10E-02 | * | 0.91 | 9.11E-01 |  | Coronary atherosclerosis and other heart disease |
| EXT020 | 5.95  (1.53 - 23.15) | 2.28E-02 | * | 3.02  (1.39 - 6.57) | 1.22E-02 | * | 1.97 | 5.65E-01 |  | External cause codes: intent of injury, accidental/unintentional |
| EXT029 | 5.95  (1.53 - 23.15) | 2.28E-02 | * | 2.61  (1.22 - 5.58) | 2.01E-02 | * | 2.28 | 5.65E-01 |  | External cause codes: subsequent encounter |
| NVS004 | 2.25  (1.36 - 3.73) | 1.54E-02 | * | 2.17  (1.45 - 3.25) | 1.57E-03 | ** | 1.04 | 9.11E-01 |  | Parkinson`s disease |
| NVS006 | 1.05  (0.30 - 3.62) | 9.43E-01 |  | 3.84  (1.67 - 8.79) | 4.44E-03 | ** | 0.27 | 4.09E-01 |  | Other specified hereditary and degenerative nervous system conditions |
| RSP002 | 2.65  (0.80 - 8.83) | 2.01E-01 |  | 1.64  (0.99 - 2.72) | 5.90E-02 |  | 1.61 | 5.89E-01 |  | Pneumonia (except that caused by tuberculosis) |
| RSP008 | 0.80  (0.32 - 2.01) | 7.12E-01 |  | 1.72  (1.26 - 2.33) | 2.40E-03 | ** | 0.47 | 4.09E-01 |  | Chronic obstructive pulmonary disease and bronchiectasis |
| HR(CI)_present_ : Hazard ratio and 95% confidence inteval for the risk factor present subgroup with the examined characteristics; p.adj_present_: FDR-adjusted p-value for the risk factor present subgroup; HR(CI)_absent_: Hazard ratio and 95% confidence inteval for the risk factor absent subgroup without the examined characteristics; p.adj_absent_: FDR-adjusted p-value for the risk factor absent subgroup; Sig_present_ and Sig_absent_: * indicates an FDR-adjusted p-value between 0.01 and 0.05, ** indicates between 0.001 and 0.01, and *** indicates a value smaller than 0.001 for the risk factor present and absent subgroups, respectively. RHR: Ratio of hazard ratios for comparisons between the risk factor present and absent subgroups. p.adj_RHR_*: FDR-adjusted p-value for comparisons between the risk factor present and absent subgroups, where a p.adj_RHR_ < 0.05 indicates significant differences.  CCSR: Clinical Classifications Software Refined, a method to classify disease categories. | | | | | | | | | | |

| **Supplementary Table 10a. Differences in Associations between Post-acute Mortality Risks in Organ Systems and Overall COVID-19 in Comparisons of Subgroup (Heart failure vs. Heart failure-free).** | | | | | | | | | | |
| --- | --- | --- | --- | --- | --- | --- | --- | --- | --- | --- |
| **Outcome** | **HR(CI)_present_** | **p.adj_present_** | **Sig_present_** | **HR(CI)_absent_** | **p.adj_absent_** | **Sig_absent_** | **RHR** | **p.adj_RHR_** | **Comparison** | **Organ System Description** |
| All-cause mortality | 1.42  (1.26 - 1.61) | 1.21E-07 | *** | 1.53  (1.44 - 1.63) | 1.98E-42 | *** | 0.93 | 7.90E-01 | Heart failure vs. Heart failure-free |  |
| DIG | 1.69  (0.75 - 3.80) | 2.72E-01 |  | 1.40  (0.92 - 2.11) | 1.50E-01 |  | 1.21 | 8.77E-01 |  | Diseases of the digestive system |
| EXT | 6.44  (1.51 - 27.50) | 3.87E-02 | * | 3.19  (1.55 - 6.56) | 3.28E-03 | ** | 2.02 | 7.91E-01 |  | External cause codes |
| GEN | 1.86  (0.77 - 4.50) | 2.72E-01 |  | 2.51  (1.31 - 4.80) | 8.72E-03 | ** | 0.74 | 8.77E-01 |  | Diseases of the genitourinary system |
| INF | 1.31  (0.53 - 3.27) | 5.60E-01 |  | 1.23  (0.72 - 2.12) | 5.03E-01 |  | 1.06 | 9.09E-01 |  | Certain infectious and parasitic diseases |
| NEO | 1.13  (0.88 - 1.45) | 3.77E-01 |  | 1.42  (1.29 - 1.56) | 3.00E-12 | *** | 0.80 | 7.90E-01 |  | Neoplasms |
| NVS | 1.94  (1.14 - 3.30) | 3.87E-02 | * | 2.11  (1.74 - 2.57) | 3.04E-13 | *** | 0.92 | 8.77E-01 |  | Diseases of the nervous system |
| RSP | 1.44  (0.97 - 2.13) | 1.45E-01 |  | 1.11  (0.82 - 1.48) | 5.03E-01 |  | 1.30 | 7.90E-01 |  | Diseases of the respiratory system |
| **Supplementary Table 10b. Differences in Associations between Post-acute Mortality Risks in Specific Disorders and Overall COVID-19 in Comparisons of Subgroup (Heart failure vs. Heart failure-free).** | | | | | | | | | | |
| **Outcome** | **HR(CI)_present_** | **p.adj_present_** | **Sig_present_** | **HR(CI)_absent_** | **p.adj_absent_** | **Sig_absent_** | **RHR** | **p.adj_RHR_** | **Comparison** | **CCSR Category Description** |
| CIR008 | 2.55  (0.76 - 8.50) | 2.90E-01 |  | 1.13  (0.51 - 2.50) | 7.64E-01 |  | 2.25 | 5.39E-01 | Heart failure vs. Heart failure-free | Hypertension with complications and secondary hypertension |
| CIR009 | 0.65  (0.33 - 1.28) | 3.79E-01 |  | 0.87  (0.58 - 1.32) | 5.78E-01 |  | 0.75 | 7.02E-01 |  | Acute myocardial infarction |
| CIR011 | 0.93  (0.57 - 1.52) | 8.13E-01 |  | 1.57  (1.02 - 2.43) | 7.24E-02 |  | 0.59 | 5.12E-01 |  | Coronary atherosclerosis and other heart disease |
| GEN004 | 1.50  (0.39 - 5.73) | 7.22E-01 |  | 4.28  (1.65 - 11.11) | 8.59E-03 | ** | 0.35 | 5.39E-01 |  | Urinary tract infections |
| NEO015 | 1.10  (0.49 - 2.48) | 8.13E-01 |  | 1.40  (1.02 - 1.92) | 7.24E-02 |  | 0.79 | 7.63E-01 |  | Gastrointestinal cancers - colorectal |
| NVS004 | 6.66  (2.73 - 16.26) | 2.81E-04 | *** | 1.99  (1.37 - 2.88) | 1.21E-03 | ** | 3.35 | 1.28E-01 |  | Parkinson`s disease |
| NVS011 | 2.14  (0.94 - 4.88) | 2.11E-01 |  | 2.41  (1.85 - 3.13) | 4.14E-10 | *** | 0.89 | 8.31E-01 |  | Neurocognitive disorders |
| RSP002 | 1.34  (0.50 - 3.64) | 7.22E-01 |  | 1.52  (0.84 - 2.75) | 2.43E-01 |  | 0.88 | 8.31E-01 |  | Pneumonia (except that caused by tuberculosis) |
| RSP008 | 1.67  (1.03 - 2.70) | 1.73E-01 |  | 1.19  (0.78 - 1.81) | 5.54E-01 |  | 1.41 | 5.39E-01 |  | Chronic obstructive pulmonary disease and bronchiectasis |
| **Supplementary Table 10c. Differences in Associations between Overall Mortality Risks in Organ Systems and Overall COVID-19 in Comparisons of Subgroup (Heart failure vs. Heart failure-free).** | | | | | | | | | | |
| **Outcome** | **HR(CI)_present_** | **p.adj_present_** | **Sig_present_** | **HR(CI)_absent_** | **p.adj_absent_** | **Sig_absent_** | **RHR** | **p.adj_RHR_** | **Comparison** | **CCSR Category Description** |
| All-cause mortality | 2.35  (2.13 - 2.60) | 6.45E-64 | *** | 2.41  (2.30 - 2.53) | 4.43E-266 | *** | 0.97 | 7.22E-01 | Heart failure vs. Heart failure-free |  |
| DIG | 2.18  (1.14 - 4.18) | 3.43E-02 | * | 1.81  (1.26 - 2.60) | 2.19E-03 | ** | 1.20 | 7.22E-01 |  | Diseases of the digestive system |
| EXT | 6.30  (1.81 - 21.91) | 9.00E-03 | ** | 2.70  (1.34 - 5.44) | 8.20E-03 | ** | 2.33 | 4.92E-01 |  | External cause codes |
| GEN | 2.00  (0.88 - 4.55) | 1.26E-01 |  | 2.89  (1.60 - 5.21) | 9.26E-04 | *** | 0.69 | 7.22E-01 |  | Diseases of the genitourinary system |
| INF | 1.39  (0.61 - 3.17) | 4.55E-01 |  | 1.44  (0.89 - 2.33) | 1.40E-01 |  | 0.96 | 9.39E-01 |  | Certain infectious and parasitic diseases |
| INJ | 0.69  (0.27 - 1.81) | 4.55E-01 |  | 2.48  (1.14 - 5.39) | 2.90E-02 | * | 0.28 | 4.37E-01 |  | Injury poisoning and certain other consequences of external causes |
| NEO | 1.27  (1.01 - 1.60) | 5.97E-02 |  | 1.57  (1.44 - 1.71) | 7.17E-23 | *** | 0.81 | 4.73E-01 |  | Neoplasms |
| NVS | 2.52  (1.50 - 4.24) | 2.18E-03 | ** | 2.16  (1.80 - 2.60) | 1.01E-15 | *** | 1.17 | 7.22E-01 |  | Diseases of the nervous system |
| RSP | 1.70  (1.18 - 2.44) | 9.00E-03 | ** | 1.23  (0.93 - 1.61) | 1.40E-01 |  | 1.39 | 4.92E-01 |  | Diseases of the respiratory system |
| **Supplementary Table 10d. Differences in Associations between Overall Mortality Risks in Specific Disorders and Overall COVID-19 in Comparisons of Subgroup (Heart failure vs. Heart failure-free).** | | | | | | | | | | |
| **Outcome** | **HR(CI)_present_** | **p.adj_present_** | **Sig_present_** | **HR(CI)_absent_** | **p.adj_absent_** | **Sig_absent_** | **RHR** | **p.adj_RHR_** | **Comparison** | **CCSR Category Description** |
| CIR008 | 3.96  (1.35 - 11.64) | 3.39E-02 | * | 0.89  (0.40 - 1.96) | 7.88E-01 |  | 4.46 | 2.41E-01 | Heart failure vs. Heart failure-free | Hypertension with complications and secondary hypertension |
| CIR009 | 0.79  (0.45 - 1.38) | 5.46E-01 |  | 1.05  (0.73 - 1.52) | 7.88E-01 |  | 0.75 | 6.92E-01 |  | Acute myocardial infarction |
| CIR011 | 1.25  (0.81 - 1.93) | 4.53E-01 |  | 1.54  (1.01 - 2.34) | 7.87E-02 |  | 0.81 | 6.92E-01 |  | Coronary atherosclerosis and other heart disease |
| CIR021 | 1.25  (0.44 - 3.54) | 7.45E-01 |  | 1.53  (0.97 - 2.40) | 9.98E-02 |  | 0.82 | 8.31E-01 |  | Acute hemorrhagic cerebrovascular disease |
| EXT020 | 6.14  (1.43 - 26.46) | 3.39E-02 | * | 3.63  (1.73 - 7.61) | 1.97E-03 | ** | 1.69 | 6.92E-01 |  | External cause codes: intent of injury, accidental/unintentional |
| EXT029 | 6.14  (1.43 - 26.46) | 3.39E-02 | * | 3.16  (1.54 - 6.50) | 3.55E-03 | ** | 1.94 | 6.92E-01 |  | External cause codes: subsequent encounter |
| GEN004 | 1.50  (0.39 - 5.73) | 6.83E-01 |  | 5.94  (2.47 - 14.24) | 3.54E-04 | *** | 0.25 | 4.05E-01 |  | Urinary tract infections |
| INF003 | 1.88  (0.60 - 5.85) | 4.45E-01 |  | 2.51  (0.88 - 7.18) | 1.14E-01 |  | 0.75 | 8.31E-01 |  | Bacterial infections |
| NEO015 | 1.13  (0.51 - 2.48) | 7.68E-01 |  | 1.64  (1.23 - 2.20) | 1.97E-03 | ** | 0.68 | 6.92E-01 |  | Gastrointestinal cancers - colorectal |
| NEO039 | 3.43  (1.43 - 8.24) | 3.06E-02 | * | 2.26  (1.63 - 3.14) | 8.07E-06 | *** | 1.52 | 6.92E-01 |  | Male reproductive system cancers - prostate |
| NVS004 | 6.66  (2.73 - 16.26) | 5.00E-04 | *** | 1.91  (1.35 - 2.69) | 1.01E-03 | ** | 3.50 | 1.76E-01 |  | Parkinson`s disease |
| NVS011 | 2.42  (1.15 - 5.11) | 4.03E-02 | * | 2.53  (1.97 - 3.25) | 4.59E-12 | *** | 0.96 | 9.11E-01 |  | Neurocognitive disorders |
| RSP002 | 1.88  (0.80 - 4.42) | 2.64E-01 |  | 1.69  (0.98 - 2.90) | 9.23E-02 |  | 1.11 | 8.88E-01 |  | Pneumonia (except that caused by tuberculosis) |
| RSP008 | 1.94  (1.24 - 3.06) | 3.06E-02 | * | 1.31  (0.88 - 1.93) | 2.21E-01 |  | 1.49 | 6.60E-01 |  | Chronic obstructive pulmonary disease and bronchiectasis |
| RSP016 | 0.79  (0.24 - 2.58) | 7.45E-01 |  | 1.31  (0.65 - 2.62) | 5.20E-01 |  | 0.61 | 6.92E-01 |  | Other specified and unspecified lower respiratory disease |
| HR(CI)_present_ : Hazard ratio and 95% confidence inteval for the risk factor present subgroup with the examined characteristics; p.adj_present_: FDR-adjusted p-value for the risk factor present subgroup; HR(CI)_absent_: Hazard ratio and 95% confidence inteval for the risk factor absent subgroup without the examined characteristics; p.adj_absent_: FDR-adjusted p-value for the risk factor absent subgroup; Sig_present_ and Sig_absent_: * indicates an FDR-adjusted p-value between 0.01 and 0.05, ** indicates between 0.001 and 0.01, and *** indicates a value smaller than 0.001 for the risk factor present and absent subgroups, respectively. RHR: Ratio of hazard ratios for comparisons between the risk factor present and absent subgroups. p.adj_RHR_*: FDR-adjusted p-value for comparisons between the risk factor present and absent subgroups, where a p.adj_RHR_ < 0.05 indicates significant differences.  CCSR: Clinical Classifications Software Refined, a method to classify disease categories. | | | | | | | | | | |

| **Supplementary Table 11a. Differences in Associations between Post-acute Mortality Risks in Organ Systems and Overall COVID-19 in Comparisons of Subgroup (HTN vs. HTN-free).** | | | | | | | | | | |
| --- | --- | --- | --- | --- | --- | --- | --- | --- | --- | --- |
| **Outcome** | **HR(CI)_present_** | **p.adj_present_** | **Sig_present_** | **HR(CI)_absent_** | **p.adj_absent_** | **Sig_absent_** | **RHR** | **p.adj_RHR_** | **Comparison** | **Organ System Description** |
| All-cause mortality | 1.59  (1.48 - 1.69) | 8.92E-42 | *** | 1.31  (1.19 - 1.44) | 2.87E-07 | *** | 1.21 | 7.52E-03 | HTN vs. HTN-free |  |
| DIG | 1.51  (1.00 - 2.28) | 5.65E-02 |  | 1.53  (0.66 - 3.53) | 3.75E-01 |  | 0.99 | 9.81E-01 |  | Diseases of the digestive system |
| EXT | 3.08  (1.29 - 7.32) | 1.92E-02 | * | 4.43  (1.71 - 11.50) | 3.83E-03 | ** | 0.69 | 9.52E-01 |  | External cause codes |
| INF | 1.34  (0.77 - 2.31) | 2.96E-01 |  | 0.65  (0.25 - 1.70) | 3.75E-01 |  | 2.07 | 4.62E-01 |  | Certain infectious and parasitic diseases |
| NEO | 1.41  (1.26 - 1.58) | 8.89E-09 | *** | 1.36  (1.17 - 1.57) | 1.20E-04 | *** | 1.04 | 9.52E-01 |  | Neoplasms |
| NVS | 2.41  (1.89 - 3.06) | 2.37E-12 | *** | 1.74  (1.31 - 2.33) | 3.70E-04 | *** | 1.38 | 3.23E-01 |  | Diseases of the nervous system |
| RSP | 1.34  (1.02 - 1.74) | 4.52E-02 | * | 1.31  (0.80 - 2.13) | 3.75E-01 |  | 1.02 | 9.81E-01 |  | Diseases of the respiratory system |
| **Supplementary Table 11b. Differences in Associations between Post-acute Mortality Risks in Specific Disorders and Overall COVID-19 in Comparisons of Subgroup (HTN vs. HTN-free).** | | | | | | | | | | |
| **Outcome** | **HR(CI)_present_** | **p.adj_present_** | **Sig_present_** | **HR(CI)_absent_** | **p.adj_absent_** | **Sig_absent_** | **RHR** | **p.adj_RHR_** | **Comparison** | **CCSR Category Description** |
| CIR009 | 0.87  (0.58 - 1.29) | 6.05E-01 |  | 0.50  (0.24 - 1.06) | 1.44E-01 |  | 1.72 | 7.49E-01 | HTN vs. HTN-free | Acute myocardial infarction |
| CIR011 | 1.43  (1.02 - 2.01) | 1.01E-01 |  | 0.59  (0.20 - 1.71) | 4.92E-01 |  | 2.44 | 7.49E-01 |  | Coronary atherosclerosis and other heart disease |
| CIR021 | 1.22  (0.68 - 2.18) | 6.05E-01 |  | 1.26  (0.60 - 2.67) | 6.51E-01 |  | 0.97 | 9.98E-01 |  | Acute hemorrhagic cerebrovascular disease |
| CIR026 | 3.63  (1.68 - 7.83) | 6.25E-03 | ** | 7.19  (1.87 - 27.68) | 1.48E-02 | * | 0.50 | 8.96E-01 |  | Peripheral and visceral vascular disease |
| EXT020 | 3.97  (1.50 - 10.49) | 2.44E-02 | * | 5.42  (2.02 - 14.51) | 4.67E-03 | ** | 0.73 | 9.84E-01 |  | External cause codes: intent of injury, accidental/unintentional |
| EXT029 | 3.58  (1.39 - 9.24) | 3.01E-02 | * | 4.43  (1.71 - 11.50) | 9.86E-03 | ** | 0.81 | 9.84E-01 |  | External cause codes: subsequent encounter |
| NEO012 | 1.07  (0.58 - 1.99) | 9.32E-01 |  | 0.47  (0.20 - 1.11) | 1.52E-01 |  | 2.27 | 7.49E-01 |  | Gastrointestinal cancers - esophagus |
| NEO015 | 1.02  (0.68 - 1.54) | 9.41E-01 |  | 1.68  (1.09 - 2.57) | 5.25E-02 |  | 0.61 | 7.49E-01 |  | Gastrointestinal cancers - colorectal |
| NEO018 | 2.13  (0.89 - 5.08) | 1.33E-01 |  | 2.46  (1.01 - 5.98) | 1.06E-01 |  | 0.87 | 9.84E-01 |  | Gastrointestinal cancers - bile duct |
| NEO025 | 2.21  (1.00 - 4.89) | 1.11E-01 |  | 1.02  (0.35 - 2.97) | 9.71E-01 |  | 2.17 | 7.62E-01 |  | Skin cancers - melanoma |
| NEO043 | 1.87  (0.96 - 3.65) | 1.20E-01 |  | 1.12  (0.42 - 2.97) | 8.62E-01 |  | 1.66 | 8.96E-01 |  | Urinary system cancers - bladder |
| NEO048 | 1.33  (0.68 - 2.60) | 5.64E-01 |  | 1.26  (0.63 - 2.51) | 6.51E-01 |  | 1.05 | 9.98E-01 |  | Nervous system cancers - brain |
| NEO051 | 0.98  (0.62 - 1.57) | 9.41E-01 |  | 0.82  (0.45 - 1.51) | 6.51E-01 |  | 1.19 | 9.84E-01 |  | Endocrine system cancers - pancreas |
| NEO071 | 1.83  (0.92 - 3.63) | 1.33E-01 |  | 2.12  (1.09 - 4.15) | 7.08E-02 |  | 0.86 | 9.84E-01 |  | Malignant neoplasm, unspecified |
| NVS004 | 2.53  (1.65 - 3.88) | 1.84E-04 | *** | 2.53  (1.48 - 4.32) | 4.67E-03 | ** | 1.00 | 9.98E-01 |  | Parkinson`s disease |
| NVS011 | 2.35  (1.69 - 3.27) | 6.47E-06 | *** | 2.18  (1.48 - 3.23) | 1.64E-03 | ** | 1.08 | 9.84E-01 |  | Neurocognitive disorders |
| RSP002 | 1.73  (0.97 - 3.08) | 1.20E-01 |  | 0.74  (0.25 - 2.20) | 6.67E-01 |  | 2.33 | 7.49E-01 |  | Pneumonia (except that caused by tuberculosis) |
| RSP008 | 1.46  (1.02 - 2.07) | 1.01E-01 |  | 1.60  (0.83 - 3.09) | 2.70E-01 |  | 0.91 | 9.84E-01 |  | Chronic obstructive pulmonary disease and bronchiectasis |
| **Supplementary Table 11c. Differences in Associations between Overall Mortality Risks in Organ Systems and Overall COVID-19 in Comparisons of Subgroup (HTN vs. HTN-free).** | | | | | | | | | | |
| **Outcome** | **HR(CI)_present_** | **p.adj_present_** | **Sig_present_** | **HR(CI)_absent_** | **p.adj_absent_** | **Sig_absent_** | **RHR** | **p.adj_RHR_** | **Comparison** | **CCSR Category Description** |
| All-cause mortality | 2.56  (2.42 - 2.70) | 1.44E-256 | *** | 2.04  (1.89 - 2.21) | 7.02E-70 | *** | 1.25 | 3.96E-05 | HTN vs. HTN-free |  |
| DIG | 1.91  (1.33 - 2.74) | 9.53E-04 | *** | 2.29  (1.21 - 4.33) | 1.76E-02 | * | 0.83 | 8.21E-01 |  | Diseases of the digestive system |
| EXT | 2.90  (1.33 - 6.34) | 9.96E-03 | ** | 3.96  (1.57 - 10.03) | 7.34E-03 | ** | 0.73 | 8.21E-01 |  | External cause codes |
| INF | 1.43  (0.87 - 2.34) | 1.81E-01 |  | 1.04  (0.47 - 2.28) | 9.29E-01 |  | 1.38 | 8.21E-01 |  | Certain infectious and parasitic diseases |
| INJ | 1.40  (0.70 - 2.79) | 3.36E-01 |  | 1.63  (0.53 - 5.07) | 4.53E-01 |  | 0.86 | 8.21E-01 |  | Injury poisoning and certain other consequences of external causes |
| NEO | 1.56  (1.40 - 1.73) | 3.88E-16 | *** | 1.52  (1.33 - 1.73) | 2.38E-09 | *** | 1.03 | 8.21E-01 |  | Neoplasms |
| NVS | 2.54  (2.02 - 3.18) | 2.99E-15 | *** | 1.78  (1.35 - 2.33) | 9.26E-05 | *** | 1.43 | 2.23E-01 |  | Diseases of the nervous system |
| RSP | 1.50  (1.17 - 1.92) | 2.38E-03 | ** | 1.60  (1.04 - 2.46) | 4.28E-02 | * | 0.94 | 8.21E-01 |  | Diseases of the respiratory system |
| **Supplementary Table 11d. Differences in Associations between Overall Mortality Risks in Specific Disorders and Overall COVID-19 in Comparisons of Subgroup (HTN vs. HTN-free).** | | | | | | | | | | |
| **Outcome** | **HR(CI)_present_** | **p.adj_present_** | **Sig_present_** | **HR(CI)_absent_** | **p.adj_absent_** | **Sig_absent_** | **RHR** | **p.adj_RHR_** | **Comparison** | **CCSR Category Description** |
| CIR009 | 1.09  (0.78 - 1.54) | 6.72E-01 |  | 0.55  (0.27 - 1.11) | 1.54E-01 |  | 2.00 | 3.99E-01 | HTN vs. HTN-free | Acute myocardial infarction |
| CIR011 | 1.57  (1.14 - 2.16) | 1.82E-02 | * | 0.81  (0.32 - 2.09) | 6.70E-01 |  | 1.93 | 6.48E-01 |  | Coronary atherosclerosis and other heart disease |
| CIR021 | 1.48  (0.88 - 2.48) | 1.89E-01 |  | 1.38  (0.69 - 2.75) | 4.43E-01 |  | 1.07 | 8.86E-01 |  | Acute hemorrhagic cerebrovascular disease |
| CIR026 | 3.23  (1.56 - 6.71) | 9.27E-03 | ** | 11.24  (3.30 - 38.28) | 1.25E-03 | ** | 0.29 | 3.99E-01 |  | Peripheral and visceral vascular disease |
| DIG025 | 1.93  (0.81 - 4.63) | 1.89E-01 |  | 10.09  (1.91 - 53.23) | 1.65E-02 | * | 0.19 | 3.99E-01 |  | Other specified and unspecified gastrointestinal disorders |
| EXT020 | 3.58  (1.46 - 8.78) | 1.82E-02 | * | 4.69  (1.81 - 12.19) | 1.16E-02 | * | 0.76 | 8.75E-01 |  | External cause codes: intent of injury, accidental/unintentional |
| EXT029 | 3.29  (1.36 - 7.93) | 2.05E-02 | * | 3.96  (1.57 - 10.03) | 1.33E-02 | * | 0.83 | 8.86E-01 |  | External cause codes: subsequent encounter |
| NEO012 | 1.03  (0.58 - 1.83) | 9.25E-01 |  | 0.67  (0.31 - 1.42) | 4.26E-01 |  | 1.54 | 8.75E-01 |  | Gastrointestinal cancers - esophagus |
| NEO015 | 1.22  (0.84 - 1.76) | 3.49E-01 |  | 1.85  (1.25 - 2.73) | 1.23E-02 | * | 0.66 | 4.98E-01 |  | Gastrointestinal cancers - colorectal |
| NEO018 | 2.09  (0.94 - 4.62) | 1.15E-01 |  | 2.69  (1.14 - 6.34) | 4.61E-02 | * | 0.77 | 8.75E-01 |  | Gastrointestinal cancers - bile duct |
| NEO025 | 2.13  (1.01 - 4.51) | 8.58E-02 |  | 1.41  (0.52 - 3.78) | 5.20E-01 |  | 1.51 | 8.75E-01 |  | Skin cancers - melanoma |
| NEO039 | 2.59  (1.78 - 3.78) | 8.60E-06 | *** | 2.22  (1.28 - 3.85) | 1.33E-02 | * | 1.17 | 8.75E-01 |  | Male reproductive system cancers - prostate |
| NEO043 | 2.16  (1.21 - 3.87) | 2.16E-02 | * | 1.40  (0.59 - 3.34) | 4.88E-01 |  | 1.54 | 8.75E-01 |  | Urinary system cancers - bladder |
| NEO048 | 1.45  (0.78 - 2.72) | 3.00E-01 |  | 1.36  (0.70 - 2.65) | 4.43E-01 |  | 1.07 | 8.86E-01 |  | Nervous system cancers - brain |
| NEO051 | 1.09  (0.71 - 1.67) | 7.29E-01 |  | 1.27  (0.77 - 2.10) | 4.43E-01 |  | 0.86 | 8.75E-01 |  | Endocrine system cancers - pancreas |
| NEO071 | 1.97  (1.06 - 3.66) | 6.18E-02 |  | 2.11  (1.10 - 4.05) | 4.61E-02 | * | 0.93 | 8.86E-01 |  | Malignant neoplasm, unspecified |
| NVS004 | 2.54  (1.69 - 3.81) | 5.96E-05 | *** | 2.06  (1.25 - 3.39) | 1.33E-02 | * | 1.23 | 8.75E-01 |  | Parkinson`s disease |
| NVS005 | 2.87  (0.79 - 10.50) | 1.69E-01 |  | 1.90  (0.59 - 6.13) | 4.26E-01 |  | 1.51 | 8.75E-01 |  | Multiple sclerosis |
| NVS011 | 2.76  (2.02 - 3.77) | 3.86E-09 | *** | 2.11  (1.46 - 3.06) | 1.25E-03 | ** | 1.31 | 7.98E-01 |  | Neurocognitive disorders |
| RSP002 | 2.19  (1.30 - 3.69) | 1.49E-02 | * | 0.62  (0.21 - 1.84) | 4.51E-01 |  | 3.51 | 3.99E-01 |  | Pneumonia (except that caused by tuberculosis) |
| RSP008 | 1.54  (1.10 - 2.15) | 2.49E-02 | * | 2.01  (1.13 - 3.58) | 4.00E-02 | * | 0.77 | 8.75E-01 |  | Chronic obstructive pulmonary disease and bronchiectasis |
| RSP016 | 0.62  (0.28 - 1.39) | 3.00E-01 |  | 2.57  (0.91 - 7.22) | 1.31E-01 |  | 0.24 | 3.99E-01 |  | Other specified and unspecified lower respiratory disease |
| HR(CI)_present_ : Hazard ratio and 95% confidence inteval for the risk factor present subgroup with the examined characteristics; p.adj_present_: FDR-adjusted p-value for the risk factor present subgroup; HR(CI)_absent_: Hazard ratio and 95% confidence inteval for the risk factor absent subgroup without the examined characteristics; p.adj_absent_: FDR-adjusted p-value for the risk factor absent subgroup; Sig_present_ and Sig_absent_: * indicates an FDR-adjusted p-value between 0.01 and 0.05, ** indicates between 0.001 and 0.01, and *** indicates a value smaller than 0.001 for the risk factor present and absent subgroups, respectively. RHR: Ratio of hazard ratios for comparisons between the risk factor present and absent subgroups. p.adj_RHR_*: FDR-adjusted p-value for comparisons between the risk factor present and absent subgroups, where a p.adj_RHR_ < 0.05 indicates significant differences.  CCSR: Clinical Classifications Software Refined, a method to classify disease categories. | | | | | | | | | | |

| **Supplementary Table 12a. Differences in Associations between Post-acute Mortality Risks in Organ Systems and Overall COVID-19 in Comparisons of Subgroup (Renal failure vs. Renal failure-free).** | | | | | | | | | | |
| --- | --- | --- | --- | --- | --- | --- | --- | --- | --- | --- |
| **Outcome** | **HR(CI)_present_** | **p.adj_present_** | **Sig_present_** | **HR(CI)_absent_** | **p.adj_absent_** | **Sig_absent_** | **RHR** | **p.adj_RHR_** | **Comparison** | **Organ System Description** |
| All-cause mortality | 1.46  (1.33 - 1.60) | 2.05E-14 | *** | 1.48  (1.38 - 1.58) | 3.37E-29 | *** | 0.99 | 8.24E-01 | Renal failure vs. Renal failure-free |  |
| CIR | 1.27  (1.00 - 1.61) | 8.32E-02 |  | 1.10  (0.89 - 1.36) | 4.26E-01 |  | 1.16 | 8.24E-01 |  | Diseases of the circulatory system |
| DIG | 1.81  (1.00 - 3.27) | 8.32E-02 |  | 1.39  (0.86 - 2.22) | 2.93E-01 |  | 1.30 | 8.24E-01 |  | Diseases of the digestive system |
| EXT | 4.94  (1.48 - 16.54) | 2.37E-02 | * | 3.41  (1.60 - 7.27) | 3.74E-03 | ** | 1.45 | 8.24E-01 |  | External cause codes |
| INF | 1.63  (0.74 - 3.61) | 3.22E-01 |  | 1.34  (0.77 - 2.33) | 4.26E-01 |  | 1.22 | 8.24E-01 |  | Certain infectious and parasitic diseases |
| INJ | 1.19  (0.46 - 3.09) | 7.18E-01 |  | 1.53  (0.62 - 3.77) | 4.26E-01 |  | 0.78 | 8.24E-01 |  | Injury poisoning and certain other consequences of external causes |
| MUS | 1.28  (0.41 - 3.95) | 7.18E-01 |  | 3.92  (1.26 - 12.18) | 3.67E-02 | * | 0.33 | 8.24E-01 |  | Diseases of the musculoskeletal system and connective tissue |
| NEO | 1.29  (1.09 - 1.53) | 8.36E-03 | ** | 1.38  (1.24 - 1.53) | 1.12E-08 | *** | 0.94 | 8.24E-01 |  | Neoplasms |
| NVS | 1.80  (1.29 - 2.52) | 2.74E-03 | ** | 2.33  (1.88 - 2.91) | 1.71E-13 | *** | 0.77 | 8.24E-01 |  | Diseases of the nervous system |
| RSP | 1.22  (0.86 - 1.74) | 3.38E-01 |  | 1.15  (0.82 - 1.60) | 4.26E-01 |  | 1.07 | 8.24E-01 |  | Diseases of the respiratory system |
| **Supplementary Table 12b. Differences in Associations between Post-acute Mortality Risks in Specific Disorders and Overall COVID-19 in Comparisons of Subgroup (Renal failure vs. Renal failure-free).** | | | | | | | | | | |
| **Outcome** | **HR(CI)_present_** | **p.adj_present_** | **Sig_present_** | **HR(CI)_absent_** | **p.adj_absent_** | **Sig_absent_** | **RHR** | **p.adj_RHR_** | **Comparison** | **CCSR Category Description** |
| CIR008 | 2.73  (0.90 - 8.27) | 1.38E-01 |  | 0.83  (0.36 - 1.95) | 7.57E-01 |  | 3.27 | 5.77E-01 | Renal failure vs. Renal failure-free | Hypertension with complications and secondary hypertension |
| CIR009 | 0.86  (0.52 - 1.41) | 5.75E-01 |  | 0.73  (0.44 - 1.22) | 3.80E-01 |  | 1.17 | 9.14E-01 |  | Acute myocardial infarction |
| CIR011 | 1.40  (0.91 - 2.14) | 2.01E-01 |  | 1.14  (0.69 - 1.88) | 7.55E-01 |  | 1.23 | 8.85E-01 |  | Coronary atherosclerosis and other heart disease |
| CIR019 | 2.62  (0.94 - 7.30) | 1.33E-01 |  | 1.43  (0.54 - 3.77) | 6.68E-01 |  | 1.83 | 8.85E-01 |  | Heart failure |
| CIR026 | 4.16  (1.24 - 13.90) | 6.60E-02 |  | 4.00  (1.78 - 8.95) | 3.15E-03 | ** | 1.04 | 9.57E-01 |  | Peripheral and visceral vascular disease |
| EXT020 | 6.19  (1.45 - 26.51) | 6.31E-02 |  | 4.29  (1.94 - 9.47) | 1.91E-03 | ** | 1.44 | 9.14E-01 |  | External cause codes: intent of injury, accidental/unintentional |
| EXT029 | 6.19  (1.45 - 26.51) | 6.31E-02 |  | 3.68  (1.71 - 7.93) | 3.15E-03 | ** | 1.68 | 8.85E-01 |  | External cause codes: subsequent encounter |
| NEO013 | 3.92  (1.18 - 13.02) | 6.60E-02 |  | 0.89  (0.36 - 2.18) | 8.45E-01 |  | 4.41 | 5.77E-01 |  | Gastrointestinal cancers - stomach |
| NEO015 | 0.97  (0.48 - 1.98) | 9.36E-01 |  | 1.48  (1.07 - 2.05) | 4.43E-02 | * | 0.65 | 8.35E-01 |  | Gastrointestinal cancers - colorectal |
| NEO018 | 5.53  (1.57 - 19.56) | 6.31E-02 |  | 1.79  (0.86 - 3.75) | 2.45E-01 |  | 3.09 | 5.88E-01 |  | Gastrointestinal cancers - bile duct |
| NEO048 | 0.50  (0.15 - 1.72) | 3.77E-01 |  | 1.61  (0.94 - 2.76) | 1.92E-01 |  | 0.31 | 5.77E-01 |  | Nervous system cancers - brain |
| NEO051 | 0.80  (0.39 - 1.62) | 5.75E-01 |  | 0.90  (0.58 - 1.39) | 7.55E-01 |  | 0.89 | 9.28E-01 |  | Endocrine system cancers - pancreas |
| NEO071 | 1.71  (0.57 - 5.08) | 4.33E-01 |  | 2.14  (1.28 - 3.59) | 1.10E-02 | * | 0.80 | 9.14E-01 |  | Malignant neoplasm, unspecified |
| NVS004 | 2.73  (1.49 - 5.02) | 2.19E-02 | * | 2.51  (1.64 - 3.85) | 2.10E-04 | *** | 1.09 | 9.28E-01 |  | Parkinson`s disease |
| NVS011 | 1.73  (1.08 - 2.78) | 6.60E-02 |  | 2.55  (1.90 - 3.43) | 9.76E-09 | *** | 0.68 | 6.18E-01 |  | Neurocognitive disorders |
| RSP002 | 2.12  (1.01 - 4.45) | 1.06E-01 |  | 1.28  (0.65 - 2.53) | 6.68E-01 |  | 1.66 | 8.35E-01 |  | Pneumonia (except that caused by tuberculosis) |
| RSP008 | 1.39  (0.86 - 2.23) | 2.69E-01 |  | 1.32  (0.85 - 2.05) | 3.80E-01 |  | 1.05 | 9.40E-01 |  | Chronic obstructive pulmonary disease and bronchiectasis |
| RSP016 | 0.60  (0.19 - 1.94) | 4.74E-01 |  | 0.94  (0.41 - 2.14) | 8.84E-01 |  | 0.64 | 8.85E-01 |  | Other specified and unspecified lower respiratory disease |
| **Supplementary Table 12c. Differences in Associations between Overall Mortality Risks in Organ Systems and Overall COVID-19 in Comparisons of Subgroup (Renal failure vs. Renal failure-free).** | | | | | | | | | | |
| **Outcome** | **HR(CI)_present_** | **p.adj_present_** | **Sig_present_** | **HR(CI)_absent_** | **p.adj_absent_** | **Sig_absent_** | **RHR** | **p.adj_RHR_** | **Comparison** | **CCSR Category Description** |
| All-cause mortality | 2.46  (2.28 - 2.65) | 1.25E-121 | *** | 2.29  (2.17 - 2.42) | 8.56E-191 | *** | 1.07 | 5.67E-01 | Renal failure vs. Renal failure-free |  |
| CIR | 1.56  (1.26 - 1.93) | 1.29E-04 | *** | 1.27  (1.05 - 1.54) | 2.22E-02 | * | 1.23 | 5.67E-01 |  | Diseases of the circulatory system |
| DIG | 2.23  (1.34 - 3.71) | 4.09E-03 | ** | 1.89  (1.27 - 2.81) | 3.42E-03 | ** | 1.18 | 6.80E-01 |  | Diseases of the digestive system |
| EXT | 3.99  (1.26 - 12.64) | 3.08E-02 | * | 3.28  (1.63 - 6.60) | 2.16E-03 | ** | 1.22 | 7.75E-01 |  | External cause codes |
| INF | 2.09  (1.07 - 4.07) | 3.91E-02 | * | 1.34  (0.79 - 2.27) | 2.73E-01 |  | 1.56 | 5.67E-01 |  | Certain infectious and parasitic diseases |
| INJ | 1.25  (0.51 - 3.05) | 6.27E-01 |  | 2.02  (0.92 - 4.44) | 9.44E-02 |  | 0.62 | 6.53E-01 |  | Injury poisoning and certain other consequences of external causes |
| MUS | 1.59  (0.55 - 4.58) | 4.36E-01 |  | 2.68  (1.02 - 7.08) | 6.64E-02 |  | 0.59 | 6.53E-01 |  | Diseases of the musculoskeletal system and connective tissue |
| NEO | 1.40  (1.20 - 1.63) | 7.80E-05 | *** | 1.55  (1.40 - 1.70) | 1.01E-17 | *** | 0.91 | 5.67E-01 |  | Neoplasms |
| NVS | 1.92  (1.40 - 2.63) | 1.29E-04 | *** | 2.39  (1.94 - 2.94) | 6.09E-16 | *** | 0.80 | 5.67E-01 |  | Diseases of the nervous system |
| RSP | 1.47  (1.05 - 2.04) | 3.40E-02 | * | 1.30  (0.96 - 1.77) | 9.44E-02 |  | 1.12 | 6.80E-01 |  | Diseases of the respiratory system |
| **Supplementary Table 12d. Differences in Associations between Overall Mortality Risks in Specific Disorders and Overall COVID-19 in Comparisons of Subgroup (Renal failure vs. Renal failure-free).** | | | | | | | | | | |
| **Outcome** | **HR(CI)_present_** | **p.adj_present_** | **Sig_present_** | **HR(CI)_absent_** | **p.adj_absent_** | **Sig_absent_** | **RHR** | **p.adj_RHR_** | **Comparison** | **CCSR Category Description** |
| CIR003 | 0.52  (0.13 - 2.10) | 5.10E-01 |  | 3.51  (1.38 - 8.90) | 2.24E-02 | * | 0.15 | 4.76E-01 | Renal failure vs. Renal failure-free | Nonrheumatic and unspecified valve disorders |
| CIR005 | 0.61  (0.20 - 1.91) | 5.46E-01 |  | 1.45  (0.51 - 4.15) | 5.84E-01 |  | 0.42 | 5.89E-01 |  | Myocarditis and cardiomyopathy |
| CIR008 | 3.02  (1.11 - 8.20) | 6.54E-02 |  | 0.96  (0.43 - 2.13) | 9.50E-01 |  | 3.14 | 4.76E-01 |  | Hypertension with complications and secondary hypertension |
| CIR009 | 1.06  (0.68 - 1.65) | 7.98E-01 |  | 0.84  (0.54 - 1.30) | 5.66E-01 |  | 1.27 | 8.41E-01 |  | Acute myocardial infarction |
| CIR011 | 1.74  (1.18 - 2.58) | 2.27E-02 | * | 1.14  (0.71 - 1.83) | 6.38E-01 |  | 1.53 | 5.63E-01 |  | Coronary atherosclerosis and other heart disease |
| CIR019 | 3.27  (1.25 - 8.58) | 4.78E-02 | * | 1.50  (0.61 - 3.67) | 5.42E-01 |  | 2.19 | 5.63E-01 |  | Heart failure |
| CIR021 | 1.21  (0.50 - 2.92) | 7.52E-01 |  | 1.42  (0.89 - 2.26) | 2.42E-01 |  | 0.85 | 9.56E-01 |  | Acute hemorrhagic cerebrovascular disease |
| CIR024 | 1.84  (0.58 - 5.89) | 4.55E-01 |  | 1.90  (0.31 - 11.49) | 5.84E-01 |  | 0.97 | 9.79E-01 |  | Other and ill-defined cerebrovascular disease |
| CIR026 | 4.70  (1.46 - 15.10) | 3.53E-02 | * | 4.29  (2.07 - 8.90) | 8.94E-04 | *** | 1.09 | 9.56E-01 |  | Peripheral and visceral vascular disease |
| DIG017 | 4.01  (1.03 - 15.71) | 8.11E-02 |  | 3.52  (1.06 - 11.73) | 9.38E-02 |  | 1.14 | 9.56E-01 |  | Biliary tract disease |
| EXT020 | 4.50  (1.18 - 17.19) | 6.42E-02 |  | 4.01  (1.89 - 8.50) | 1.79E-03 | ** | 1.12 | 9.56E-01 |  | External cause codes: intent of injury, accidental/unintentional |
| EXT029 | 4.50  (1.18 - 17.19) | 6.42E-02 |  | 3.49  (1.68 - 7.26) | 2.98E-03 | ** | 1.29 | 9.56E-01 |  | External cause codes: subsequent encounter |
| GEN004 | 6.95  (2.75 - 17.52) | 1.21E-03 | ** | 2.36  (0.77 - 7.26) | 2.37E-01 |  | 2.94 | 5.63E-01 |  | Urinary tract infections |
| INF003 | 3.33  (1.05 - 10.51) | 7.61E-02 |  | 1.54  (0.52 - 4.53) | 5.66E-01 |  | 2.16 | 6.76E-01 |  | Bacterial infections |
| NEO013 | 4.26  (1.57 - 11.56) | 2.23E-02 | * | 1.24  (0.58 - 2.64) | 6.38E-01 |  | 3.44 | 4.76E-01 |  | Gastrointestinal cancers - stomach |
| NEO015 | 1.10  (0.59 - 2.03) | 7.98E-01 |  | 1.71  (1.27 - 2.30) | 2.21E-03 | ** | 0.64 | 5.63E-01 |  | Gastrointestinal cancers - colorectal |
| NEO017 | 0.50  (0.15 - 1.69) | 4.18E-01 |  | 1.62  (0.63 - 4.19) | 4.97E-01 |  | 0.31 | 5.63E-01 |  | Gastrointestinal cancers - liver |
| NEO018 | 6.71  (1.98 - 22.78) | 1.68E-02 | * | 1.83  (0.92 - 3.62) | 1.60E-01 |  | 3.68 | 4.76E-01 |  | Gastrointestinal cancers - bile duct |
| NEO025 | 1.45  (0.49 - 4.23) | 6.26E-01 |  | 2.31  (1.14 - 4.68) | 5.13E-02 |  | 0.63 | 8.41E-01 |  | Skin cancers - melanoma |
| NEO039 | 2.33  (1.36 - 3.99) | 1.68E-02 | * | 2.41  (1.64 - 3.53) | 1.02E-04 | *** | 0.97 | 9.56E-01 |  | Male reproductive system cancers - prostate |
| NEO043 | 0.75  (0.27 - 2.05) | 6.87E-01 |  | 2.38  (1.34 - 4.22) | 9.45E-03 | ** | 0.31 | 4.76E-01 |  | Urinary system cancers - bladder |
| NEO048 | 0.68  (0.22 - 2.08) | 6.26E-01 |  | 1.61  (0.96 - 2.69) | 1.43E-01 |  | 0.42 | 5.63E-01 |  | Nervous system cancers - brain |
| NEO051 | 1.08  (0.58 - 2.01) | 7.98E-01 |  | 1.14  (0.78 - 1.67) | 5.88E-01 |  | 0.95 | 9.56E-01 |  | Endocrine system cancers - pancreas |
| NEO071 | 2.00  (0.78 - 5.13) | 2.50E-01 |  | 2.13  (1.30 - 3.49) | 9.00E-03 | ** | 0.94 | 9.56E-01 |  | Malignant neoplasm, unspecified |
| NVS004 | 2.52  (1.41 - 4.50) | 1.68E-02 | * | 2.06  (1.39 - 3.04) | 1.79E-03 | ** | 1.22 | 9.56E-01 |  | Parkinson`s disease |
| NVS011 | 1.95  (1.26 - 3.03) | 1.68E-02 | * | 2.71  (2.04 - 3.59) | 1.34E-10 | *** | 0.72 | 5.63E-01 |  | Neurocognitive disorders |
| RSP002 | 2.37  (1.19 - 4.70) | 4.63E-02 | * | 1.35  (0.72 - 2.55) | 5.26E-01 |  | 1.75 | 5.63E-01 |  | Pneumonia (except that caused by tuberculosis) |
| RSP008 | 1.63  (1.04 - 2.56) | 6.54E-02 |  | 1.50  (1.01 - 2.24) | 1.01E-01 |  | 1.09 | 9.56E-01 |  | Chronic obstructive pulmonary disease and bronchiectasis |
| RSP016 | 0.79  (0.27 - 2.27) | 7.52E-01 |  | 0.98  (0.45 - 2.16) | 9.70E-01 |  | 0.80 | 9.56E-01 |  | Other specified and unspecified lower respiratory disease |
| HR(CI)_present_ : Hazard ratio and 95% confidence inteval for the risk factor present subgroup with the examined characteristics; p.adj_present_: FDR-adjusted p-value for the risk factor present subgroup; HR(CI)_absent_: Hazard ratio and 95% confidence inteval for the risk factor absent subgroup without the examined characteristics; p.adj_absent_: FDR-adjusted p-value for the risk factor absent subgroup; Sig_present_ and Sig_absent_: * indicates an FDR-adjusted p-value between 0.01 and 0.05, ** indicates between 0.001 and 0.01, and *** indicates a value smaller than 0.001 for the risk factor present and absent subgroups, respectively. RHR: Ratio of hazard ratios for comparisons between the risk factor present and absent subgroups. p.adj_RHR_*: FDR-adjusted p-value for comparisons between the risk factor present and absent subgroups, where a p.adj_RHR_ < 0.05 indicates significant differences.  CCSR: Clinical Classifications Software Refined, a method to classify disease categories. | | | | | | | | | | |

| **Supplementary Table 13a. Differences in Associations between Post-acute Mortality Risks in Organ Systems and Overall COVID-19 in Comparisons of Subgroup (Stroke vs. Stroke-free).** | | | | | | | | | | |
| --- | --- | --- | --- | --- | --- | --- | --- | --- | --- | --- |
| **Outcome** | **HR(CI)_present_** | **p.adj_present_** | **Sig_present_** | **HR(CI)_absent_** | **p.adj_absent_** | **Sig_absent_** | **RHR** | **p.adj_RHR_** | **Comparison** | **Organ System Description** |
| All-cause mortality | 1.54 (1.37 - 1.74) | 1.22E-11 | *** | 1.54 (1.44 - 1.63) | 6.16E-43 | *** | 1.00 | 9.55E-01 | Stroke vs. Stroke-free |  |
| DIG | 1.64 (0.80 - 3.35) | 1.75E-01 |  | 1.47 (0.96 - 2.26) | 1.19E-01 |  | 1.12 | 9.54E-01 |  | Diseases of the digestive system |
| GEN | 4.80 (1.47 - 15.74) | 2.88E-02 | * | 2.13 (1.15 - 3.94) | 3.24E-02 | * | 2.25 | 6.71E-01 |  | Diseases of the genitourinary system |
| INF | 1.92 (0.78 - 4.71) | 1.75E-01 |  | 1.28 (0.76 - 2.17) | 3.56E-01 |  | 1.50 | 6.71E-01 |  | Certain infectious and parasitic diseases |
| NEO | 1.27 (0.99 - 1.64) | 9.40E-02 |  | 1.42 (1.29 - 1.56) | 2.51E-12 | *** | 0.90 | 6.71E-01 |  | Neoplasms |
| RSP | 1.80 (1.09 - 2.98) | 4.27E-02 | * | 1.15 (0.88 - 1.51) | 3.56E-01 |  | 1.57 | 6.71E-01 |  | Diseases of the respiratory system |
| **Supplementary Table 13b. Differences in Associations between Post-acute Mortality Risks in Specific Disorders and Overall COVID-19 in Comparisons of Subgroup (Stroke vs. Stroke-free).** | | | | | | | | | | |
| **Outcome** | **HR(CI)_present_** | **p.adj_present_** | **Sig_present_** | **HR(CI)_absent_** | **p.adj_absent_** | **Sig_absent_** | **RHR** | **p.adj_RHR_** | **Comparison** | **CCSR Category Description** |
| CIR009 | 0.73 (0.32 - 1.65) | 6.66E-01 |  | 0.78 (0.52 - 1.16) | 3.29E-01 |  | 0.93 | 8.76E-01 | Stroke vs. Stroke-free | Acute myocardial infarction |
| CIR011 | 2.62 (1.44 - 4.77) | 1.44E-02 | * | 0.98 (0.45 - 2.13) | 9.50E-01 |  | 2.69 | 1.63E-01 |  | Coronary atherosclerosis and other heart disease |
| CIR021 | 1.05 (0.42 - 2.63) | 9.12E-01 |  | 1.19 (0.70 - 2.02) | 5.84E-01 |  | 0.89 | 8.76E-01 |  | Acute hemorrhagic cerebrovascular disease |
| NEO015 | 2.78 (1.24 - 6.22) | 3.80E-02 | * | 1.28 (0.93 - 1.76) | 2.88E-01 |  | 2.18 | 1.63E-01 |  | Gastrointestinal cancers - colorectal |
| NEO048 | 1.19 (0.38 - 3.76) | 9.12E-01 |  | 1.43 (0.83 - 2.46) | 3.29E-01 |  | 0.83 | 8.76E-01 |  | Nervous system cancers - brain |
| NVS004 | 1.05 (0.52 - 2.13) | 9.12E-01 |  | 3.52 (2.39 - 5.19) | 1.29E-03 | ** | 0.30 | 1.41E-01 |  | Parkinson`s disease |
| NVS011 | 1.45 (0.85 - 2.47) | 3.03E-01 |  | 2.58 (1.94 - 3.42) | 5.75E-10 | *** | 0.56 | 1.63E-01 |  | Neurocognitive disorders |
| RSP002 | 3.95 (1.36 - 11.49) | 3.80E-02 | * | 1.38 (0.76 - 2.49) | 3.74E-01 |  | 2.87 | 1.63E-01 |  | Pneumonia (except that caused by tuberculosis) |
| RSP008 | 1.70 (0.89 - 3.23) | 2.42E-01 |  | 1.35 (0.93 - 1.95) | 2.88E-01 |  | 1.26 | 8.12E-01 |  | Chronic obstructive pulmonary disease and bronchiectasis |
| **Supplementary Table 13c. Differences in Associations between Overall Mortality Risks in Organ Systems and Overall COVID-19 in Comparisons of Subgroup (Stroke vs. Stroke-free).** | | | | | | | | | | |
| **Outcome** | **HR(CI)_present_** | **p.adj_present_** | **Sig_present_** | **HR(CI)_absent_** | **p.adj_absent_** | **Sig_absent_** | **RHR** | **p.adj_RHR_** | **Comparison** | **CCSR Category Description** |
| All-cause mortality | 2.48 (2.24 - 2.74) | 5.51E-69 | *** | 2.45 (2.33 - 2.57) | 3.21E-277 | *** | 1.01 | 8.77E-01 | Stroke vs. Stroke-free |  |
| DIG | 2.07 (1.07 - 3.99) | 4.29E-02 | * | 1.95 (1.36 - 2.79) | 5.85E-04 | *** | 1.06 | 8.77E-01 |  | Diseases of the digestive system |
| GEN | 6.02 (2.21 - 16.42) | 1.56E-03 | ** | 2.27 (1.28 - 4.02) | 8.99E-03 | ** | 2.66 | 2.91E-01 |  | Diseases of the genitourinary system |
| INF | 2.03 (0.86 - 4.77) | 1.21E-01 |  | 1.51 (0.95 - 2.39) | 9.59E-02 |  | 1.35 | 8.22E-01 |  | Certain infectious and parasitic diseases |
| MUS | 2.39 (0.56 - 10.19) | 2.39E-01 |  | 1.82 (0.79 - 4.19) | 1.60E-01 |  | 1.31 | 8.77E-01 |  | Diseases of the musculoskeletal system and connective tissue |
| NEO | 1.45 (1.15 - 1.83) | 3.95E-03 | ** | 1.58 (1.45 - 1.73) | 6.93E-24 | *** | 0.91 | 8.22E-01 |  | Neoplasms |
| RSP | 2.12 (1.31 - 3.44) | 3.95E-03 | ** | 1.33 (1.04 - 1.70) | 3.23E-02 | * | 1.59 | 2.91E-01 |  | Diseases of the respiratory system |
| **Supplementary Table 13d. Differences in Associations between Overall Mortality Risks in Specific Disorders and Overall COVID-19 in Comparisons of Subgroup (Stroke vs. Stroke-free).** | | | | | | | | | | |
| **Outcome** | **HR(CI)_present_** | **p.adj_present_** | **Sig_present_** | **HR(CI)_absent_** | **p.adj_absent_** | **Sig_absent_** | **RHR** | **p.adj_RHR_** | **Comparison** | **CCSR Category Description** |
| CIR003 | 3.70 (1.21 - 11.28) | 4.69E-02 | * | 1.19 (0.44 - 3.28) | 8.04E-01 |  | 3.10 | 2.81E-01 | Stroke vs. Stroke-free | Nonrheumatic and unspecified valve disorders |
| CIR009 | 0.82 (0.39 - 1.72) | 6.61E-01 |  | 1.00 (0.71 - 1.41) | 9.88E-01 |  | 0.82 | 8.24E-01 |  | Acute myocardial infarction |
| CIR011 | 2.62 (1.46 - 4.70) | 1.06E-02 | * | 1.20 (0.84 - 1.70) | 3.84E-01 |  | 2.19 | 1.25E-01 |  | Coronary atherosclerosis and other heart disease |
| CIR021 | 1.41 (0.65 - 3.06) | 5.37E-01 |  | 1.36 (0.83 - 2.21) | 3.01E-01 |  | 1.04 | 9.41E-01 |  | Acute hemorrhagic cerebrovascular disease |
| NEO015 | 2.76 (1.32 - 5.77) | 2.54E-02 | * | 1.47 (1.10 - 1.96) | 2.36E-02 | * | 1.87 | 2.81E-01 |  | Gastrointestinal cancers - colorectal |
| NEO039 | 2.71 (1.17 - 6.27) | 4.69E-02 | * | 2.37 (1.70 - 3.32) | 1.66E-06 | *** | 1.14 | 8.45E-01 |  | Male reproductive system cancers - prostate |
| NEO048 | 1.38 (0.49 - 3.93) | 6.61E-01 |  | 1.76 (1.05 - 2.95) | 6.00E-02 |  | 0.79 | 8.24E-01 |  | Nervous system cancers - brain |
| NVS004 | 1.16 (0.59 - 2.26) | 6.67E-01 |  | 3.05 (2.12 - 4.37) | 7.84E-09 | *** | 0.38 | 1.25E-01 |  | Parkinson`s disease |
| NVS011 | 1.48 (0.88 - 2.46) | 2.15E-01 |  | 2.79 (2.13 - 3.64) | 6.59E-13 | *** | 0.53 | 1.25E-01 |  | Neurocognitive disorders |
| RSP002 | 5.22 (1.84 - 14.85) | 1.06E-02 | * | 1.65 (0.98 - 2.80) | 9.74E-02 |  | 3.16 | 1.61E-01 |  | Pneumonia (except that caused by tuberculosis) |
| RSP008 | 1.83 (0.99 - 3.41) | 1.01E-01 |  | 1.51 (1.08 - 2.12) | 3.56E-02 | * | 1.21 | 8.24E-01 |  | Chronic obstructive pulmonary disease and bronchiectasis |
| HR(CI)_present_ : Hazard ratio and 95% confidence inteval for the risk factor present subgroup with the examined characteristics; p.adj_present_: FDR-adjusted p-value for the risk factor present subgroup; HR(CI)_absent_: Hazard ratio and 95% confidence inteval for the risk factor absent subgroup without the examined characteristics; p.adj_absent_: FDR-adjusted p-value for the risk factor absent subgroup; Sig_present_ and Sig_absent_: * indicates an FDR-adjusted p-value between 0.01 and 0.05, ** indicates between 0.001 and 0.01, and *** indicates a value smaller than 0.001 for the risk factor present and absent subgroups, respectively. RHR: Ratio of hazard ratios for comparisons between the risk factor present and absent subgroups. p.adj_RHR_*: FDR-adjusted p-value for comparisons between the risk factor present and absent subgroups, where a p.adj_RHR_ < 0.05 indicates significant differences.  CCSR: Clinical Classifications Software Refined, a method to classify disease categories. | | | | | | | | | | |

| **Supplementary Table 14a. Differences in Associations between Post-acute Mortality Risks in Organ Systems and Overall COVID-19 in Comparisons of Subgroup (T2DM vs. T2DM-free).** | | | | | | | | | | |
| --- | --- | --- | --- | --- | --- | --- | --- | --- | --- | --- |
| **Outcome** | **HR(CI)_present_** | **p.adj_present_** | **Sig_present_** | **HR(CI)_absent_** | **p.adj_absent_** | **Sig_absent_** | **RHR** | **p.adj_RHR_** | **Comparison** | **Organ System Description** |
| All-cause mortality | 1.52 (1.36 - 1.70) | 2.71E-12 | *** | 1.52 (1.43 - 1.61) | 4.21E-39 | *** | 1.00 | 9.61E-01 | T2DM vs. T2DM-free |  |
| CIR | 1.56 (1.19 - 2.06) | 6.84E-03 | ** | 1.05 (0.87 - 1.28) | 6.13E-01 |  | 1.49 | 1.93E-01 |  | Diseases of the circulatory system |
| DIG | 1.88 (0.97 - 3.62) | 9.02E-02 |  | 1.23 (0.79 - 1.91) | 4.03E-01 |  | 1.53 | 6.24E-01 |  | Diseases of the digestive system |
| EXT | 5.94 (1.38 - 25.51) | 4.97E-02 | * | 3.23 (1.57 - 6.66) | 3.27E-03 | ** | 1.84 | 6.68E-01 |  | External cause codes |
| GEN | 1.86 (0.83 - 4.17) | 1.70E-01 |  | 2.64 (1.30 - 5.36) | 1.26E-02 | * | 0.70 | 6.68E-01 |  | Diseases of the genitourinary system |
| INF | 0.85 (0.36 - 2.03) | 7.20E-01 |  | 1.40 (0.80 - 2.44) | 3.04E-01 |  | 0.61 | 6.24E-01 |  | Certain infectious and parasitic diseases |
| NEO | 1.25 (1.02 - 1.55) | 6.21E-02 |  | 1.48 (1.34 - 1.64) | 1.36E-14 | *** | 0.84 | 6.24E-01 |  | Neoplasms |
| NVS | 1.72 (1.05 - 2.82) | 6.21E-02 |  | 2.25 (1.84 - 2.74) | 5.43E-15 | *** | 0.77 | 6.24E-01 |  | Diseases of the nervous system |
| RSP | 1.29 (0.83 - 2.00) | 2.96E-01 |  | 1.30 (0.99 - 1.72) | 9.07E-02 |  | 0.99 | 9.61E-01 |  | Diseases of the respiratory system |
| **Supplementary Table 14b. Differences in Associations between Post-acute Mortality Risks in Specific Disorders and Overall COVID-19 in Comparisons of Subgroup (T2DM vs. T2DM-free).** | | | | | | | | | | |
| **Outcome** | **HR(CI)_present_** | **p.adj_present_** | **Sig_present_** | **HR(CI)_absent_** | **p.adj_absent_** | **Sig_absent_** | **RHR** | **p.adj_RHR_** | **Comparison** | **CCSR Category Description** |
| CIR008 | 1.39 (0.49 - 3.98) | 7.53E-01 |  | 1.40 (0.62 - 3.19) | 5.86E-01 |  | 0.99 | 9.90E-01 | T2DM vs. T2DM-free | Hypertension with complications and secondary hypertension |
| CIR009 | 1.00 (0.54 - 1.85) | 9.90E-01 |  | 0.58 (0.37 - 0.90) | 7.40E-02 |  | 1.72 | 7.65E-01 |  | Acute myocardial infarction |
| CIR011 | 1.98 (1.21 - 3.22) | 8.66E-02 |  | 0.98 (0.62 - 1.53) | 9.37E-01 |  | 2.02 | 5.28E-01 |  | Coronary atherosclerosis and other heart disease |
| CIR019 | 1.55 (0.57 - 4.20) | 7.43E-01 |  | 1.82 (0.68 - 4.87) | 4.59E-01 |  | 0.85 | 9.90E-01 |  | Heart failure |
| CIR021 | 1.44 (0.53 - 3.90) | 7.43E-01 |  | 1.03 (0.61 - 1.77) | 9.37E-01 |  | 1.39 | 9.90E-01 |  | Acute hemorrhagic cerebrovascular disease |
| GEN004 | 3.44 (1.28 - 9.24) | 1.01E-01 |  | 2.00 (0.64 - 6.31) | 4.59E-01 |  | 1.71 | 9.90E-01 |  | Urinary tract infections |
| NEO013 | 3.21 (1.06 - 9.73) | 1.36E-01 |  | 0.96 (0.39 - 2.38) | 9.37E-01 |  | 3.33 | 6.90E-01 |  | Gastrointestinal cancers - stomach |
| NEO015 | 1.14 (0.55 - 2.38) | 8.38E-01 |  | 1.41 (1.02 - 1.94) | 1.29E-01 |  | 0.81 | 9.90E-01 |  | Gastrointestinal cancers - colorectal |
| NEO043 | 1.36 (0.35 - 5.32) | 8.38E-01 |  | 1.32 (0.72 - 2.40) | 5.73E-01 |  | 1.03 | 9.90E-01 |  | Urinary system cancers - bladder |
| NEO051 | 0.93 (0.43 - 2.04) | 9.31E-01 |  | 0.90 (0.59 - 1.37) | 7.99E-01 |  | 1.04 | 9.90E-01 |  | Endocrine system cancers - pancreas |
| NVS004 | 1.98 (0.71 - 5.53) | 4.48E-01 |  | 2.56 (1.78 - 3.69) | 2.81E-06 | *** | 0.77 | 9.90E-01 |  | Parkinson`s disease |
| NVS011 | 2.09 (1.10 - 3.96) | 1.11E-01 |  | 2.29 (1.74 - 3.01) | 4.29E-08 | *** | 0.91 | 9.90E-01 |  | Neurocognitive disorders |
| RSP002 | 1.47 (0.56 - 3.84) | 7.43E-01 |  | 1.41 (0.77 - 2.59) | 4.59E-01 |  | 1.04 | 9.90E-01 |  | Pneumonia (except that caused by tuberculosis) |
| RSP008 | 1.73 (0.96 - 3.12) | 1.87E-01 |  | 1.24 (0.85 - 1.81) | 4.59E-01 |  | 1.40 | 9.90E-01 |  | Chronic obstructive pulmonary disease and bronchiectasis |
| CIR008 | 1.39 (0.49 - 3.98) | 7.53E-01 |  | 1.40 (0.62 - 3.19) | 5.86E-01 |  | 0.99 | 9.90E-01 |  | Hypertension with complications and secondary hypertension |
| CIR009 | 1.00 (0.54 - 1.85) | 9.90E-01 |  | 0.58 (0.37 - 0.90) | 7.40E-02 |  | 1.72 | 7.65E-01 |  | Acute myocardial infarction |
| CIR011 | 1.98 (1.21 - 3.22) | 8.66E-02 |  | 0.98 (0.62 - 1.53) | 9.37E-01 |  | 2.02 | 5.28E-01 |  | Coronary atherosclerosis and other heart disease |
| CIR019 | 1.55 (0.57 - 4.20) | 7.43E-01 |  | 1.82 (0.68 - 4.87) | 4.59E-01 |  | 0.85 | 9.90E-01 |  | Heart failure |
| CIR021 | 1.44 (0.53 - 3.90) | 7.43E-01 |  | 1.03 (0.61 - 1.77) | 9.37E-01 |  | 1.39 | 9.90E-01 |  | Acute hemorrhagic cerebrovascular disease |
| GEN004 | 3.44 (1.28 - 9.24) | 1.01E-01 |  | 2.00 (0.64 - 6.31) | 4.59E-01 |  | 1.71 | 9.90E-01 |  | Urinary tract infections |
| **Supplementary Table 14c. Differences in Associations between Overall Mortality Risks in Organ Systems and Overall COVID-19 in Comparisons of Subgroup (T2DM vs. T2DM-free).** | | | | | | | | | | |
| **Outcome** | **HR(CI)_present_** | **p.adj_present_** | **Sig_present_** | **HR(CI)_absent_** | **p.adj_absent_** | **Sig_absent_** | **RHR** | **p.adj_RHR_** | **Comparison** | **CCSR Category Description** |
| All-cause mortality | 2.68 (2.45 - 2.94) | 1.59E-100 | *** | 2.34 (2.23 - 2.46) | 1.99E-236 | *** | 1.15 | 6.55E-02 | T2DM vs. T2DM-free |  |
| CIR | 1.84 (1.43 - 2.36) | 8.72E-06 | *** | 1.26 (1.06 - 1.50) | 1.06E-02 | * | 1.46 | 6.55E-02 |  | Diseases of the circulatory system |
| DIG | 2.20 (1.26 - 3.86) | 8.87E-03 | ** | 1.72 (1.19 - 2.50) | 7.38E-03 | ** | 1.28 | 7.58E-01 |  | Diseases of the digestive system |
| EXT | 2.84 (0.85 - 9.49) | 1.02E-01 |  | 3.53 (1.78 - 7.00) | 7.02E-04 | *** | 0.80 | 7.58E-01 |  | External cause codes |
| GEN | 2.99 (1.48 - 6.02) | 4.92E-03 | ** | 2.43 (1.25 - 4.71) | 1.06E-02 | * | 1.23 | 7.58E-01 |  | Diseases of the genitourinary system |
| INF | 1.03 (0.49 - 2.16) | 9.41E-01 |  | 1.57 (0.95 - 2.60) | 7.75E-02 |  | 0.65 | 7.58E-01 |  | Certain infectious and parasitic diseases |
| NEO | 1.39 (1.15 - 1.68) | 2.33E-03 | ** | 1.64 (1.49 - 1.79) | 1.21E-25 | *** | 0.85 | 3.95E-01 |  | Neoplasms |
| NVS | 2.04 (1.28 - 3.27) | 5.17E-03 | ** | 2.31 (1.91 - 2.78) | 5.66E-18 | *** | 0.88 | 7.58E-01 |  | Diseases of the nervous system |
| RSP | 1.61 (1.08 - 2.40) | 2.47E-02 | * | 1.45 (1.12 - 1.87) | 7.38E-03 | ** | 1.12 | 7.58E-01 |  | Diseases of the respiratory system |
| **Supplementary Table 14d. Differences in Associations between Overall Mortality Risks in Specific Disorders and Overall COVID-19 in Comparisons of Subgroup (T2DM vs. T2DM-free).** | | | | | | | | | | |
| **Outcome** | **HR(CI)_present_** | **p.adj_present_** | **Sig_present_** | **HR(CI)_absent_** | **p.adj_absent_** | **Sig_absent_** | **RHR** | **p.adj_RHR_** | **Comparison** | **CCSR Category Description** |
| CIR008 | 1.69 (0.63 - 4.52) | 3.83E-01 |  | 1.52 (0.72 - 3.19) | 3.40E-01 |  | 1.11 | 9.21E-01 | T2DM vs. T2DM-free | Hypertension with complications and secondary hypertension |
| CIR009 | 1.35 (0.81 - 2.26) | 3.56E-01 |  | 0.71 (0.47 - 1.05) | 1.76E-01 |  | 1.92 | 9.21E-01 |  | Acute myocardial infarction |
| CIR011 | 2.02 (1.27 - 3.22) | 1.93E-02 | * | 1.20 (0.80 - 1.80) | 4.36E-01 |  | 1.69 | 9.21E-01 |  | Coronary atherosclerosis and other heart disease |
| CIR019 | 1.83 (0.70 - 4.77) | 3.38E-01 |  | 2.14 (0.89 - 5.14) | 1.76E-01 |  | 0.85 | 9.21E-01 |  | Heart failure |
| CIR021 | 1.92 (0.79 - 4.68) | 3.00E-01 |  | 1.23 (0.76 - 2.00) | 4.36E-01 |  | 1.56 | 9.21E-01 |  | Acute hemorrhagic cerebrovascular disease |
| CIR026 | 2.70 (0.64 - 11.43) | 3.21E-01 |  | 4.24 (2.13 - 8.41) | 1.85E-04 | *** | 0.64 | 9.21E-01 |  | Peripheral and visceral vascular disease |
| DIG017 | 4.36 (1.02 - 18.73) | 1.27E-01 |  | 2.83 (0.91 - 8.78) | 1.76E-01 |  | 1.54 | 9.21E-01 |  | Biliary tract disease |
| GEN004 | 5.01 (2.00 - 12.58) | 1.18E-02 | * | 2.36 (0.80 - 6.98) | 2.02E-01 |  | 2.13 | 9.21E-01 |  | Urinary tract infections |
| INF003 | 2.09 (0.65 - 6.75) | 3.38E-01 |  | 1.84 (0.67 - 5.10) | 3.20E-01 |  | 1.13 | 9.21E-01 |  | Bacterial infections |
| NEO013 | 3.31 (1.18 - 9.27) | 7.58E-02 |  | 1.57 (0.77 - 3.18) | 3.04E-01 |  | 2.11 | 9.21E-01 |  | Gastrointestinal cancers - stomach |
| NEO015 | 1.38 (0.73 - 2.60) | 3.83E-01 |  | 1.60 (1.19 - 2.15) | 7.32E-03 | ** | 0.86 | 9.21E-01 |  | Gastrointestinal cancers - colorectal |
| NEO017 | 1.11 (0.38 - 3.19) | 8.50E-01 |  | 1.34 (0.53 - 3.40) | 5.67E-01 |  | 0.83 | 9.21E-01 |  | Gastrointestinal cancers - liver |
| NEO039 | 2.43 (1.20 - 4.92) | 5.36E-02 |  | 2.48 (1.75 - 3.53) | 4.04E-06 | *** | 0.98 | 9.58E-01 |  | Male reproductive system cancers - prostate |
| NEO043 | 3.19 (0.96 - 10.52) | 1.27E-01 |  | 1.65 (0.97 - 2.81) | 1.76E-01 |  | 1.93 | 9.21E-01 |  | Urinary system cancers - bladder |
| NEO048 | 1.79 (0.50 - 6.47) | 3.94E-01 |  | 1.40 (0.85 - 2.32) | 2.82E-01 |  | 1.27 | 9.21E-01 |  | Nervous system cancers - brain |
| NEO051 | 1.35 (0.71 - 2.57) | 3.94E-01 |  | 1.09 (0.75 - 1.59) | 6.57E-01 |  | 1.24 | 9.21E-01 |  | Endocrine system cancers - pancreas |
| NVS004 | 1.67 (0.62 - 4.53) | 3.83E-01 |  | 2.35 (1.68 - 3.31) | 5.13E-06 | *** | 0.71 | 9.21E-01 |  | Parkinson`s disease |
| NVS011 | 2.56 (1.40 - 4.67) | 1.93E-02 | * | 2.42 (1.87 - 3.14) | 5.00E-10 | *** | 1.06 | 9.21E-01 |  | Neurocognitive disorders |
| RSP002 | 2.32 (1.00 - 5.38) | 1.27E-01 |  | 1.57 (0.91 - 2.72) | 1.94E-01 |  | 1.48 | 9.21E-01 |  | Pneumonia (except that caused by tuberculosis) |
| RSP008 | 1.99 (1.16 - 3.39) | 5.36E-02 |  | 1.39 (0.98 - 1.98) | 1.76E-01 |  | 1.43 | 9.21E-01 |  | Chronic obstructive pulmonary disease and bronchiectasis |
| HR(CI)_present_ : Hazard ratio and 95% confidence inteval for the risk factor present subgroup with the examined characteristics; p.adj_present_: FDR-adjusted p-value for the risk factor present subgroup; HR(CI)_absent_: Hazard ratio and 95% confidence inteval for the risk factor absent subgroup without the examined characteristics; p.adj_absent_: FDR-adjusted p-value for the risk factor absent subgroup; Sig_present_ and Sig_absent_: * indicates an FDR-adjusted p-value between 0.01 and 0.05, ** indicates between 0.001 and 0.01, and *** indicates a value smaller than 0.001 for the risk factor present and absent subgroups, respectively. RHR: Ratio of hazard ratios for comparisons between the risk factor present and absent subgroups. p.adj_RHR_*: FDR-adjusted p-value for comparisons between the risk factor present and absent subgroups, where a p.adj_RHR_ < 0.05 indicates significant differences.  CCSR: Clinical Classifications Software Refined, a method to classify disease categories. | | | | | | | | | | |

| **Supplementary Table 15a. Differences in Associations between Post-acute Mortality Risks in Organ Systems and Overall COVID-19 in Comparisons of Subgroup (Advanced age vs. Middle age).** | | | | | | | | | | |
| --- | --- | --- | --- | --- | --- | --- | --- | --- | --- | --- |
| **Outcome** | **HR(CI)_present_** | **p.adj_present_** | **Sig_present_** | **HR(CI)_absent_** | **p.adj_absent_** | **Sig_absent_** | **RHR** | **p.adj_RHR_** | **Comparison** | **Organ System Description** |
| All-cause mortality | 1.61  (1.52 - 1.70) | 3.84E-59 | *** | 1.15  (0.98 - 1.36) | 1.94E-01 |  | 1.40 | 1.18E-03 | Advanced age^1^ vs. Middle age^2^ |  |
| CIR | 1.37  (1.16 - 1.61) | 2.39E-04 | *** | 0.51  (0.28 - 0.92) | 1.50E-01 |  | 2.67 | 4.61E-03 |  | Diseases of the circulatory system |
| INF | 1.41  (0.82 - 2.44) | 2.15E-01 |  | 0.95  (0.39 - 2.33) | 9.17E-01 |  | 1.48 | 4.62E-01 |  | Certain infectious and parasitic diseases |
| NEO | 1.46  (1.33 - 1.61) | 1.84E-14 | *** | 1.23  (0.96 - 1.56) | 1.94E-01 |  | 1.20 | 3.28E-01 |  | Neoplasms |
| NVS | 2.23  (1.85 - 2.69) | 1.98E-16 | *** | 1.31  (0.52 - 3.33) | 6.79E-01 |  | 1.70 | 3.28E-01 |  | Diseases of the nervous system |
| RSP | 1.30  (1.02 - 1.66) | 3.96E-02 | * | 0.73  (0.30 - 1.82) | 6.79E-01 |  | 1.77 | 3.28E-01 |  | Diseases of the respiratory system |
| **Supplementary Table 15b. Differences in Associations between Post-acute Mortality Risks in Specific Disorders and Overall COVID-19 in Comparisons of Subgroup (Advanced age vs. Middle age).** | | | | | | | | | | |
| **Outcome** | **HR(CI)_present_** | **p.adj_present_** | **Sig_present_** | **HR(CI)_absent_** | **p.adj_absent_** | **Sig_absent_** | **RHR** | **p.adj_RHR_** | **Comparison** | **CCSR Category Description** |
| CIR011 | 1.50  (1.07 - 2.11) | 4.78E-02 | * | 0.31  (0.09 - 1.11) | 1.80E-01 |  | 4.85 | 9.62E-02 | Advanced age^1^ vs. Middle age^2^ | Coronary atherosclerosis and other heart disease |
| NEO012 | 0.78  (0.44 - 1.39) | 5.00E-01 |  | 1.18  (0.42 - 3.31) | 7.53E-01 |  | 0.66 | 6.90E-01 |  | Gastrointestinal cancers - esophagus |
| NEO015 | 1.39  (1.00 - 1.92) | 7.97E-02 |  | 1.68  (0.86 - 3.31) | 2.18E-01 |  | 0.83 | 6.90E-01 |  | Gastrointestinal cancers - colorectal |
| NEO051 | 0.97  (0.65 - 1.45) | 8.91E-01 |  | 0.78  (0.28 - 2.18) | 7.53E-01 |  | 1.25 | 6.90E-01 |  | Endocrine system cancers - pancreas |
| NVS011 | 2.35  (1.82 - 3.03) | 2.00E-10 | *** | 4.32  (1.05 - 17.80) | 1.80E-01 |  | 0.54 | 6.90E-01 |  | Neurocognitive disorders |
| **Supplementary Table 15c. Differences in Associations between Overall Mortality Risks in Organ Systems and Overall COVID-19 in Comparisons of Subgroup (Advanced age vs. Middle age).** | | | | | | | | | | |
| **Outcome** | **HR(CI)_present_** | **p.adj_present_** | **Sig_present_** | **HR(CI)_absent_** | **p.adj_absent_** | **Sig_absent_** | **RHR** | **p.adj_RHR_** | **Comparison** | **CCSR Category Description** |
| All-cause mortality | 2.58  (2.46 - 2.70) | 0.00E+00 | *** | 1.70  (1.48 - 1.95) | 5.87E-13 | *** | 1.52 | 1.98E-07 | Advanced age^1^ vs. Middle age^2^ |  |
| CIR | 1.59  (1.38 - 1.84) | 7.44E-10 | *** | 0.77  (0.48 - 1.24) | 6.65E-01 |  | 2.07 | 1.60E-02 |  | Diseases of the circulatory system |
| DIG | 2.14  (1.55 - 2.96) | 5.93E-06 | *** | 0.70  (0.23 - 2.13) | 7.82E-01 |  | 3.04 | 1.02E-01 |  | Diseases of the digestive system |
| INF | 1.64  (1.01 - 2.66) | 4.53E-02 | * | 1.12  (0.50 - 2.50) | 7.82E-01 |  | 1.46 | 4.26E-01 |  | Certain infectious and parasitic diseases |
| NEO | 1.64  (1.50 - 1.79) | 1.70E-27 | *** | 1.30  (1.04 - 1.62) | 7.30E-02 |  | 1.26 | 1.02E-01 |  | Neoplasms |
| NVS | 2.34  (1.96 - 2.80) | 9.48E-21 | *** | 1.22  (0.49 - 3.05) | 7.82E-01 |  | 1.92 | 2.40E-01 |  | Diseases of the nervous system |
| RSP | 1.49  (1.19 - 1.87) | 5.46E-04 | *** | 0.89  (0.40 - 1.99) | 7.82E-01 |  | 1.68 | 2.63E-01 |  | Diseases of the respiratory system |
| **Supplementary Table 15d. Differences in Associations between Overall Mortality Risks in Specific Disorders and Overall COVID-19 in Comparisons of Subgroup (Advanced age vs. Middle age).** | | | | | | | | | | |
| **Outcome** | **HR(CI)_present_** | **p.adj_present_** | **Sig_present_** | **HR(CI)_absent_** | **p.adj_absent_** | **Sig_absent_** | **RHR** | **p.adj_RHR_** | **Comparison** | **CCSR Category Description** |
| CIR009 | 0.99  (0.71 - 1.38) | 9.44E-01 |  | 0.86  (0.36 - 2.05) | 8.68E-01 |  | 1.15 | 9.78E-01 | Advanced age^1^ vs. Middle age^2^ | Acute myocardial infarction |
| CIR011 | 1.65  (1.21 - 2.26) | 3.46E-03 | ** | 0.43  (0.13 - 1.35) | 3.87E-01 |  | 3.88 | 1.57E-01 |  | Coronary atherosclerosis and other heart disease |
| NEO012 | 0.93  (0.56 - 1.56) | 9.44E-01 |  | 0.92  (0.34 - 2.50) | 8.68E-01 |  | 1.02 | 9.78E-01 |  | Gastrointestinal cancers - esophagus |
| NEO015 | 1.65  (1.22 - 2.23) | 3.46E-03 | ** | 1.51  (0.81 - 2.82) | 3.87E-01 |  | 1.09 | 9.78E-01 |  | Gastrointestinal cancers - colorectal |
| NEO051 | 1.20  (0.84 - 1.70) | 4.77E-01 |  | 1.22  (0.52 - 2.87) | 8.68E-01 |  | 0.98 | 9.78E-01 |  | Endocrine system cancers - pancreas |
| NVS011 | 2.56  (2.01 - 3.25) | 8.75E-14 | *** | 3.20  (0.87 - 11.77) | 3.87E-01 |  | 0.80 | 9.78E-01 |  | Neurocognitive disorders |
| 1. Advanced age: >65 years old. 2. Middle age: 50-65 years old. | | | | | | | | | | |
| HR(CI)_present_ : Hazard ratio and 95% confidence inteval for the risk factor present subgroup with the examined characteristics; p.adj_present_: FDR-adjusted p-value for the risk factor present subgroup; HR(CI)_absent_: Hazard ratio and 95% confidence inteval for the risk factor absent subgroup without the examined characteristics; p.adj_absent_: FDR-adjusted p-value for the risk factor absent subgroup; Sig_present_ and Sig_absent_: * indicates an FDR-adjusted p-value between 0.01 and 0.05, ** indicates between 0.001 and 0.01, and *** indicates a value smaller than 0.001 for the risk factor present and absent subgroups, respectively. RHR: Ratio of hazard ratios for comparisons between the risk factor present and absent subgroups. p.adj_RHR_*: FDR-adjusted p-value for comparisons between the risk factor present and absent subgroups, where a p.adj_RHR_ < 0.05 indicates significant differences.  CCSR: Clinical Classifications Software Refined, a method to classify disease categories. | | | | | | | | | | |

| **Supplementary Table 16a. Differences in Associations between Post-acute Mortality Risks in Organ Systems and Overall COVID-19 in Comparisons of Subgroup (Male vs. Female).** | | | | | | | | | | |
| --- | --- | --- | --- | --- | --- | --- | --- | --- | --- | --- |
| **Outcome** | **HR(CI)_present_** | **p.adj_present_** | **Sig_present_** | **HR(CI)_absent_** | **p.adj_absent_** | **Sig_absent_** | **RHR** | **p.adj_RHR_** | **Comparison** | **Organ System Description** |
| All-cause mortality | 1.48  (1.38 - 1.59) | 9.75E-26 | *** | 1.51  (1.39 - 1.64) | 1.06E-21 | *** | 0.98 | 8.84E-01 | Male vs. Female |  |
| CIR | 1.13  (0.93 - 1.38) | 3.27E-01 |  | 1.20  (0.92 - 1.56) | 2.10E-01 |  | 0.95 | 8.84E-01 |  | Diseases of the circulatory system |
| DIG | 1.43  (0.86 - 2.38) | 2.81E-01 |  | 1.92  (1.11 - 3.32) | 2.71E-02 | * | 0.74 | 8.74E-01 |  | Diseases of the digestive system |
| EXT | 3.00  (1.30 - 6.95) | 2.59E-02 | * | 4.53  (1.71 - 11.99) | 5.93E-03 | ** | 0.66 | 8.84E-01 |  | External cause codes |
| GEN | 1.75  (0.91 - 3.35) | 1.87E-01 |  | 3.57  (1.47 - 8.70) | 1.02E-02 | * | 0.49 | 5.09E-01 |  | Diseases of the genitourinary system |
| INF | 1.37  (0.74 - 2.51) | 3.92E-01 |  | 1.20  (0.58 - 2.52) | 6.22E-01 |  | 1.13 | 8.84E-01 |  | Certain infectious and parasitic diseases |
| MUS | 0.92  (0.30 - 2.83) | 8.90E-01 |  | 3.30  (0.98 - 11.09) | 6.73E-02 |  | 0.28 | 5.09E-01 |  | Diseases of the musculoskeletal system and connective tissue |
| NEO | 1.35  (1.20 - 1.52) | 3.80E-06 | *** | 1.36  (1.19 - 1.56) | 2.55E-05 | *** | 0.99 | 9.36E-01 |  | Neoplasms |
| NVS | 1.76  (1.34 - 2.32) | 1.93E-04 | *** | 2.59  (2.02 - 3.33) | 3.46E-13 | *** | 0.68 | 4.24E-01 |  | Diseases of the nervous system |
| RSP | 1.11  (0.81 - 1.53) | 5.57E-01 |  | 1.56  (1.10 - 2.21) | 2.05E-02 | * | 0.71 | 5.09E-01 |  | Diseases of the respiratory system |
| **Supplementary Table 16b. Differences in Associations between Post-acute Mortality Risks in Specific Disorders and Overall COVID-19 in Comparisons of Subgroup (Male vs. Female).** | | | | | | | | | | |
| **Outcome** | **HR(CI)_present_** | **p.adj_present_** | **Sig_present_** | **HR(CI)_absent_** | **p.adj_absent_** | **Sig_absent_** | **RHR** | **p.adj_RHR_** | **Comparison** | **CCSR Category Description** |
| CIR009 | 0.70  (0.45 - 1.07) | 2.15E-01 |  | 1.11  (0.59 - 2.08) | 7.84E-01 |  | 0.63 | 6.77E-01 | Male vs. Female | Acute myocardial infarction |
| CIR011 | 1.23  (0.84 - 1.81) | 4.63E-01 |  | 1.33  (0.70 - 2.53) | 6.34E-01 |  | 0.93 | 9.04E-01 |  | Coronary atherosclerosis and other heart disease |
| CIR021 | 1.22  (0.59 - 2.49) | 6.29E-01 |  | 1.11  (0.60 - 2.04) | 7.84E-01 |  | 1.10 | 9.04E-01 |  | Acute hemorrhagic cerebrovascular disease |
| CIR026 | 4.17  (1.62 - 10.69) | 1.81E-02 | * | 3.76  (1.47 - 9.63) | 1.48E-02 | * | 1.11 | 9.04E-01 |  | Peripheral and visceral vascular disease |
| EXT020 | 4.61  (1.74 - 12.18) | 1.81E-02 | * | 4.19  (1.57 - 11.20) | 1.27E-02 | * | 1.10 | 9.04E-01 |  | External cause codes: intent of injury, accidental/unintentional |
| EXT029 | 3.52  (1.41 - 8.82) | 2.57E-02 | * | 4.19  (1.57 - 11.19) | 1.27E-02 | * | 0.84 | 9.04E-01 |  | External cause codes: subsequent encounter |
| NEO012 | 0.74  (0.40 - 1.39) | 4.85E-01 |  | 1.26  (0.56 - 2.84) | 7.84E-01 |  | 0.59 | 6.89E-01 |  | Gastrointestinal cancers - esophagus |
| NEO015 | 1.52  (1.01 - 2.27) | 1.29E-01 |  | 1.10  (0.71 - 1.72) | 7.84E-01 |  | 1.37 | 6.89E-01 |  | Gastrointestinal cancers - colorectal |
| NEO018 | 2.02  (0.81 - 5.01) | 2.59E-01 |  | 2.51  (1.06 - 5.93) | 7.27E-02 |  | 0.81 | 9.04E-01 |  | Gastrointestinal cancers - bile duct |
| NEO025 | 2.12  (0.98 - 4.60) | 1.49E-01 |  | 1.13  (0.40 - 3.22) | 8.20E-01 |  | 1.88 | 6.89E-01 |  | Skin cancers - melanoma |
| NEO043 | 1.03  (0.55 - 1.93) | 9.35E-01 |  | 4.19  (1.16 - 15.10) | 6.37E-02 |  | 0.24 | 3.22E-01 |  | Urinary system cancers - bladder |
| NEO048 | 1.44  (0.71 - 2.90) | 4.69E-01 |  | 1.19  (0.59 - 2.41) | 7.84E-01 |  | 1.21 | 9.04E-01 |  | Nervous system cancers - brain |
| NEO051 | 0.85  (0.53 - 1.39) | 5.91E-01 |  | 0.82  (0.46 - 1.46) | 7.40E-01 |  | 1.05 | 9.04E-01 |  | Endocrine system cancers - pancreas |
| NEO071 | 1.34  (0.67 - 2.71) | 5.27E-01 |  | 3.91  (2.00 - 7.66) | 6.29E-04 | *** | 0.34 | 2.81E-01 |  | Malignant neoplasm, unspecified |
| NVS004 | 2.34  (1.54 - 3.56) | 1.19E-03 | ** | 3.05  (1.72 - 5.39) | 7.86E-04 | *** | 0.77 | 8.38E-01 |  | Parkinson`s disease |
| NVS011 | 1.77  (1.18 - 2.67) | 2.57E-02 | * | 2.74  (2.00 - 3.76) | 7.42E-09 | *** | 0.65 | 3.56E-01 |  | Neurocognitive disorders |
| RSP008 | 0.86  (0.54 - 1.37) | 5.91E-01 |  | 1.98  (1.28 - 3.07) | 9.06E-03 | ** | 0.43 | 1.82E-01 |  | Chronic obstructive pulmonary disease and bronchiectasis |
| RSP016 | 0.49  (0.17 - 1.42) | 3.39E-01 |  | 1.54  (0.66 - 3.60) | 5.72E-01 |  | 0.32 | 3.56E-01 |  | Other specified and unspecified lower respiratory disease |
| **Supplementary Table 16c. Differences in Associations between Overall Mortality Risks in Organ Systems and Overall COVID-19 in Comparisons of Subgroup (Male vs. Female).** | | | | | | | | | | |
| **Outcome** | **HR(CI)_present_** | **p.adj_present_** | **Sig_present_** | **HR(CI)_absent_** | **p.adj_absent_** | **Sig_absent_** | **RHR** | **p.adj_RHR_** | **Comparison** | **CCSR Category Description** |
| All-cause mortality | 2.45  (2.31 - 2.59) | 1.23E-201 | *** | 2.32  (2.16 - 2.48) | 2.72E-125 | *** | 1.06 | 4.08E-01 | Male vs. Female |  |
| CIR | 1.35  (1.14 - 1.61) | 2.11E-03 | ** | 1.43  (1.12 - 1.82) | 6.27E-03 | ** | 0.95 | 7.59E-01 |  | Diseases of the circulatory system |
| DIG | 1.60  (1.03 - 2.49) | 5.49E-02 |  | 2.93  (1.85 - 4.64) | 1.48E-05 | *** | 0.55 | 3.88E-01 |  | Diseases of the digestive system |
| END | 1.74  (0.64 - 4.69) | 3.32E-01 |  | 2.17  (0.78 - 6.03) | 1.65E-01 |  | 0.80 | 7.59E-01 |  | Endocrine nutritional and metabolic diseases |
| EXT | 2.91  (1.36 - 6.24) | 1.45E-02 | * | 4.01  (1.55 - 10.36) | 6.27E-03 | ** | 0.73 | 7.59E-01 |  | External cause codes |
| GEN | 2.10  (1.18 - 3.74) | 2.40E-02 | * | 4.01  (1.76 - 9.14) | 1.93E-03 | ** | 0.52 | 4.08E-01 |  | Diseases of the genitourinary system |
| INF | 1.81  (1.08 - 3.03) | 4.08E-02 | * | 1.03  (0.49 - 2.15) | 9.39E-01 |  | 1.76 | 4.08E-01 |  | Certain infectious and parasitic diseases |
| INJ | 1.26  (0.60 - 2.65) | 5.85E-01 |  | 1.55  (0.58 - 4.12) | 4.12E-01 |  | 0.81 | 7.59E-01 |  | Injury poisoning and certain other consequences of external causes |
| MUS | 1.04  (0.38 - 2.84) | 9.32E-01 |  | 3.52  (1.10 - 11.20) | 4.46E-02 | * | 0.30 | 4.08E-01 |  | Diseases of the musculoskeletal system and connective tissue |
| NEO | 1.48  (1.32 - 1.65) | 2.94E-11 | *** | 1.55  (1.37 - 1.76) | 9.95E-12 | *** | 0.95 | 7.59E-01 |  | Neoplasms |
| NVS | 1.79  (1.38 - 2.32) | 3.80E-05 | *** | 2.81  (2.22 - 3.57) | 7.12E-17 | *** | 0.64 | 1.40E-01 |  | Diseases of the nervous system |
| RSP | 1.29  (0.96 - 1.73) | 1.20E-01 |  | 1.75  (1.27 - 2.41) | 1.63E-03 | ** | 0.74 | 4.08E-01 |  | Diseases of the respiratory system |
| **Supplementary Table 16d. Differences in Associations between Overall Mortality Risks in Specific Disorders and Overall COVID-19 in Comparisons of Subgroup (Male vs. Female).** | | | | | | | | | | |
| **Outcome** | **HR(CI)_present_** | **p.adj_present_** | **Sig_present_** | **HR(CI)_absent_** | **p.adj_absent_** | **Sig_absent_** | **RHR** | **p.adj_RHR_** | **Comparison** | **CCSR Category Description** |
| CIR003 | 1.10  (0.37 - 3.29) | 8.97E-01 |  | 2.57  (0.86 - 7.65) | 1.63E-01 |  | 0.43 | 7.42E-01 | Male vs. Female | Nonrheumatic and unspecified valve disorders |
| CIR009 | 0.89  (0.62 - 1.28) | 5.78E-01 |  | 1.24  (0.70 - 2.20) | 5.69E-01 |  | 0.72 | 8.03E-01 |  | Acute myocardial infarction |
| CIR011 | 1.42  (1.00 - 2.02) | 1.16E-01 |  | 1.53  (0.83 - 2.83) | 2.67E-01 |  | 0.93 | 9.36E-01 |  | Coronary atherosclerosis and other heart disease |
| CIR021 | 1.54  (0.84 - 2.85) | 2.51E-01 |  | 1.22  (0.69 - 2.17) | 5.75E-01 |  | 1.26 | 8.67E-01 |  | Acute hemorrhagic cerebrovascular disease |
| CIR024 | 1.60  (0.45 - 5.63) | 5.65E-01 |  | 4.59  (1.10 - 19.24) | 7.69E-02 |  | 0.35 | 7.42E-01 |  | Other and ill-defined cerebrovascular disease |
| CIR026 | 4.23  (1.81 - 9.85) | 1.23E-02 | * | 4.50  (1.86 - 10.87) | 4.85E-03 | ** | 0.94 | 9.39E-01 |  | Peripheral and visceral vascular disease |
| DIG017 | 2.95  (0.93 - 9.36) | 1.36E-01 |  | 6.59  (1.40 - 30.97) | 4.07E-02 | * | 0.45 | 8.03E-01 |  | Biliary tract disease |
| DIG025 | 2.27  (0.76 - 6.79) | 2.42E-01 |  | 3.72  (1.38 - 10.07) | 2.52E-02 | * | 0.61 | 8.67E-01 |  | Other specified and unspecified gastrointestinal disorders |
| END016 | 3.47  (0.98 - 12.32) | 1.22E-01 |  | 2.79  (0.81 - 9.55) | 1.76E-01 |  | 1.25 | 9.36E-01 |  | Other specified and unspecified nutritional and metabolic disorders |
| EXT020 | 3.56  (1.49 - 8.54) | 2.12E-02 | * | 4.19  (1.57 - 11.20) | 1.54E-02 | * | 0.85 | 9.36E-01 |  | External cause codes: intent of injury, accidental/unintentional |
| EXT029 | 2.96  (1.28 - 6.85) | 3.63E-02 | * | 4.19  (1.57 - 11.19) | 1.54E-02 | * | 0.71 | 8.67E-01 |  | External cause codes: subsequent encounter |
| GEN004 | 3.47  (1.54 - 7.82) | 1.93E-02 | * | 4.31  (1.28 - 14.50) | 4.07E-02 | * | 0.80 | 9.36E-01 |  | Urinary tract infections |
| INF003 | 1.75  (0.63 - 4.86) | 4.05E-01 |  | 2.12  (0.61 - 7.37) | 3.28E-01 |  | 0.83 | 9.36E-01 |  | Bacterial infections |
| NEO012 | 0.78  (0.43 - 1.40) | 5.36E-01 |  | 1.38  (0.66 - 2.88) | 4.85E-01 |  | 0.56 | 7.42E-01 |  | Gastrointestinal cancers - esophagus |
| NEO013 | 1.79  (0.86 - 3.72) | 2.28E-01 |  | 1.88  (0.74 - 4.74) | 2.67E-01 |  | 0.95 | 9.39E-01 |  | Gastrointestinal cancers - stomach |
| NEO015 | 1.69  (1.17 - 2.44) | 2.28E-02 | * | 1.34  (0.90 - 2.00) | 2.36E-01 |  | 1.25 | 8.03E-01 |  | Gastrointestinal cancers - colorectal |
| NEO018 | 2.61  (1.17 - 5.80) | 5.50E-02 |  | 2.37  (1.01 - 5.54) | 9.08E-02 |  | 1.10 | 9.36E-01 |  | Gastrointestinal cancers - bile duct |
| NEO025 | 2.34  (1.14 - 4.81) | 5.52E-02 |  | 1.13  (0.40 - 3.22) | 8.81E-01 |  | 2.07 | 7.42E-01 |  | Skin cancers - melanoma |
| NEO043 | 1.48  (0.85 - 2.58) | 2.51E-01 |  | 4.76  (1.56 - 14.58) | 1.99E-02 | * | 0.31 | 4.84E-01 |  | Urinary system cancers - bladder |
| NEO048 | 1.62  (0.86 - 3.07) | 2.42E-01 |  | 1.24  (0.61 - 2.50) | 6.20E-01 |  | 1.31 | 8.67E-01 |  | Nervous system cancers - brain |
| NEO051 | 1.13  (0.74 - 1.73) | 6.00E-01 |  | 1.04  (0.62 - 1.74) | 9.01E-01 |  | 1.09 | 9.36E-01 |  | Endocrine system cancers - pancreas |
| NEO071 | 1.41  (0.73 - 2.72) | 4.16E-01 |  | 3.76  (2.01 - 7.07) | 5.39E-04 | *** | 0.38 | 4.55E-01 |  | Malignant neoplasm, unspecified |
| NVS004 | 2.20  (1.49 - 3.26) | 2.42E-03 | ** | 2.94  (1.71 - 5.04) | 6.91E-04 | *** | 0.75 | 8.03E-01 |  | Parkinson`s disease |
| NVS006 | 1.46  (0.46 - 4.63) | 5.78E-01 |  | 3.29  (1.39 - 7.80) | 1.99E-02 | * | 0.45 | 7.42E-01 |  | Other specified hereditary and degenerative nervous system conditions |
| NVS011 | 1.77  (1.21 - 2.60) | 1.93E-02 | * | 2.91  (2.14 - 3.94) | 1.75E-10 | *** | 0.61 | 4.55E-01 |  | Neurocognitive disorders |
| RSP002 | 2.32  (1.35 - 4.00) | 1.93E-02 | * | 1.06  (0.44 - 2.54) | 9.01E-01 |  | 2.20 | 7.42E-01 |  | Pneumonia (except that caused by tuberculosis) |
| RSP008 | 1.01  (0.66 - 1.55) | 9.71E-01 |  | 2.25  (1.50 - 3.38) | 6.91E-04 | *** | 0.45 | 2.30E-01 |  | Chronic obstructive pulmonary disease and bronchiectasis |
| RSP016 | 0.69  (0.28 - 1.73) | 5.47E-01 |  | 1.46  (0.63 - 3.38) | 4.85E-01 |  | 0.48 | 7.42E-01 |  | Other specified and unspecified lower respiratory disease |
| HR(CI)_present_ : Hazard ratio and 95% confidence inteval for the risk factor present subgroup with the examined characteristics; p.adj_present_: FDR-adjusted p-value for the risk factor present subgroup; HR(CI)_absent_: Hazard ratio and 95% confidence inteval for the risk factor absent subgroup without the examined characteristics; p.adj_absent_: FDR-adjusted p-value for the risk factor absent subgroup; Sig_present_ and Sig_absent_: * indicates an FDR-adjusted p-value between 0.01 and 0.05, ** indicates between 0.001 and 0.01, and *** indicates a value smaller than 0.001 for the risk factor present and absent subgroups, respectively. RHR: Ratio of hazard ratios for comparisons between the risk factor present and absent subgroups. p.adj_RHR_*: FDR-adjusted p-value for comparisons between the risk factor present and absent subgroups, where a p.adj_RHR_ < 0.05 indicates significant differences.  CCSR: Clinical Classifications Software Refined, a method to classify disease categories. | | | | | | | | | | |

| **Supplementary Table 17: Quantitative Bias Analysis on Associations between Post-acute Mortality Risks in Organ Systems and Overall COVID-19.** | | | | | | |
| --- | --- | --- | --- | --- | --- | --- |
| **Outcome** | **Original HR** | **Moderate Scenario (π^1^ = 0.20, γ^2^ = 1.05)** | | **Extreme Scenario (π^1^ = 0.45, γ^2^ = 1.50)** | | **Organ System Description** |
|  |  | **Correction factor^3^** | **HR_corrected_** | **Correction factor^3^** | **HR_corrected_** |  |
| All-cause mortality | 1.50 | 1.01 | 1.52 | 1.225 | 1.84 |  |
| CIR | 1.22 |  | 1.24 |  | 1.50 | Diseases of the circulatory system |
| DIG | 1.50 |  | 1.52 |  | 1.84 | Diseases of the digestive system |
| END | 1.61 |  | 1.63 |  | 1.97 | Endocrine nutritional and metabolic diseases |
| EXT | 3.75 |  | 3.79 |  | 4.59 | External cause codes |
| GEN | 2.19 |  | 2.21 |  | 2.68 | Diseases of the genitourinary system |
| INF | 1.25 |  | 1.26 |  | 1.53 | Certain infectious and parasitic diseases |
| INJ | 1.05 |  | 1.06 |  | 1.28 | Injury poisoning and certain other consequences of external causes |
| MUS | 1.61 |  | 1.63 |  | 1.98 | Diseases of the musculoskeletal system and connective tissue |
| NEO | 1.38 |  | 1.39 |  | 1.69 | Neoplasms |
| NVS | 2.13 |  | 2.15 |  | 2.61 | Diseases of the nervous system |
| RSP | 1.21 |  | 1.22 |  | 1.48 | Diseases of the respiratory system |
| HR: hazard ratio; HR_corrected_: Bias-corrected HR = Origianl HR * Correct factor.  1. π: Contamination rate, proportion of the reference cohort that is actually infected. 2. γ: Hazard ratio for undocumented infection relative to truly uninfected. 3. Correction factor = π*γ + (1-π). It was derived by:   Suppose the mortality of our reference cohort (λ_ref_) is a mixture of the undocumented infection and truly uninfected population, we have:  λ_ref_ = π*λ_undocumented_ + (1-π)*λ_uninfected_ = π*γ*λ_uninfected_ + (1-π)*λ_uninfected_ = [π*γ + (1-π)]*λ_uninfected_;  For our primary analysis, original HR meausres the effect of COVID-19 in exposure compared to reference cohort = λ_exp_/λ_ref_ = (HR_corrected_*λ_uninfected_)/([π*γ + (1-π)]*λ_uninfected_);  Finally we have: HR_corrected_ = original HR * [π*γ + (1-π)], and therefore the [π*γ + (1-π)] value is set as correction factor. | | | | | | |

| **Supplementary Table 18: Associations between Post-acute Mortality Risks in Organ Systems and Overall COVID-19 Using Negative-Test Restricted Reference Group.** | | | | | |
| --- | --- | --- | --- | --- | --- |
| **Outcome** | **Additional Analysis  (Negative-Test Restricted  Group as Reference)** | | **Original Analysis (General Population without  Known Infection as Reference)** | | **Organ System Description** |
|  | **HR (95% CI)** | **p.adj^1^** | **HR (95% CI)** | **p.adj^1^** |  |
| All-cause mortality | **1.82 (1.73 - 1.91)** | **2.24E-122** | **1.50 (1.42 - 1.58)** | **2.83E-48** |  |
| CIR | **1.26 (1.09 - 1.47)** | **4.75E-03** | **1.22 (1.05 - 1.43)** | **2.17E-02** | Diseases of the circulatory system |
| DIG | **1.65 (1.18 - 2.30)** | **5.41E-03** | **1.50 (1.04 - 2.16)** | **4.26E-02** | Diseases of the digestive system |
| END | 1.66 (0.79 - 3.46) | 1.78E-01 | 1.61 (0.75 - 3.46) | 2.88E-01 | Endocrine nutritional and metabolic diseases |
| EXT | **3.59 (1.94 - 6.64)** | **1.42E-04** | **3.75 (1.98 - 7.09)** | **1.45E-04** | External cause codes |
| GEN | **2.39 (1.45 - 3.92)** | **1.40E-03** | **2.19 (1.29 - 3.71)** | **8.51E-03** | Diseases of the genitourinary system |
| INF | 1.43 (0.93 - 2.19) | 1.26E-01 | 1.25 (0.79 - 1.98) | 3.77E-01 | Certain infectious and parasitic diseases |
| INJ | 1.53 (0.83 - 2.83) | 1.78E-01 | 1.05 (0.52 - 2.10) | 8.94E-01 | Injury poisoning and certain other consequences of external causes |
| MUS | 1.90 (0.91 - 3.96) | 1.14E-01 | 1.61 (0.73 - 3.58) | 2.88E-01 | Diseases of the musculoskeletal system and connective tissue |
| NEO | **1.45 (1.33 - 1.58)** | **1.54E-16** | **1.38 (1.26 - 1.51)** | **6.22E-12** | Neoplasms |
| NVS | **2.15 (1.80 - 2.57)** | **1.54E-16** | **2.13 (1.78 - 2.56)** | **2.66E-15** | Diseases of the nervous system |
| RSP | **1.31 (1.05 - 1.64)** | **2.50E-02** | **1.21 (1.04 - 1.41)** | **2.62E-02** | Diseases of the respiratory system |
| HR: hazard ratio; 95% CI represents the lower and upper 95% confidence interval of HR; p.adj^1^: FDR-adjusted p-value.  1. Statistically significant associations (p.adj < 0.05) were labelled in bold face. | | | | | |

| **Supplementary Table 19: Associations between Post-acute Mortality Risks in Organ Systems and Overall COVID-19 Stratified by Vaccination Status.** | | | | | |
| --- | --- | --- | --- | --- | --- |
| **Outcome** | **Receiving any doses** | | **No vaccination** | | **Organ System Description** |
|  | **HR (95% CI)** | **p.adj^1^** | **HR (95% CI)** | **p.adj^1^** |  |
| All-cause mortality | **1.48 (1.37 - 1.60)** | **7.90E-21** | **1.66 (1.54 - 1.79)** | **7.10E-38** |  |
| CIR | 1.18 (0.94 - 1.47) | 2.32E-01 | **1.33 (1.06 - 1.68)** | **2.17E-02** | Diseases of the circulatory system |
| DIG | 1.76 (1.08 - 2.88) | 5.24E-02 | 1.39 (0.77 - 2.49) | 3.26E-01 | Diseases of the digestive system |
| END | N/A^2^ | | **5.10 (1.87 - 13.92)** | **2.99E-03** | Endocrine nutritional and metabolic diseases |
| EXT | 2.38 (1.01 - 5.61) | 8.68E-02 | **7.88 (3.08 - 20.13)** | **3.85E-05** | External cause codes |
| GEN | 0.92 (0.32 - 2.63) | 8.78E-01 | **4.24 (2.24 - 8.03)** | **2.66E-05** | Diseases of the genitourinary system |
| INF | 0.94 (0.47 - 1.87) | 8.78E-01 | 1.95 (1.02 - 3.74) | 5.96E-02 | Certain infectious and parasitic diseases |
| INJ | 1.31 (0.54 - 3.17) | 6.68E-01 | 0.63 (0.18 - 2.18) | 4.64E-01 | Injury poisoning and certain other consequences of external causes |
| MUS | 1.53 (0.53 - 4.41) | 5.88E-01 | 1.71 (0.54 - 5.41) | 3.96E-01 | Diseases of the musculoskeletal system and connective tissue |
| NEO | **1.35 (1.19 - 1.53)** | **1.33E-05** | **1.40 (1.23 - 1.61)** | **3.29E-06** | Neoplasms |
| NVS | **2.12 (1.61 - 2.79)** | **4.25E-07** | **2.22 (1.70 - 2.91)** | **4.06E-08** | Diseases of the nervous system |
| RSP | **1.22 (1.04 - 1.43)** | **4.16E-02** | **1.20 (1.04 - 1.39)** | **1.96E-02** | Diseases of the respiratory system |
| HR: hazard ratio; 95% CI represents the lower and upper 95% confidence interval of HR; p.adj^1^: FDR-adjusted p-value.  1. FDR correction was separately performed for analysis with different vaccination status; Statistically significant associations (p.adj < 0.05) were labelled in bold face. 2. N/A indicates that the analysis for target outcome cannot proceed due to insufficient number (<5) of events in either the COVID-19 exposure or reference cohort. | | | | | |

| **Supplementary Table 20: Interaction Analysis between Vaccination Status and Overall COVID-19 on Associations with  Post-acute Mortality Risks in Organ Systems.** | | | |
| --- | --- | --- | --- |
| **Outcome** | **HR (95% CI)** | **p.adj^1^** | **Organ System Description** |
| All-cause mortality | 0.94 (0.85 - 1.05) | 5.98E-01 |  |
| CIR | 0.81 (0.59 - 1.10) | 4.22E-01 | Diseases of the circulatory system |
| DIG | 1.40 (0.66 - 2.97) | 6.56E-01 | Diseases of the digestive system |
| END | 0.14 (0.03 - 0.71) | 1.02E-01 | Endocrine nutritional and metabolic diseases |
| EXT | 0.19 (0.06 - 0.64) | 8.34E-02 | External cause codes |
| GEN | 0.27 (0.08 - 0.90) | 1.32E-01 | Diseases of the genitourinary system |
| INF | 0.42 (0.17 - 1.04) | 1.80E-01 | Certain infectious and parasitic diseases |
| INJ | 1.31 (0.28 - 6.16) | 7.98E-01 | Injury poisoning and certain other consequences of external causes |
| MUS | 0.69 (0.21 - 2.24) | 7.11E-01 | Diseases of the musculoskeletal system and connective tissue |
| NEO | 0.97 (0.81 - 1.16) | 7.98E-01 | Neoplasms |
| NVS | 1.15 (0.80 - 1.65) | 6.67E-01 | Diseases of the nervous system |
| RSP | 0.98 (0.61 - 1.56) | 9.19E-01 | Diseases of the respiratory system |
| HR: hazard ratio; 95% CI represents the lower and upper 95% confidence interval of HR; p.adj^1^: FDR-adjusted p-value.  1. Statistically significant associations (p.adj < 0.05) were labelled in bold face. | | | |

| **Supplementary Table 21: Associations between COVID-19 Infection and Post-acute Mortality Risks in Organ Systems  with Follow-up Extended Beyond Reinfection.** | | | | | |
| --- | --- | --- | --- | --- | --- |
| **Outcome** | **Additional Analysis  (Follow-up Extended**  **Beyond Reinfection)** | | **Original Analysis (Follow-up Censored  After Reinfection)** | | **Organ System Description** |
|  | **HR (95% CI)** | **p.adj^1^** | **HR (95% CI)** | **p.adj^1^** |  |
| All-cause mortality | **1.52 (1.44 - 1.61)** | **3.85E-50** | **1.50 (1.42 - 1.58)** | **2.83E-48** |  |
| CIR | **1.23 (1.05 - 1.44)** | **1.89E-02** | **1.22 (1.05 - 1.43)** | **2.17E-02** | Diseases of the circulatory system |
| DIG | **1.52 (1.06 - 2.18)** | **3.63E-02** | **1.50 (1.04 - 2.16)** | **4.26E-02** | Diseases of the digestive system |
| END | 1.62 (0.76 - 3.49) | 2.85E-01 | 1.61 (0.75 - 3.46) | 2.88E-01 | Endocrine nutritional and metabolic diseases |
| EXT | **3.39 (1.76 - 6.56)** | **8.28E-04** | **3.75 (1.98 - 7.09)** | **1.45E-04** | External cause codes |
| GEN | **2.28 (1.31 - 3.95)** | **8.21E-03** | **2.19 (1.29 - 3.71)** | **8.51E-03** | Diseases of the genitourinary system |
| INF | 1.26 (0.79 - 2.00) | 3.55E-01 | 1.25 (0.79 - 1.98) | 3.77E-01 | Certain infectious and parasitic diseases |
| INJ | 0.95 (0.46 - 1.96) | 8.96E-01 | 1.05 (0.52 - 2.10) | 8.94E-01 | Injury poisoning and certain other consequences of external causes |
| MUS | 1.62 (0.73 - 3.59) | 2.85E-01 | 1.61 (0.73 - 3.58) | 2.88E-01 | Diseases of the musculoskeletal system and connective tissue |
| NEO | **1.40 (1.28 - 1.54)** | **7.67E-13** | **1.38 (1.26 - 1.51)** | **6.22E-12** | Neoplasms |
| NVS | **2.16 (1.80 - 2.60)** | **1.39E-15** | **2.13 (1.78 - 2.56)** | **2.66E-15** | Diseases of the nervous system |
| RSP | **1.27 (1.06 - 1.51)** | **1.66E-02** | **1.21 (1.04 - 1.41)** | **2.62E-02** | Diseases of the respiratory system |
| HR: hazard ratio; 95% CI represents the lower and upper 95% confidence interval of HR; p.adj^1^: FDR-adjusted p-value.  1. Statistically significant associations (p.adj < 0.05) were labelled in bold face. | | | | | |

| **Supplementary Table 22: Associations between Post-acute Mortality Risks in Organ Systems and Overall COVID-19 Stratified by Number of SARS-CoV-2 infections.** | | | | | | | |
| --- | --- | --- | --- | --- | --- | --- | --- |
| **Outcome** | **Single Infection** | | **One reinfection** | | **Two or more reinfection** | |  |
|  | **HR (95% CI)** | **p.adj^1^** | **HR (95% CI)** | **p.adj^1^** | **HR (95% CI)** | **p.adj^1^** | **Organ System Description** |
| All-cause mortality | **1.47 (1.39 - 1.55)** | **1.19E-39** | **3.42 (2.72 - 4.29)** | **1.61E-25** | **3.10 (2.33 - 4.13)** | **4.02E-14** |  |
| CIR | **1.21 (1.03 - 1.42)** | **4.37E-02** | 1.32 (0.55 - 3.20) | 5.35E-01 | **2.84 (1.22 - 6.61)** | **1.53E-02** | Diseases of the circulatory system |
| DIG | 1.48 (1.02 - 2.14) | 5.60E-02 | N/A^2^ | | N/A | | Diseases of the digestive system |
| END | 1.67 (0.78 - 3.60) | 2.49E-01 | N/A | | N/A | | Endocrine nutritional and metabolic diseases |
| EXT | **3.32 (1.70 - 6.49)** | **1.37E-03** | N/A | | N/A | | External cause codes |
| GEN | **2.24 (1.28 - 3.91)** | **1.15E-02** | N/A | | N/A | | Diseases of the genitourinary system |
| INF | 1.21 (0.75 - 1.94) | 4.72E-01 | N/A | | N/A | | Certain infectious and parasitic diseases |
| INJ | 0.98 (0.48 - 2.03) | 9.63E-01 | N/A | | N/A | | Injury poisoning and certain other consequences of external causes |
| MUS | 1.66 (0.75 - 3.69) | 2.54E-01 | N/A | | N/A | | Diseases of the musculoskeletal system and connective tissue |
| NEO | **1.36 (1.24 - 1.49)** | **2.72E-10** | **2.25 (1.51 - 3.35)** | **1.16E-04** | **5.06 (2.98 - 8.60)** | **5.23E-09** | Neoplasms |
| NVS | **2.02 (1.67 - 2.44)** | **2.20E-12** | **6.20 (3.40 - 11.28)** | **5.94E-09** | **11.66 (4.69 - 28.96)** | **1.53E-07** | Diseases of the nervous system |
| RSP | **1.17 (1.02 - 1.35)** | **4.74E-02** | **3.83 (1.56 - 9.36)** | **4.10E-03** | **12.43 (5.05 - 30.63)** | **7.09E-08** | Diseases of the respiratory system |
| HR: hazard ratio; 95% CI represents the lower and upper 95% confidence interval of HR; p.adj^1^: FDR-adjusted p-value.  1. FDR correction was separately performed for analysis with different number of reinfections; Statistically significant associations (p.adj < 0.05) were labelled in bold face. 2. N/A indicates that the analysis for target outcome cannot proceed due to insufficient number (<5) of events in either the COVID-19 exposure or reference cohort. | | | | | | | |
